# Early iron supplementation of exclusively breastfed African infants: a proof-of-principle, placebo-controlled, randomised, double-blinded efficacy trial

**DOI:** 10.1101/2023.01.12.22284059

**Authors:** Mamadou Bah, Isabella Stelle, Hans Verhoef, Alasana Saidykhan, Sophie E. Moore, Babucarr Susso, Andrew M. Prentice, Carla Cerami

## Abstract

**Background:** We have previously shown that breastfed Gambian children have depleted their neonatal iron endowment before 6 months. We measured the effect of daily iron supplementation for 14 weeks on serum iron concentration and other iron markers among breastfed Gambian infants.

**Method:** In a double-blind trial, healthy exclusively breastfed rural Gambian infants aged 6 to 10 weeks were identified from vaccination clinics and local communities. Eligible children (n=101) were individually randomised to 14 weeks of daily supplementation with either iron (7·5mg as ferrous sulphate in sorbitol solution) or placebo (sorbitol solution). The primary outcome was serum iron concentration after 99 days of supplementation (98 days intervention plus 1-day washout). We used intention-to-treat analysis with multiple imputation to replace missing values. This trial was registered with clinicaltrials.gov (NCT04751994).

**Findings:** Iron administration increased serum iron concentration (crude difference in means: 2.5 μmol/L; 95%CI: 0·6 to 4·3μmol/L, p=0.0091) and meaningfully improved additional markers of iron and haematological status. There were 10 serious adverse events (5 iron/5 placebo) and 106 non-serious adverse events (54 iron/52 placebo) with no deaths. There were no marked group differences in maternally-reported episodes of diarrhoea, fever, cough, skin infection, eye infection and nasal discharge.

**Interpretation:** In exclusively breastfed infants, early introduction of iron supplements can enhance iron supply to rapidly developing tissues in early infancy and warrants further investigation in large-scale trials with additional measurements of functional outcomes and safety.

**Funding:** UK Department for International Development, Medical Research Council UK, National Institute for Health Research and Care Research, Wellcome Trust.

**Research in context:** *Evidence before the study:* The World Health Organisation (WHO) recommends that infants should be exclusively breastfed for 6 months. Human milk contains very little iron so exclusively breastfed infants are forced to utilise their birth endowment of liver ferritin and fetal haemoglobin to meet the needs of growth and tissue development. In low-income countries many infants are born prematurely, at low birthweight or to iron deficient mothers. These infants start life with low iron reserves and hence frequently become very iron deficient by 6 months of age. In many high-income countries it is recommended that such infants should receive iron supplements from soon after birth. Although not specifically precluded by WHO’s recommendation on exclusive breastfeeding, the provision of supplements is widely viewed as being unnecessary and as undermining the ethos of the recommendation.

*Added value of this study:* In this proof-of-principle trial we demonstrated that providing 7·5mg iron per day to exclusively breastfed Gambian infants from 6 weeks of age substantially improved all markers of iron status at 6 months of age. There were no signals of adverse effects on growth or on infections.

*Implications of all the available evidence:* Serious iron deficiency in many exclusively breastfed infants in low-income countries impairs iron supply to rapidly developing tissues including immune and neural cells and the expanding erythroid pool. Early introduction of iron supplements can reverse this deficiency and warrants further testing in large-scale trials with additional measurements of functional outcomes and safety.

## Introduction

There are increasing doubts whether the iron supplied by exclusive breastfeeding is adequate to meet infant requirements in the first 6 months of life. The iron content of human milk is low (at ∼0·35 mg/L).^1^ So a young infant typically receiving 750-1,000 mL breastmilk per day will receive around 0·25-0·35 mg iron daily. In calculating iron requirements, the US Institute of Medicine (IOM) assumed that breastmilk iron is adequate and hence set the requirement at 0·27 mg/d for 0-6 months.^2^ At 6 months, based upon factorial calculations of iron losses and the needs for growth, the IOM recommendation jumps 40-fold to 11 mg/d.^2^ This non-physiological disjuncture is an artefact, and back-extrapolation of factorial calculations to ages below 6 months, when infants are growing faster, would indicate a true need of at least 10 mg/d if supplied by diet. Breastmilk iron is largely bound to lactoferrin and benefits from efficient receptor-mediated absorption compared to non-milk iron but, even allowing for this, breastmilk provides an order of magnitude less iron than an infant requires.^1^ Hence, exclusively breastfed infants must draw on their endowed iron reserves (liver ferritin and recycled iron from fetal haemoglobin) to support their daily needs and for growth (especially of their expanding red cell mass).^3^

The fetus accrues most of its iron during the final trimester of pregnancy and is dependent upon an adequate maternal iron status to supply these needs.^4^ Infants born prematurely, at low birthweight, or to iron deficient mothers, and whose cord is clamped rapidly after birth, start life with smaller iron stores and hence a lower capacity to subsidise the iron requirements of growth and metabolism whilst being breastfed. Thus, in many high-income countries, it is recommended that they should receive supplemental iron at rates of 2-4 mg/kg when given orally, starting soon after birth.^1^

In establishing the recommendation that infants should be exclusively breastfed until 6 months the World Health Organisation’s expert panel acknowledged evidence that many children would become iron deficient if not given supplemental iron but offered no solution.^5^

In low-income settings infants are much more likely to be born with low iron reserves and are also more likely to be exclusively breastfed in early infancy.^6^ In two independent community cohorts we have previously shown that breastfed Gambian children have utilised their birth endowment of iron before 6 months, and that functional indicators of iron deficiency (concentrations of transferrin and soluble transferrin receptor) start to rise at 2 months of age.^3^ Haemoglobin falls and the proportion of anaemic children rises. ^3^ Most remarkably, concentrations of serum iron were within the standard range at birth (∼15 μmol/L) but fell rapidly to levels below 8 μmol/L at 2 months and around 2-4 μmol/L by 6 months; these are well below the reference levels for well-nourished children (10-20 μmol/L).^3^

All human cells require iron and many tissues undergoing critical phases of development in a young infant (eg, neurones and lineages involved in adaptive immunity) have high iron needs.^7^ These cells can only access iron from circulating plasma and hence the very low levels of serum iron in breastfed infants likely constitute a substantial limitation on development.^3^

In this proof-of-principle, placebo-controlled, randomised trial we measured the effect of supplying 7·5 mg/d iron as ferrous sulphate for 14 weeks to exclusively breastfed Gambian infants aged 6-10 weeks, with a view to improve their iron status. The primary outcome was serum iron, and safety was assessed by daily review of possible adverse events. Secondary outcomes included a range of additional markers of iron and haematological status.

## Subjects and methods

### Trial design

This was a double-blind, individually randomised placebo-controlled trial conducted in Jarra West in the Lower River Region of The Gambia. Malaria used to be endemic but is now virtually non-existent in this area, as indicated by the low prevalence of *Plasmodium* infection (0.4%) among under-five children according to the 2019 Demographic and Health Survey (DHS).^8^ Full details of study design are available in the published protocol.^9^

The study was approved by the Scientific Coordinating Committee of the Medical Research Council (MRC) Unit The Gambia and the Joint Gambia Government MRC Ethics Committee and the Ethics Committee at the London School of Hygiene and Tropical Medicine (SCC19092). The trial was conducted according to the principles of Good Clinical Practice, had oversight from a Data Safety and Monitoring Board and a Trial Steering Committee and was monitored by the Clinical Trials Office at the MRCG@LSHTM. No interim analysis for efficacy was done. This trial was registered with clincaltrials.gov (NCT04751994).

### Recruitment

Participants were identified from vaccination clinics and in the communities by field workers. Following sensitisation those willing to participate were followed up in their homes for consenting. After obtaining written informed consent, participants were invited to the Jarra Soma Health Centre and screened for eligibility before randomisation (Day 0). All trial participants were generally healthy and were exclusively breastfed as reported by their mothers. Eligibility criteria included willingness to stay in the participating communities and to adhere to the trial protocol. Formula-fed infants and those with fever, haemoglobin <7·0 g/dL, or acute or chronic illnesses were excluded.

On Day 0 (baseline), weight (Seca 336 infant scale, Hamburg, Germany; to the nearest 10 g) and length (Seca 417 lengths board; to the nearest 1 mm) were measured. Eligibility screening and blood sample collection was done by the research nurses. Participants that met all the inclusion criteria were enrolled and a venous blood sample was collected and stored on ice until analysis. Full blood count and reticulocyte parameters were analysed in EDTA stabilised blood within 4 hours after collection using an automated haematology analyser (Sysmex XN-Series 1500, Sysmex Corporation, Japan), and a serum sample was stored at –80°C for subsequent measurement of iron and inflammation markers. Children were randomised either on Day 0 or 1 after haemoglobin concentration was measured and eligibility to randomisation had been fully established.

### Randomisation and blinding

Permuted block randomisation (with a fixed block size of six) with an allocation ratio of 1:1 was used. The randomisation sequence was produced using RStudio by the trial statistician and uploaded as an electronic case report form (eCRF) into the RedCap database by the data manager. The iron and placebo groups were sub-divided into 3 groups each (total 6 groups) to further conceal the treatment allocation. The eCRF was a random number generator used to assign each enrolled participant into one of the six intervention codes by the data manager. The interpretation of these codes was only known by the independent pharmacist. The randomisation code was broken after data were cleaned and locked.

The ferrous sulphate supplement (containing 7·5 mg/day iron in sorbitol solution) and placebo (sorbitol solution USP 70%) were supplied in liquid form by Pharmacy Innovation (NC, USA). To ensure masking, both supplements were repackaged in identical bottles by a pharmacist independent from the study.

### Follow up

Enrolled participants were provided with infant mosquito nets to decrease the risk of subsequent malaria infection. Participants were supplemented with 0·5 mL supplemental solution (7·5 mg/day iron or placebo) by fieldworkers from Day 1 to Day 98 (14 weeks) with the support of parents/guardians. The investigational products were returned daily to the field office and the storage temperature was monitored. Daily iron supplementation, daily health and weekly feeding questionnaires were completed for 98 days.

An endline sample of venous blood (3·0 mL) was collected on Day 99 to allow one day of wash-out. The full blood count and reticulocyte parameters were assessed, and a serum sample was collected as described for baseline. Post-supplementation follow-up to assess health status was continued daily up to Day 112 (16 weeks).

### Laboratory measurements

Baseline and endline serum samples were used to measure concentrations of iron, ferritin, soluble transferrin receptor (sTfR), unsaturated iron binding capacity (UIBC), transferrin saturation, C-reactive protein (CRP) and α_1_-acid glycoprotein (AGP), using an automated biochemistry analyser (Cobas Integra 400 plus; Roche Diagnostics, Rotkreuz, Switzerland). Serum hepcidin (DRG instruments, manufacturer code HS EIA-5782, Marburg, Germany), erythropoietin (Human Erythropoietin SimpleStep Kit, ABCAM, ab274397, Cambridge, UK) and erythroferrone (IE, Intrinsic Life Science, SKU# ERF-001 California, USA) concentrations were all analysed by ELISA.

### Outcomes

The primary outcome was serum iron concentration at Day 99. Secondary outcomes included anaemia, iron deficiency and iron deficiency anaemia, erythroferrone, erythropoietin, hepcidin with other iron and haematological markers (see full list of variables in **Tables 1 and 2**) at Day 99. Safety was monitored by daily assessment of morbidity and adverse events during the intervention period. Adverse events were defined as any untoward or unfavourable medical occurrence whether considered related to the child’s participation in the study or not. Serious adverse events were defined as adverse events that were life-threatening, resulted in death, required hospitalization or prolongation of hospitalisation, or resulted in a persistent and significant disability or incapacity, and were always investigated by the trial clinician. In addition, we daily asked mothers about the occurrence of specific events of interest (diarrhoea, fever, cough, vomiting, skin infection, eye infection, nasal discharge).

**Table 1.** Characteristics of the study population at baseline (intention-to-treat population)

| Baseline variable | Iron | Placebo |
| --- | --- | --- |
| n, as randomised | 50 | 51 |
| Age, weeks |  |  |
| 6-6.99 weeks | 29 (58.0%) | 25 (49.0%) |
| 7-7.99 weeks | 12 (24.0%) | 12 (23.5%) |
| 8-8.99 weeks | 3 (6.0%) | 8 (15.7%) |
| 9+ weeks | 6 (12.0%) | 6 (11.8%) |
| Female | 26 (52.0%) | 29 (56.9%) |
| Number of children born to the mother | 3.9 (2.2) | 3.9 (2.1) |
| Serum iron, $\mu\text{mol/L}$ | 12.6 (4.5) | 12.5 (6.9) |
| Serum ferritin, $\mu\text{g/L}$ † | 283.2 (1.81) | 262.6 (1.76) |
| Serum soluble transferrin receptor, $\text{mg/L}$ † | 2.81 (1.28) | 2.76 (1.30) |
| Serum sTfR/$\log_{10}(\text{ferritin})$ index † | 1.15 (1.34) | 1.15 (1.31) |
| Serum transferrin, $\text{g/L}$ | 2.2 (0.4) | 2.1 (0.4) |
| UIBC, $\mu\text{mol/L}$ | 37.0 (11.0) | 35.2 (11.0) |
| TSAT, % † | 24.0 (1.65) | 22.6 (2.00) |
| Haemoglobin, $\text{g/dL}$ † | 10.9 (1.6) | 11.0 (1.2) |
| MCV, fL | 89.6 (5.3) | 89.2 (5.4) |
| MCH, pg | 29.4 (1.7) | 28.9 (2.1) |
| MCHC, $\text{g/dL}$ | 32.9 (1.4) | 32.6 (1.3) |
| Reticulocyte count, $\times 10^9/\text{L}$ † | 0.07 (1.44) | 0.07 (1.49) |
| Reticulocyte count, % † | 1.83 (1.57) | 1.91 (1.54) |
| Immature reticulocyte fraction, % † | 17.2 (1.50) | 16.4 (1.52) |
| Reticulocyte haemoglobin, pg | 31.8 (1.8) | 31.3 (2.4) |
| Serum hepcidin, $\mu\text{g/L}$ † | 20.3 (2.21) | 19.6 (2.36) |
| Serum erythropoietin, IU/L † | 12.7 (2.05) | 10.8 (1.85) |
| Serum $\alpha_1$-acid glycoprotein, g/L † | 0.66 (1.48) | 0.61 (1.80) |
| Serum erythroferrone, $\mu\text{g/L}$ * § | | |
| Below or equal to LOQ ($\leq 0.16 \mu\text{g/L}$) | 66.0% (33) | 74.5% (38) |
| 0.16-0.499 | 22.0% (11) | 3.9% (2) |
| 0.5-0.99 | 10.0% (5) | 5.9% (3) |
| 1.0+ | 2.0% (1) | 15.7% (8) |
| Serum C-reactive protein * |  |  |
| Below or equal to LOQ ($\leq 0.60 \text{ mg/L}$) | 34 (68.0%) | 33 (64.7%) |
| 0.60-4.99 mg/L | 12 (24.0%) | 12 (23.5%) |
| 5-14.99 mg/L | 2 (4.0%) | 1 (2.0%) |
| 15+ mg/L | 2 (4.0%) | 5 (9.8%) |
| Height-for-age z-score, SD | -0.59 (1.18) | -0.68 (1.59) |
| Weight-for-age z-score, SD | -0.71 (1.19) | -0.70 (1.25) |
| Weight-for-height z-score, SD | -0.22 (1.53) | -0.11 (1.11) |

LOQ: limit of quantification; MCH: mean corpuscular haemoglobin; MCHC: mean corpuscular haemoglobin concentration; MCV: mean corpuscular volume; sTfR: soluble transferrin receptor; TSAT: transferrin saturation; UIBC: unsaturated iron-binding capacity. Values indicate n, mean (SD); n, geometric mean (geometric SD); † or prevalence n (%). \* median (25-, and 75%-percentiles); The geometric SD is a dimensionless, multiplicative factor such that dividing or multiplication of the geometric mean by this ratio indicates a variation that is equivalent to subtraction or addition of one standard deviation on a log- transformed scale. ‡ Excluding one observation with an outlying value (86.8).

**Table 2.**
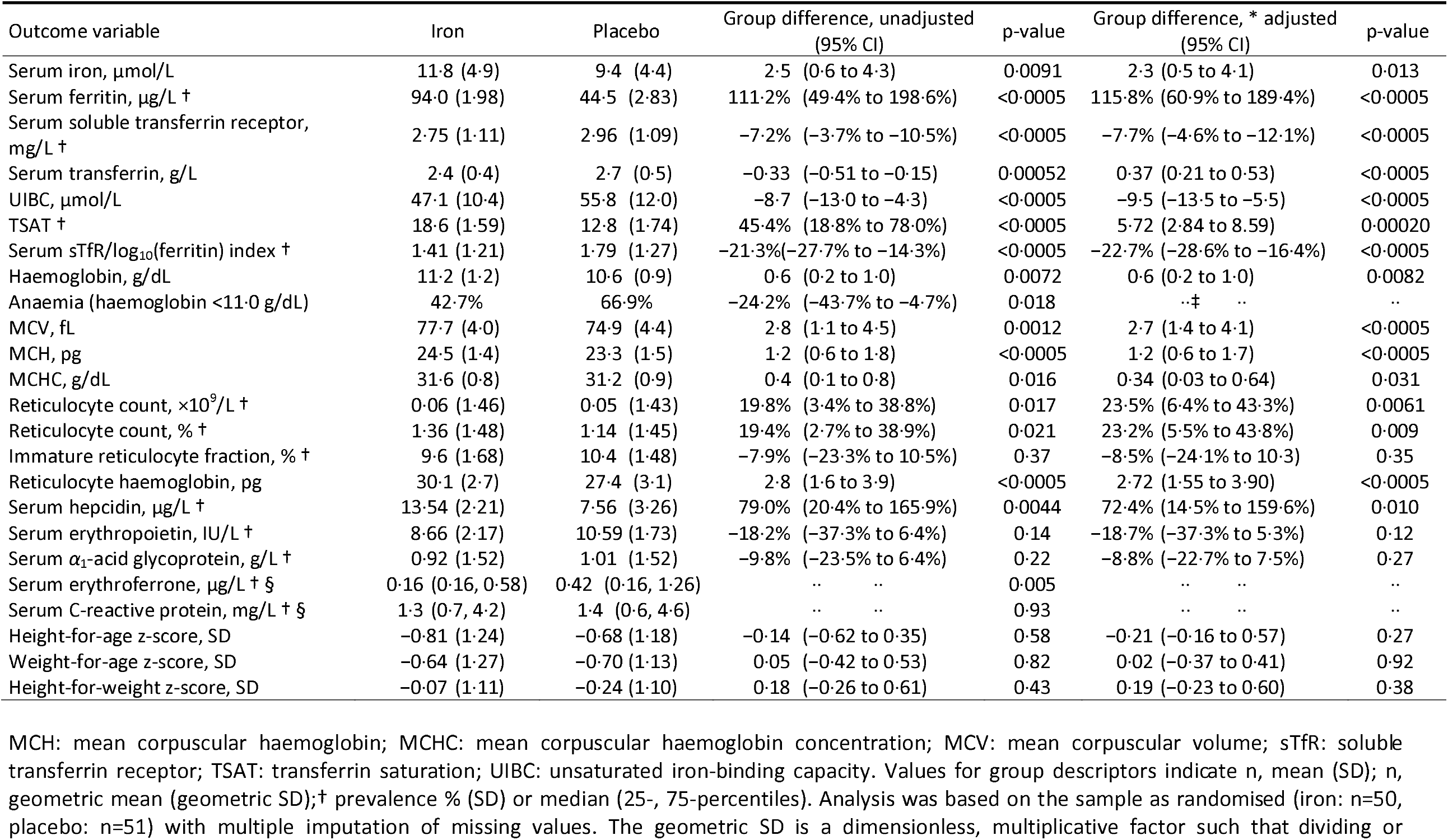
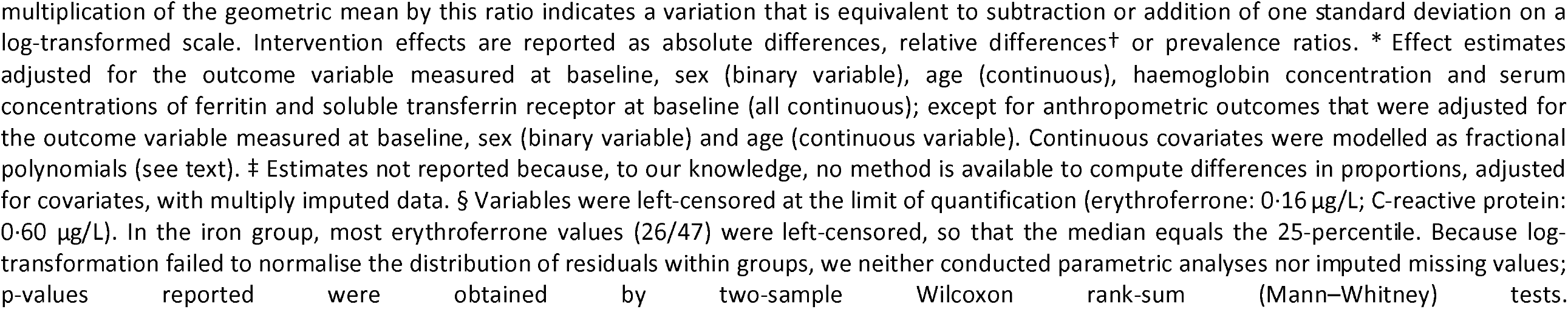
Effect of iron supplementation on primary and secondary outcomes (intention-to-treat population)

| Outcome variable | Iron | Placebo | Group difference, unadjusted<br>(95% CI) | p-value | Group difference, * adjusted<br>(95% CI) | p-value |
| --- | --- | --- | --- | --- | --- | --- |
| Serum iron, $\mu\text{mol/L}$ | 11.8 (4.9) | 9.4 (4.4) | 2.5 (0.6 to 4.3) | 0.0091 | 2.3 (0.5 to 4.1) | 0.013 |
| Serum ferritin, $\mu\text{g/L}$ † | 94.0 (1.98) | 44.5 (2.83) | 111.2% (49.4% to 198.6%) | <0.0005 | 115.8% (60.9% to 189.4%) | <0.0005 |
| Serum soluble transferrin receptor,<br>$\text{mg/L}$ † | 2.75 (1.11) | 2.96 (1.09) | -7.2% (-3.7% to -10.5%) | <0.0005 | -7.7% (-4.6% to -12.1%) | <0.0005 |
| Serum transferrin, $\text{g/L}$ | 2.4 (0.4) | 2.7 (0.5) | -0.33 (-0.51 to -0.15) | 0.00052 | 0.37 (0.21 to 0.53) | <0.0005 |
| UIBC, $\mu\text{mol/L}$ | 47.1 (10.4) | 55.8 (12.0) | -8.7 (-13.0 to -4.3) | <0.0005 | -9.5 (-13.5 to -5.5) | <0.0005 |
| TSAT † | 18.6 (1.59) | 12.8 (1.74) | 45.4% (18.8% to 78.0%) | <0.0005 | 5.72 (2.84 to 8.59) | 0.00020 |
| Serum sTfR/$\log_{10}$(ferritin) index † | 1.41 (1.21) | 1.79 (1.27) | -21.3% (-27.7% to -14.3%) | <0.0005 | -22.7% (-28.6% to -16.4%) | <0.0005 |
| Haemoglobin, $\text{g/dL}$ | 11.2 (1.2) | 10.6 (0.9) | 0.6 (0.2 to 1.0) | 0.0072 | 0.6 (0.2 to 1.0) | 0.0082 |
| Anaemia (haemoglobin <11.0 $\text{g/dL}$) | 42.7% | 66.9% | -24.2% (-43.7% to -4.7%) | 0.018 | .. ‡ .. | .. |
| MCV, $\text{fL}$ | 77.7 (4.0) | 74.9 (4.4) | 2.8 (1.1 to 4.5) | 0.0012 | 2.7 (1.4 to 4.1) | <0.0005 |
| MCH, $\text{pg}$ | 24.5 (1.4) | 23.3 (1.5) | 1.2 (0.6 to 1.8) | <0.0005 | 1.2 (0.6 to 1.7) | <0.0005 |
| MCHC, $\text{g/dL}$ | 31.6 (0.8) | 31.2 (0.9) | 0.4 (0.1 to 0.8) | 0.016 | 0.34 (0.03 to 0.64) | 0.031 |
| Reticulocyte count, $\times 10^9/\text{L}$ † | 0.06 (1.46) | 0.05 (1.43) | 19.8% (3.4% to 38.8%) | 0.017 | 23.5% (6.4% to 43.3%) | 0.0061 |
| Reticulocyte count, % † | 1.36 (1.48) | 1.14 (1.45) | 19.4% (2.7% to 38.9%) | 0.021 | 23.2% (5.5% to 43.8%) | 0.009 |
| Immature reticulocyte fraction, % † | 9.6 (1.68) | 10.4 (1.48) | -7.9% (-23.3% to 10.5%) | 0.37 | -8.5% (-24.1% to 10.3) | 0.35 |
| Reticulocyte haemoglobin, $\text{pg}$ | 30.1 (2.7) | 27.4 (3.1) | 2.8 (1.6 to 3.9) | <0.0005 | 2.72 (1.55 to 3.90) | <0.0005 |
| Serum hepcidin, $\mu\text{g/L}$ † | 13.54 (2.21) | 7.56 (3.26) | 79.0% (20.4% to 165.9%) | 0.0044 | 72.4% (14.5% to 159.6%) | 0.010 |
| Serum erythropoietin, $\text{IU/L}$ † | 8.66 (2.17) | 10.59 (1.73) | -18.2% (-37.3% to 6.4%) | 0.14 | -18.7% (-37.3% to 5.3%) | 0.12 |
| Serum $\alpha_1$-acid glycoprotein, $\text{g/L}$ † | 0.92 (1.52) | 1.01 (1.52) | -9.8% (-23.5% to 6.4%) | 0.22 | -8.8% (-22.7% to 7.5%) | 0.27 |
| Serum erythroferrone, $\mu\text{g/L}$ † § | 0.16 (0.16, 0.58) | 0.42 (0.16, 1.26) | .. .. | 0.005 | .. .. | .. |
| Serum C-reactive protein, $\text{mg/L}$ † § | 1.3 (0.7, 4.2) | 1.4 (0.6, 4.6) | .. .. | 0.93 | .. .. | .. |
| Height-for-age z-score, SD | -0.81 (1.24) | -0.68 (1.18) | -0.14 (-0.62 to 0.35) | 0.58 | -0.21 (-0.16 to 0.57) | 0.27 |
| Weight-for-age z-score, SD | -0.64 (1.27) | -0.70 (1.13) | 0.05 (-0.42 to 0.53) | 0.82 | 0.02 (-0.37 to 0.41) | 0.92 |
| Height-for-weight z-score, SD | -0.07 (1.11) | -0.24 (1.10) | 0.18 (-0.26 to 0.61) | 0.43 | 0.19 (-0.23 to 0.60) | 0.38 |

MCH: mean corpuscular haemoglobin; MCHC: mean corpuscular haemoglobin concentration; MCV: mean corpuscular volume; sTfR: soluble transferrin receptor; TSAT: transferrin saturation; UIBC: unsaturated iron-binding capacity. Values for group descriptors indicate n, mean (SD); n, geometric mean (geometric SD); † prevalence % (SD) or median (25-, 75-percentiles). Analysis was based on the sample as randomised (iron: n=50, placebo: n=51) with multiple imputation of missing values. The geometric SD is a dimensionless, multiplicative factor such that dividing or
multiplication of the geometric mean by this ratio indicates a variation that is equivalent to subtraction or addition of one standard deviation on a log-transformed scale. Intervention effects are reported as absolute differences, relative differences<sup>†</sup> or prevalence ratios. \* Effect estimates adjusted for the outcome variable measured at baseline, sex (binary variable), age (continuous), haemoglobin concentration and serum concentrations of ferritin and soluble transferrin receptor at baseline (all continuous); except for anthropometric outcomes that were adjusted for the outcome variable measured at baseline, sex (binary variable) and age (continuous variable). Continuous covariates were modelled as fractional polynomials (see text). ‡ Estimates not reported because, to our knowledge, no method is available to compute differences in proportions, adjusted for covariates, with multiply imputed data. § Variables were left-censored at the limit of quantification (erythroferrone: 0·16 µg/L; C-reactive protein: 0·60 µg/L). In the iron group, most erythroferrone values (26/47) were left-censored, so that the median equals the 25-percentile. Because log-transformation failed to normalise the distribution of residuals within groups, we neither conducted parametric analyses nor imputed missing values; p-values reported were obtained by two-sample Wilcoxon rank-sum (Mann–Whitney) tests.

### Statistical issues

#### Sample size calculations

Sample size was calculated assuming a geometric SD for serum iron concentration of 2.39 for both groups, based on results from a previous study.^3^ The geometric SD is a dimensionless factor that indicates variation that is equivalent to subtraction or addition of 1 SD on a log-transformed scale.^3^ Assuming <10% drop-out, recruitment targeted 100 infants, providing 80% probability that an increase in geometric mean serum iron concentration by at least 70% in the intervention group compared to the placebo would exclude no effect (ie, ratio of geometric means of 1) from the 95% CI. In absolute terms, assuming a geometric mean serum iron concentration in the placebo group of 4·3 μmol/L,^3^ an increase by 70% would correspond to +3·0 μmol/L.

#### Definitions

Transferrin saturation was calculated as serum iron concentration (μmol/L)/total iron-binding capacity (μmol/L) × 100. Anthropometric z-scores for were derived using the WHO international growth reference.^10^ Anaemia was defined as haemoglobin concentration <11.0 g/dL even though there are no agreed definitions for infants aged below 6 months.^11^ Adherence was defined as the percentage of scheduled days (98) that the supplement/placebo was actually ingested.

#### Group descriptions

Analyses were conducted using Stata version 17 (Stata Corp, College Station, TX) and R version 4.2.1 (https://www.R-project.org/). We visually inspected histograms of variables and residuals by intervention group to assess the shape of their distribution and to identify possible outliers. Skewed variables were normalised by log transformation as appropriate. Groups were described using mean (SD) for normally distributed variables, geometric mean (geometric SD, GSD) for log-transformed data and proportion/percentage for categorical variables. GSD was calculated as the exponentiated SD of the log-transformed variable.

#### Endpoints

The primary endpoint was serum iron concentration at Day 99 in the modified intention-to-treat population (i.e., all participants that were randomised and had received at least one supplemental dose). Secondary efficacy endpoints included other markers of iron status, haemoglobin and reticulocyte haemoglobin, hepcidin, erythroferrone, and erythropoietin.

#### Intervention effects

We estimated crude intervention effects on continuous outcomes by simple linear regression. To assess the potential role of confounding due to imbalances in baseline variables, we also used multiple linear regression to estimate intervention effects adjusted for the outcome variable measured at baseline, sex (binary variable), age (continuous variable), and haemoglobin concentration and serum concentrations of ferritin and soluble transferrin receptor at baseline (all continuous variables); except for anthropometric outcomes that were adjusted for the outcome variable measured at baseline, sex (binary variable) and age (continuous variable). In these analyses, continuous variables were entered as fractional polynomials to allow for possible non-linear relationships with outcome.^12^ In all models, we accounted for heteroscedasticity using robust variance estimators. Intervention effects are reported as absolute differences in means for normally distributed outcomes, or as relative differences in geometric means for log-transformed outcomes. When log-transformation failed to normalise the distribution of residuals within groups, we assessed group differences by two-sample Wilcoxon rank-sum (Mann–Whitney) tests. In per protocol analysis, we used logistic regression models with the *adjrr* command in Stata package *st0306.pkg* to estimate prevalence differences adjusted for baseline characteristics.

As planned, the primary analysis was by modified intention to treat analysis, i.e., excluding children who were randomised but withdrawn before they received the first supplemental dose of iron or placebo. In practice, such withdrawals did not occur, so the primary analysis was de facto by intention-to-treat. To account for missing values, we used multiple imputation (see details of the imputation model in the **Annex**). As supportive analyses, we also report per protocol analyses with pairwise deletion of missing values.

#### Adverse events

We used per protocol analyses of adverse events and daily maternal reports of adverse symptoms of interest. Adverse events were reported as the number of events within each treatment group. No formal statistical analysis was done due to the small number of adverse events.

In the analysis of daily *maternal reports* of adverse symptoms of interest (diarrhoea, fever, cough, vomiting, skin infections, eye infections, nasal discharge), we defined a new episode as one or more successive days that such symptoms were reported, separated from a previous episode by successive days without such symptoms (diarrhoea, 5 days; all other symptoms, 3 days). Thus, new episodes could be recurrent within individual children. Details of case definitions and analysis are provided in the Annex.

## Results

Between 3 August 2021 and 9 March 2022, 103 children were invited, and 101 children were enrolled. Of these, 50 were randomised to iron (26 females) and 51 to placebo (29 females). Two participants were excluded from the study before randomisation (one failed the eligibility criteria and the second was excluded due to technical error) **(Figure 1).** Three participants were lost to follow up, one had a clotted sample (haematology analysis could not done) and one refused to give a sample at endline blood in the per-protocol population. At the start of the study, reticulocyte number, reticulocyte percentage, immature reticulocyte fraction and reticulocyte haemoglobin content were not measured for 17 children (9 intervention; 8 control) due to delays in equipment installation and calibration.

**Figure 1.**
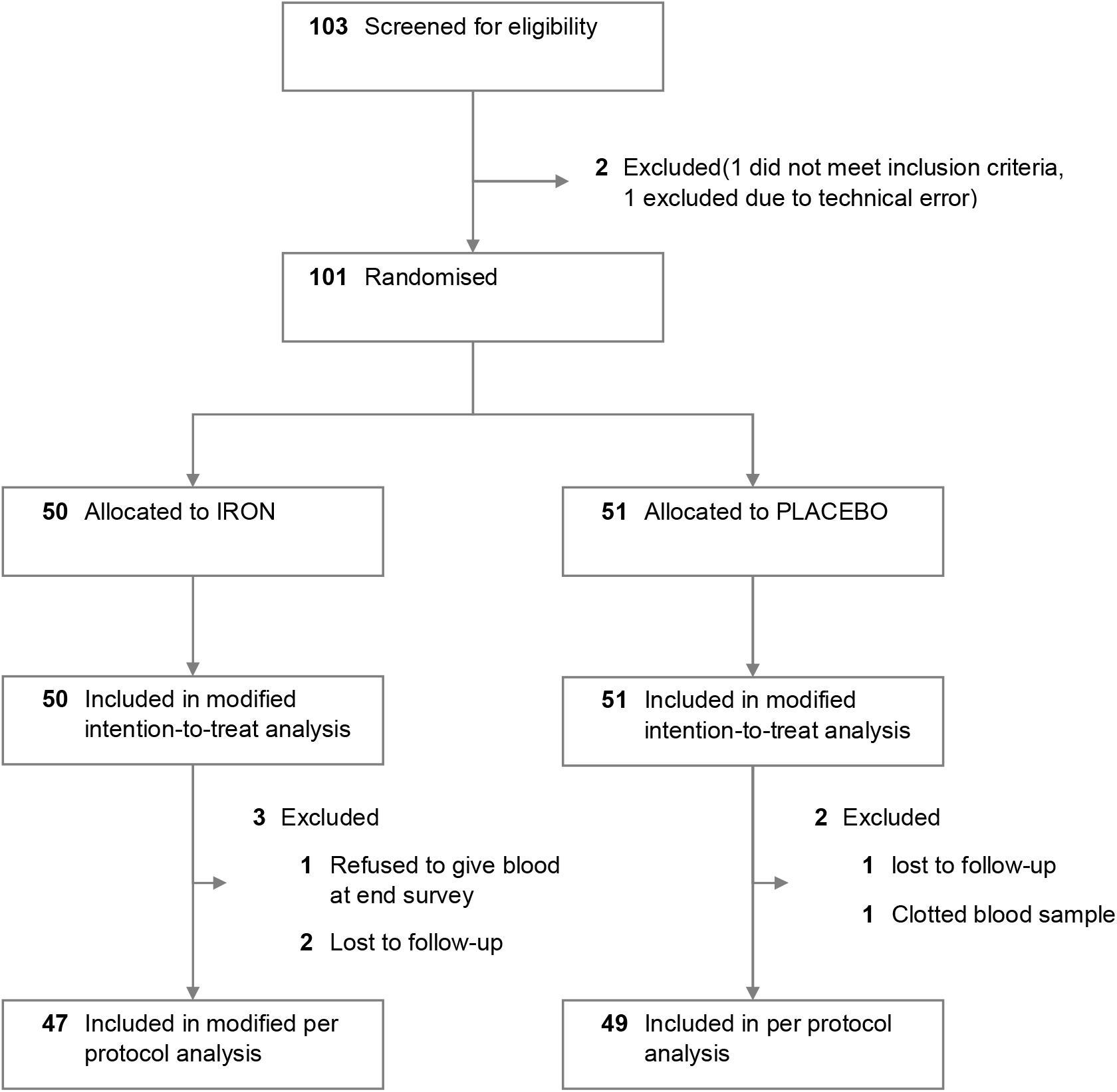
CONSORT diagram.

Intervention groups were similar regarding baseline characteristics **(Table 1)**. Adherence to directly observed daily supplementation was 91% and was similar between groups (iron group: 92.3%; placebo group: 89.1%). Participants travelling outside the study area were responsible for the majority of non-adherence events **(Figure S1)**.

Results of intention to treat analyses are shown in **Table 2** (for results of per protocol analyses: see Annex, **Tables S1 and S2**). After 98 days of supplementation, the primary outcome, serum iron was higher in the iron group compared to placebo (crude difference in means: 2.5 μmol/L; 95%CI: 0·6 to 4·3 μmol/L). All other markers of iron status improved in the treatment group including decreased soluble transferrin receptor (crude difference in geometric means: −7.2%; 95%CI: −3·7 to −10·5%), decreased UIBC (crude difference in means: −8.7 μmol/L; 95%CI: −13·1 to −4·3 μmol/L), increased transferrin saturation (crude difference in geometric means: 45·4%; 95%CI: 18·8 to 78·0%), increased ferritin (crude difference in geometric means: 111·2%; 95%CI: 49·4% to 198·6%) and increased hepcidin (crude difference in geometric means: 79·0%; 95%CI: 20·4% to 165·9%) **(Table 2).** There was strong evidence of higher erythroferrone concentrations in the placebo group compared to the ferrous sulphate group (p=0.0052).

Haemoglobin concentration (crude difference in means: 0.6 g/dL; 95%CI: 0·2 to 1·0 g/dL) and other haematological markers including MCV, MCH, MCHC, reticulocytes haemoglobin and reticulocyte number also increased in the iron supplemented group compared to placebo **(Table 2)**. There was good evidence of decreased prevalence of anaemia (crude prevalence difference: −24·2% (−43·7% to −4·7%). **Figure 2** summarises the benefits for all the iron and haematological markers expressed as standardised differences of means (i.e. expressed as standard deviations for each variable) or as the ratio of geometric means for skewed variables.

**Figure 2.**
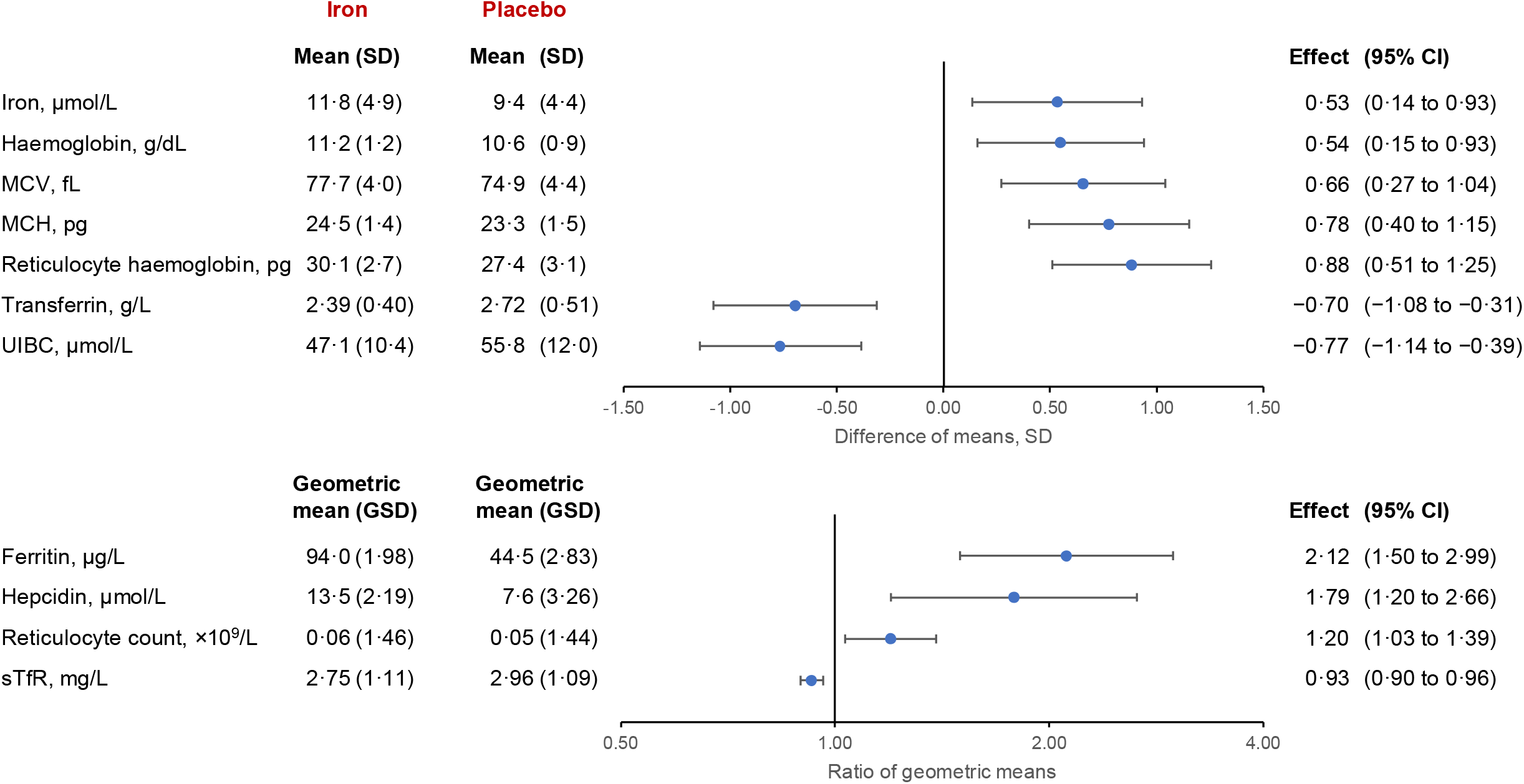
Aggregate effects of iron supplementation on selected outcomes, intention-to-treat analysis. MCH: mean corpuscular haemoglobin; MCHC: mean corpuscular haemoglobin concentration; MCV: mean corpuscular volume; sTfR: soluble transferrin receptor; UIBC: unsaturated iron-binding capacity. For outcomes with normally distributed residuals, effect sizes are standardised, i.e., they are computed as the difference in group means divided by the SD of all imputed values *(upper panel).* For outcomes with log-normally distributed residuals, effect sizes are computed as the ratio of geometric means (*lower panel*). Note that reductions in transferrin, UIBC and sTfR indicate improvedironstatus.

There were no marked differences between groups for immature reticulocyte fractions, erythropoietin, *α*_1_-acid glycoprotein, C-reactive protein, height-for-age z-score, weight-for-age z-score or weight-for-height z-score.

There were no deaths during the intervention period. Adverse events, serious adverse events and self-reported illnesses are reported in **Tables 3, 4 and S4** and **Figure S3**. A total of 10 serious adverse events (5 intervention/5 control) and 106 non-serious adverse events (54 intervention/52 control) were reported and were similar between groups. There was no evidence of differences between groups in the incidence of maternal-reported episodes of diarrhoea, fever, cough, skin infection, eye infection and nasal discharge. However, there was a decreased incidence of vomiting in the iron compared to the placebo group (incidence difference: 0.37 episodes per 100 child-days of observation; 95%CI: 0·08 to 0·67) **(Table 4)**.

**Table 3.** Effect of iron supplementation on adverse events.

|  | Iron | Placebo |
| --- | --- | --- |
| **Serious adverse events** |  |  |
| Bronchiolitis | 1 | 1 |
| Constipation | 0 | 1 |
| Lower respiratory tract infection | 1 | 0 |
| Pneumonia | 1 | 0 |
| Sepsis | 1 | 0 |
| Severe malnutrition | 0 | 2 |
| Upper respiratory tract infection | 1 | 1 |
| **Adverse events** |  |  |
| Conjunctivitis | 1/1 | 4/4 |
| Fever due to immunisation | 18/16 | 25/17 |
| Multiple abscesses | 1/1 | 0 |
| Skin rashes | 6/6 | 5/5 |
| Upper respiratory tract infection | 27/24 | 16/15 |
| Hospitalisation for somatic illness | 1/1 | 2/2 |
| **Category of adverse events** |  |  |
| Grade 1 | 44 | 47 |
| Grade 2 | 7 | 3 |
| Grade 3 | 3 | 2 |

Values indicate the number of events or the number of events/number of children with at least one adverse event. Grade 1: Awareness of sign or symptom, but easily tolerated. Grade 2: Enough discomfort to cause interference with usual activity. Grade 3: Incapacitating with inability to do usual activity.

**Table 4.**
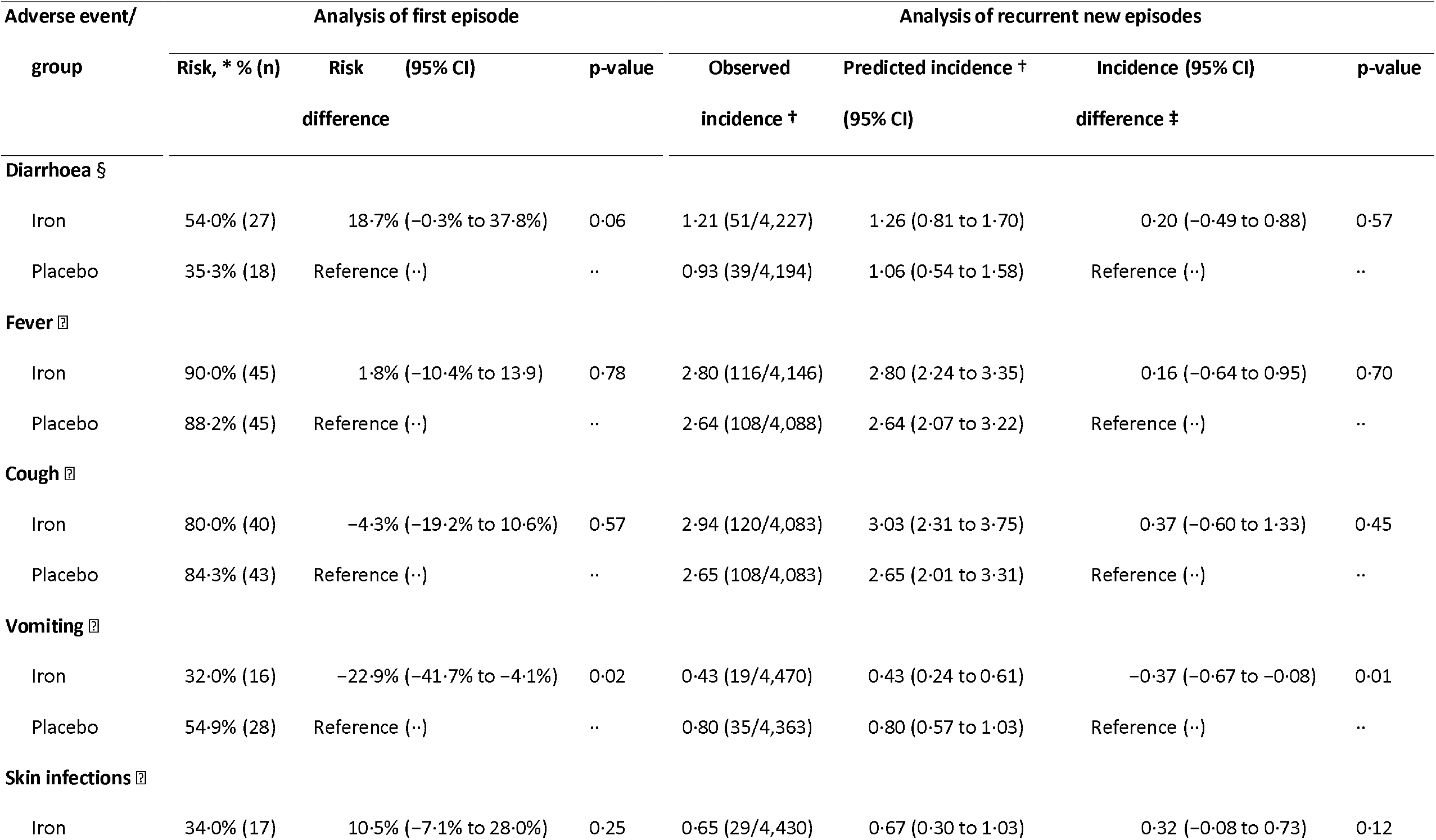
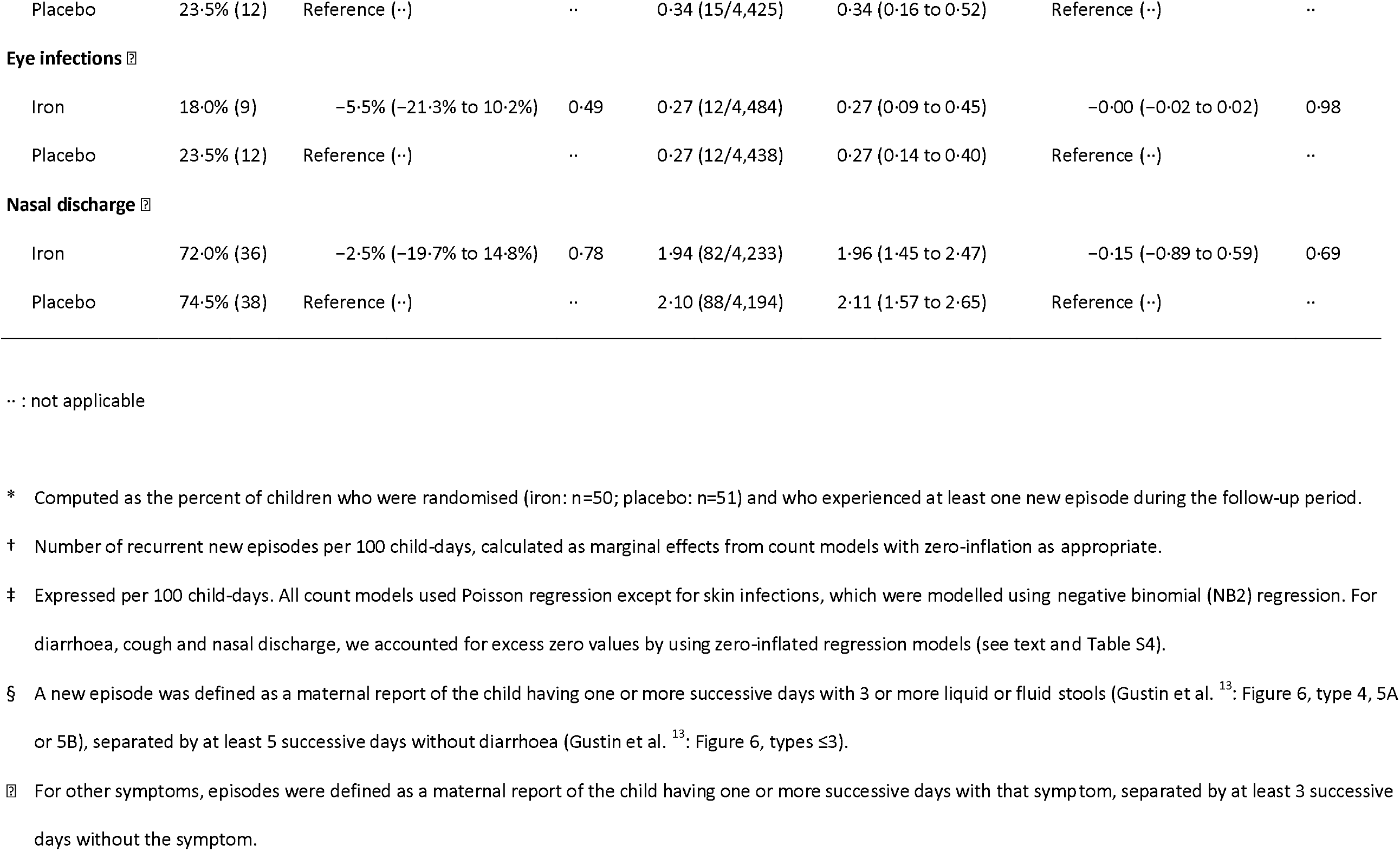
Group comparisons of maternally reported adverse events.

## Discussion

This proof-of-principle trial of early iron supplementation in fully breastfed Gambian children achieved substantial benefit across all markers of iron and haematological status. Although a full assessment of the safety of the intervention will require a much larger trial, it was reassuring that we detected no adverse effects on infections or growth. The target population was typical of many similar rural African settings where there is a high prevalence of moderate iron deficiency amongst pregnant mothers, and high proportions of infants born small and/or premature, all of which reduce an infant’s iron endowment at birth.

Iron is the nutrient most lacking in human breastmilk.^7^ and exclusively breastfed infants born with low iron reserves rapidly become very iron deficient.^6^ This might appear contrary to evolutionary theory which posits that breastfeeding has evolved as the optimal means for nourishing a young infant. However, breastfeeding evolved under very different dietary conditions to today’s predominantly cereal-based diets associated with the agrarian revolution which started only ∼12,000 years ago. For the vast majority of human evolution our hunter-gatherer predecessors consumed a mixed omnivorous diet with a high proportion of their food needs derived from animal sources and with high intakes of vitamin C (a promoter of iron absorption).^14^ Although direct evidence is lacking, it is assumed that these populations would have been replete in iron and that it was the ecological transition to cereal-based diets with low amounts of animal foods that caused a rise in deficiency diseases such as anaemia.^14^ Thus, when the composition of human milk was evolving, infants would likely have been born with a high iron endowment that would cover their needs for the first post-partum months and permit a very low milk iron content selected on the basis that it promotes a more favourable (lactobacillus and bifidobacterial dominant) gut microbiome. ^14^ In contrast modern breastfed infants become iron-deficient unless supplemented, and they will become progressively more iron deficient with increasing duration of exclusive breastfeeding.^6^

In high-income countries it is standard practice to recommend iron supplements soon after birth in infants at risk of iron deficiency,^7^ but in low-income settings, where the risk of deficiency is much higher, supplementation is viewed as potentially undermining the message that exclusive breastfeeding is the best way to feed an infant, and hence early iron supplementation has rarely been tested.^15^

To our knowledge, there are no previous trials of early iron supplementation for breastfed infants that have assessed the impact on serum iron and other markers of cellular iron supply. Most prior trials have started supplementation later (between 3 to 4 months of age) and have usually assessed haemoglobin and/or ferritin.^16^ Other trials have recruited children between 1-36 months^17^ and 2-36 months^18^ but have not analysed outcomes separately for the youngest children. A trial in Northeast India started iron supplementation very early post-natally.^19,20^ Supplementation with iron at 2 mg/kg (as ferrous ascorbate) from 36h after birth improved haemoglobin at 6 months from 97·0 to 103·7 g/L (P <0·0001) and ferritin from 78 to 134 ng/ml (P<0·001).^19^ In China, a 4-arm trial (total n=1,276) with iron offered in pregnancy and/or infancy (with ∼1mg/kg iron as iron protein succinylate oral solution from 6 weeks to 9 months) measured haemoglobin, ferritin, zinc protoporphyrin and sTfR at 9 months. ^21^ Iron supplementation reduced zinc protoporphyrin but was not found to improve other outcomes.^21^ In addition, in a trial in Honduras, infants who had been exclusively breastfed for 4 months were randomised to either continue being exclusively breastfed until 6 months or to receive iron-fortified foods in addition to breast milk from 4 to 6 months. The introduction of complementary foods in addition to breastfeeding resulted in an increase in iron intake, and in increased haemoglobin concentrations, haematocrit, and ferritin concentrations at 6 months of age. ^22^ In a similarly designed randomised trial in Iceland, the introduction of complementary foods in addition to breastfeeding resulted in an increase in serum ferritin concentrations, but there was no evidence that it improved haemoglobin concentration or other iron markers.^23^

Ideally, we would have assessed the change in our primary outcome, serum iron, over a 24h period, but this is not feasible in young infants. In light of this limitation the observed improvement in serum iron was particularly impressive given that it was assessed 24h after the last iron dose. The absorption of a bolus dose of ferrous sulphate occurs rapidly with a peak, in adults, at 3-4h post dose and with a subsequent decline towards baseline after 8h.^24^ In pilot work for a previous study we estimated that the peak serum iron level in young Gambian children was also at 3-4h post-dose. ^25^ We also demonstrated that at 6 m of age serum iron increased 5-fold (from 5.8 to 26.5 μmol/L) 4h after a 2 mg/kg dose of ferrous sulphate. Hence, in the current study, the aggregate benefit in supplying iron to iron-demanding tissues is likely to be much larger (estimated to be 3-4 fold higher) than indicated by the 24 h post-dose serum iron level. This is confirmed by Figure 2 which summarises all measures of iron status expressed as an effect size relative to the population standard deviation. The effects on the other longer-term metrics of iron status (of which reticulocyte haemoglobin (0.88 SD) and transferrin (−0.70 SD)) are particularly important) were larger than the 24-h post-dose serum iron (+0.53 SD).

A strength or our trial was the daily visits by fieldworkers who directly supervised consumption of the iron or placebo (and hence maintained a high compliance). The fieldworkers also asked mothers if their infant was sick and referred any worrying conditions to study nurses for assessment of adverse events, and onwards to a physician for any serious adverse events. There were no deaths, there were ten illnesses categorised as serious adverse events (5 intervention/5 control), all of which resolved, and there were 106 adverse events (54 intervention/52 control) all of which resolved. None of these conditions were deemed to be related to the intervention. In line with the low prevalence of *Plasmodium* infection in our study area,^8^ there were no incident cases of malaria in our study. Surprisingly we found weak evidence of a reduction in episodes of vomiting in the infants receiving iron.

Two prior trials in Honduras and Sweden reported that iron supplementation was associated with impaired growth. ^26^ We failed to find evidence of any such effect, and no other trials have replicated this finding.

The main limitation of our trial is that it is a proof-of-principle trial with a small sample size and will require replication in a much larger sample.

The benefits of early iron supplementation shown here are similar to those reported in a meta-analysis of 12 studies of delayed cord-clamping in low- and middle-income countries with ferritin, haemoglobin and MCV assessed between 2-12 months, ^27^ and in the 3 studies among these that made assessments at 6 months. ^28^ Although widely adopted as best practice it has proven difficult to implement delayed cord clamping in many settings and especially those, as in our study, with high proportions of home deliveries. It would be useful to test the combined effects of delayed clamping and early iron supplementation.

An improved haemoglobin at 6 months of age should enhance infants’ ability to learn through exploring their environment and improving attention, although these benefits have been surprisingly hard to demonstrate in studies of older children. ^29^ The health and developmental benefits of an enhanced systemic iron supply, as achieved here, might be greater for both cognitive and immune function because the critical neural and lymphoid tissues cannot access iron from liver ferritin or haemoglobin (the major body iron stores) and must utilise transferrin-bound iron. Based on our proof-of-principle success in the current trial further trials are warranted with adequate power to properly assess safety and to assess functional outcomes.

## Supporting information

Supplemental Annex

## Data Availability

All data produced in the present study are available upon reasonable request to the authors and are subject to review by the relevant Ethics committees.

https://figshare.com/account/projects/157242/articles/21878706

## Contributors

MB contributed to the study design, led the study as part of his PhD, oversaw all aspects of study implementation including data collection, data analysis and interpretation and writing of the original report and reviewing and editing. IS contributed to the study protocol design, project administration, reviewing and editing of the report and was supervised by SEM. CC conceived the study idea and acquisition of funding, design, and implementation, was overall principal investigator, contributed to the writing of the original report, reviewing and editing and supervised MB. HV contributed to the study design, data analysis and interpretation, writing of the original report, reviewing and editing and co-supervised MB. AMP contributed to the study idea and acquisition of funding, design, data visualisation, interpretation, writing of the original report, reviewing and editing. BS oversaw clinical care of the participants, contributed to data collection and reviewing and editing.

## Acknowledgments

This study was jointly funded by the UK Department for International Development (DFID), the Medical Research Council UK (MRC), the National Institute for Health Research and Care Research the Wellcome Trust and was awarded to the Medical Research Council Unit The Gambia at London School of Hygiene & Tropical Medicine (Grant Ref: MR/T003960/1). CC, BS and AMP were funded through MRCG@LSHTM. MB was funded through MRCG@LSHTM and Wageningen University and Research. HV was funded through Wageningen University and Research. IS and SEM are funded through a Wellcome Trust Senior Research Fellowship to SEM [220225]. MRCG@LSHTM is core-funded by MCA760-5QX00 from the MRC and the UK Department for International Development (DFID) under the MRC/DFID Concordat agreement. We sincerely thank Iron Babies Study Team at MRCG@LSHTM for their dedicated support. We also thank the participants and communities of Jarra Soma.

## Annexes

Supplementary Materials (including trial protocol) (Bah_etal_Supplementary Annex_FINAL)

Supplementary Figure S4 (Bah_etal_Supplementary_Fig S4)

