## Supplemental Annex for "Early iron supplementation of exclusively breastfed African infants: a proof-of-principle, placebo-controlled, randomised, double-blinded efficacy trial"

**Dealing with missing values**

In our multiple imputation model, we used all variables in the dataset except for censored variables (serum concentration of erythroferrone and C-reactive protein, both measured at baseline and at the end survey) and passive variables, derived from original variables (e.g., body mass index, derived from height and length; transferrin saturation derived from serum concentrations of iron and UIBC). We used 100 imputations for missing values in multiple variables iteratively by using chained equations. This was done separately within intervention groups, using predictive mean matching based on the nearest three neighbouring values.

**TABLE S1. Characteristics of the study population at baseline, per treatment arm (per protocol population)**

| Baseline variable | Iron | | Placebo | |
|---|---|---|---|---|
| n, randomised | 50 | | 51 | |
| Age, weeks | 50 | | 51 | |
| 6-6·99 weeks | | 58·0% (29) | | 49·0% (25) |
| 7-7·99 weeks | | 24·0% (12) | | 23·5% (12) |
| 8-8·99 weeks | | 6·0% (3) | | 15·7% (8) |
| 9+ weeks | | 12·0% (6) | | 11·8% (6) |
| Female | 50 | 52·0% (26) | 51 | 56·9% (29) |
| Number of children born to the mother | 50 | 3·9 (2·2) | 51 | 3·9 (2·1) |
| Serum iron, µmol/L | 50 | 12·6 (4·5) | 50 | 12·5 (6·9) |
| Serum ferritin, µg/L † | 50 | 283·2 (1·82) | 51 | 262·6 (1·76) |
| Serum soluble transferrin receptor, mg/L † | 50 | 2·81 (1·28) | 51 | 2·76 (1·30) |
| Serum sTfR/$\log_{10}$(ferritin) index † | 50 | 1·15 (1·34) | 51 | 1·15 (1·31) |
| Serum transferrin, g/L | 50 | 2·1 (0·4) | 51 | 2·2 (0·4) |
| UIBC, µmol/L | 50 | 37·0 (11·0) | 51 | 35·2 (11·0) |
| TSAT, % † | | 24·0 (1·64) | | 22·6 (1·99) |
| Haemoglobin, g/dL † | 50 | 10·8 (1.14) | 51 | 11·0 (1·12) |
| MCV, fL | 50 | 89.6 (5.3) | 51 | 89.2 (5.4) |
| MCH, pg | 50 | 29.5 (1.7) | 50 | 30.1 (8.4) |
| MCHC, g/dL † | 50 | 32·9 (1·4) | 51 | 32·6 (1·3) |
| Reticulocyte count, $\times 10^9$/L † | 41 | 0·07 (1·44) | 43 | 0·07 (1·49) |
| Reticulocyte count, % † | 41 | 1·83 (1·57) | 43 | 1·91 (1·54) |
| Immature reticulocyte fraction, % † | 41 | 17·2 (1·50) | 43 | 16·4 (1·52) |
| Reticulocyte haemoglobin, pg | 41 | 31·8 (1·78) | 43 | 31·3 (2·35) |
| Serum hepcidin, µg/L † | 50 | 20·3 (2·21) | 51 | 19·6 (2·36) |
| Serum erythropoietin, IU/L † | 50 | 12·7 (2·05) | 51 | 10·8 (1·85) |
| Serum erythroferrone, µg/L * | 50 | | 51 | |
| Below or equal to LOQ (≤ 0·16 µg/L) | | 66·0% (33) | | 74·5% (38) |
| 0.16-0.499 | | 22·0% (11) | | 3·9% (2) |
| 0.5-0.99 | | 10·0% (5) | | 5·9% (3) |
| 1.0+ | | 2·0% (1) | | 15·7% (8) |
| Serum $\alpha_1$-acid glycoprotein, g/L† | 50 | 0·66 (1·48) | 51 | 0·61 (1·80) |
| Serum C-reactive protein, mg/L * | 50 | | 51 | |
| Below or equal to LOQ (≤ 0·60 mg/L) | | 68·0% (34) | | 64·7% (33) |
| 0·60-4·99 mg/L | | 24·0% (12) | | 23·5% (12) |
| 5-14·99 mg/L | | 4·0% (2) | | 2·0% (1) |
| 15+ mg/L | | 4·0% (2) | | 9·8% (5) |
| Height-for-age z-score, SD | 49 | −0·59 (1·18) | 51 | −0·68 (1·59) |
| Weight-for-age z-score, SD | 49 | −0·71 (1·19) | 51 | −0·70 (1·25) |
| Weight-for-height z-score, SD | 49 | −0·22 (1·53) | 50 § | −0·11 (1·11) |

LOQ: limit of quantification; MCH: mean corpuscular haemoglobin; MCHC: mean corpuscular haemoglobin concentration; MCV: mean corpuscular volume; sTfR: soluble transferrin receptor; TSAT: transferrin saturation; UIBC: unsaturated iron-binding capacity

Values indicate n, mean (SD); n, geometric mean (geometric SD); * median (25-, and 75%-percentiles); † or prevalence % (n). The geometric SD is a dimensionless, multiplicative factor such that dividing or multiplication of the geometric mean by this ratio indicates a variation that is equivalent to subtraction or addition of one standard deviation on a log-transformed scale. ‡ Excluding one observation with an outlying value (86.8); § Data for 1 child is missing: this child had length of 41.2cm, which is below the minimum value (45cm) required to compute the weight-for-height z-score using the WHO international growth reference.

**TABLE S2. Effect of iron supplementation on selected outcomes (per protocol analysis)**

| Outcome variable | | Iron | | Placebo | Group difference, unadjusted (95% CI) | p-value | Group difference, * adjusted (95% CI) | p-value |
|---|---|---|---|---|---|---|---|---|
| Serum iron, µmol/L | 47 | 11·9 (4·6) | 50 | 9·4 (4·3) | 2·5 (0·7 to 4·3) | 0·0076 | 2·3 (0·6 to 4·1) | 0·0010 |
| Serum ferritin, µg/L † | 47 | 93·4 (1·90) | 50 | 44·3 (2·74) | 110·7% (50·2% to 195·5%) | <0·0005 | 117·2% (63·8% to 187·9%) | <0·0005 |
| Serum soluble transferrin receptor, mg/L † | 47 | 2·75 (1·10) | 50 | 2·96 (1·09) | −7·2% (−3·9% to −10·4%) | <0·0005 | −7·6% (−4·5% to −10·6%) | <0·0005 |
| Serum transferrin, g/L | 47 | 2·39 (0·38) | 50 | 2·72 (0·51) | −0·33 (−0·51 to −0·15) | <0·0005 | −0·37 (−0·52 to −0·22) | <0·0005 |
| UIBC, µmol/L | 47 | 47·0 (11·0) | 50 | 55·8 (11·0) | −8·7 (−13·0 to −4·5) | <0·0005 | −9·6 (−13·4 to −5·8) | <0·0005 |
| TSAT † | 47 | 22·7 (1·60) | 50 | 19·0 (1·87) | 19·9% (−4·0% to 49·7%) | 0·11 | 16·8% (−19·3% to 30·9%) | 0·052 |
| Serum sTfR/$\log_{10}$(ferritin) index † | 47 | 1·41 (1·20) | 50 | 1·79 (1·26) | −21·2% (−27·6% to −14·3%) | <0·0005 | −30·7% (−42·7% to −19·7%) | <0·0005 |
| Haemoglobin, g/dL | 47 | 11·2 (1·2) | 49 | 10·5 (0·9) | 0·6 (0·2 to 1·0) | 0·0061 | 0·6 (0·2 to 1·1) | 0·0092 |
| Anaemia (haemoglobin <11·0 g/dL) | 47 | 20 (42·6%) | 49 | 33 (67·3%) | −24·8% (−44·2% to −5·4%) | 0·012 | −23·3% (−41·0% to −5·5%) | 0·010 |
| MCV, fL | 47 | 77·7 (3·9) | 49 | 74·8 (4·3) | 2·9 (1·2 to 4·6) | 0·00086 | 2·7 (1·3 to 4·0) | <0·0005 |
| MCH, pg | 47 | 24·6 (1·4) | 49 | 23·3 (1·5) | 1·2 (0·7 to 1·8) | <0·0005 | 0·9 (0·5 to 1·4) | <0·0001 |
| MCHC, g/dL | 47 | 31·6 (0·8) | 49 | 31·1 (0·9) | 0·4 (0·1 to 0·8) | 0·012 | 0·34 (0·05 to 0·64) | 0·023 |
| Reticulocyte count, $\times 10^9$/L † | 47 | 0·06 (1·44) | 49 | 0·05 (1·43) | 19·6% (3·2% to 38·5%) | 0·018 | 20·3% (3·7% to 39·6%) | 0·015 |
| Reticulocyte count, % † | 47 | 1·35 (1·46) | 49 | 1·13 (1·45) | 19·4% (2·6% to 38·9%) | 0·022 | 19·8% (2·9% to 39·4%) | 0·021 |
| Immature reticulocyte fraction, % † | 47 | 9·53 (1·61) | 49 | 10·47 (1·45) | −9·0% (−23·5% to 8·4%) | 0·29 | −6·2% (−22·8% to 13·8) | 0·51 |
| Reticulocyte haemoglobin, pg | 47 | 30·2 (2·5) | 49 | 27·4 (3·1) | 2·8 (1·7 to 3·9) | <0·0005 | 2·65 (1·33 to 4·00) | <0·0005 |
| Serum hepcidin, µg/L † | 47 | 13·43 (2·11) | 50 | 7·50 (3·20) | 79·0% (20·9% to 164·8%) | 0·0040 | 72·6% (14·9% to 159·4%) | 0·0092 |
| Serum erythropoietin, IU/L † | 47 | 8·48 (2·04) | 50 | 10·58 (1·71) | −19·8% (−37·9% to 3·5%) | 0·089 | −20·3% (−37·5% to 1·7%) | 0·068 |
| Serum erythroferrone, µg/L * § | 47 | 0·16 (0·16, 0·58) | 50 | 0.42 (0.16, 1.26) | NA | 0.0052 | NA | NA |
| Serum C-reactive protein, mg/L * § | 47 | 1·3 (0·7, 4·2) | 50 | 1·4 (0·6, 4·6) | NA | 0.93 | NA | NA |
| Serum $\alpha_1$-acid glycoprotein, g/L † | 47 | 0·91 (1·48) | 50 | 1·01 (1·51) | −9·6% (−23·1% to 6·4%) | 0·22 | −8·8% (−22·5% to 7·2%) | 0·26 |
| Height-for-age z-score, SD | 47 | −0·81 (1·21) | 49 | −0·69 (1.16) | −0·12 (−0·60 to 0·36) | 0.62 | −0·14 (−0·20 to 0·49) | 0.42 |
| Weight-for-age z-score, SD | 47 | −0·64 (1·25) | 49 | −0·71 (1·12) | 0·07 (−0·41 to 0·55) | 0.78 | 0·12 (−0·17 to 0·42) | 0.42 |
| Height-for-weight z-score, SD | 47 | −0·07 (1·07) | 49 | −0·25 (1·07) | 0·18 (−0·26 to 0·61) | 0.42 | 0·19 (−0·23 to 0·61) | 0.38 |

Values for group descriptors indicate n, mean (SD); n, geometric mean (geometric SD); † median (25- and 75-percentiles); ‡ or prevalence (n) %. The geometric SD is a dimensionless, multiplicative factor such that dividing or multiplication of the geometric mean by this ratio indicates a variation that is equivalent to subtraction or addition of one standard deviation on a log-transformed scale. Intervention effects are reported as absolute differences, relative differences † or prevalence ratios.

MCH: mean corpuscular haemoglobin; MCHC: mean corpuscular haemoglobin concentration; MCV: mean corpuscular volume; NA: not applicable; sTfR: soluble transferrin receptor; UIBC: unsaturated iron-binding capacity

* Effect estimates adjusted for the outcome variable measured at baseline, sex (binary variable), age (continuous variable), and haemoglobin concentration and serum concentrations of ferritin and soluble transferrin receptor at baseline (all continuous variables); except for anthropometric outcomes that were adjusted for the outcome variable measured at baseline, sex (binary variable) and age (continuous variable). Continuous covariates were modelled as fractional polynomials as appropriate to account for non-linear relationships with the outcome (see text). ‡ Variables were left-censored at 0·16 µg/L (erythroferrone) and 0·60 µg/L (C-reactive protein) due to values being below the limit of quantification. In the iron group, most erythroferrone values (26/47) were left-censored, so that the median equals the 25-percentile. Because log-transformation failed to normalise the distribution of residuals within groups, we did not conduct parametric analyses but only report p-values obtained by two-sample Wilcoxon rank-sum (Mann–Whitney) tests.

**TABLE S4. Group comparisons of maternally reported adverse events – zero-inflated models ***

| Adverse event/ group | IRR (95% CI) | p-value | OR (95% CI) | p-value |
|---|---|---|---|---|
| **Diarrhoea** | | | | |
| Iron | 0·71 † (0·40 to 1·26) | 0·24 | 3·05 † (1·03 to 9·05) | 0·04 |
| Placebo | Reference – | – | Reference – | – |
| **Cough** | | | | |
| Iron | 1·28 (0·90 to 1·82) | 0·17 | 0·30 (0·03 to 3·36) | 0·33 |
| Placebo | Reference – | – | Reference – | – |
| **Nasal discharge** | | | | |
| Iron | 0·98 (0·66 to 1·45) | 0·90 | 0·71 (0·12 to 4·12) | 0·71 |
| Placebo | Reference – | – | Reference – | – |

– : not applicable; IRR: incidence rate ratio; OR: odds ratio

* See also footnotes.

† Interpretation, for example: among a latent class of children susceptible to diarrhoea, iron supplementation is associated with a reduction in the incidence of recurrent new episodes by 29% (= 0.71 – 1) × 100). Iron supplementation is also associated with an increase by 205% (= (3.05 – 1) × 100) in the odds of being in a latent group of children that is not susceptible to diarrhoea compared to being in the group that is susceptible to diarrhoea. The incidence difference (reported in Table S3) is a marginal effect that is calculated by combining of the effects among these latent classes.

**FIGURE S1. Adherence to intervention, by group**

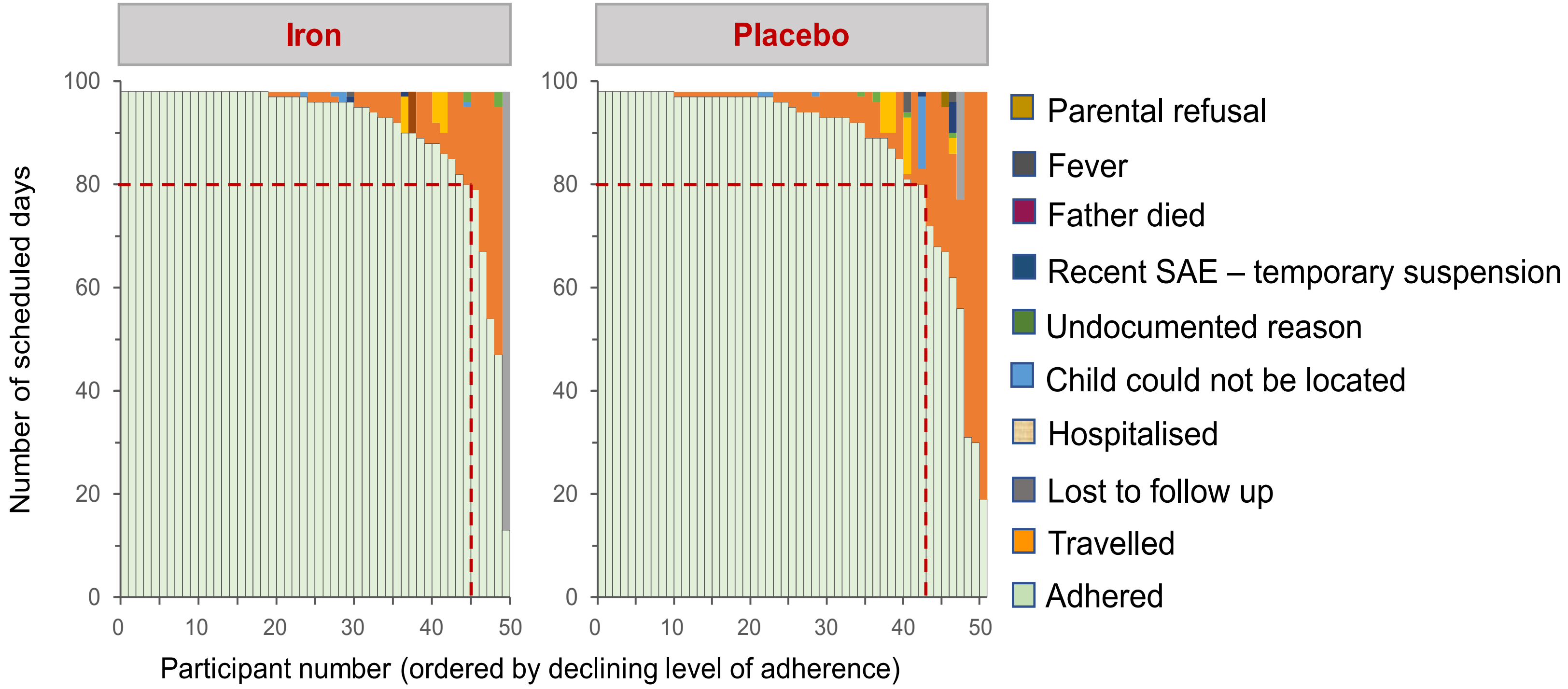

SAE: severe adverse event.

The Y-axes in these bar graphs denote the number of days that supplementation was scheduled (maximum: 98days). The X-axes show group participants in order of declining adherence, which was indicated by the number of days that supplements were actually ingested ('Adhered'; light green bars). Thus, the step-function indicated by the top of these bars indicates the inverse cumulative adherence for each intervention group. For example, 90% (= 45 × 100/50 and 84% (= 43 × 100/51) of participants in groups receiving iron and placebo consumed at least 80 supplements (indicated by red dashed lines), which, expressed as a percentage of the total number of scheduled supplements per person, corresponds to an adherence of 81·6% (= 80/98 supplements). On the other hand, when calculated as the area under the curve, expressed as a percentage of the total number of scheduled supplements (= number of children × 98 supplements), adherence was 92·3% and 89·1% for groups receiving iron and placebo, respectively. Reasons for non-compliance are indicated by different colours.

**Computation and group comparisons of frequencies of episodes of maternally-reported adverse symptoms**

Kaplan-Meier analysis and Cox proportional hazard models are commonly used to estimate time to first event in longitudinal studies. However, because trial participants may experience multiple events, particularly when due to infectious diseases, such models may lead to biased estimates of the total burden of events. In this section, we explain how we handled the analyses of maternally reported recurrent adverse events. To illustrate our approach, we start with the analysis of fever.

We report the frequency of episodes by group both as risks (the percent of enrolled children who experienced at least one new episode during the follow-up period) and as incidence rates (the number of new episodes per 100 child-days of observation). We compared groups only when there were sufficient numbers of episodes to allow meaningful analysis. We used Kaplan-Meier graphs with corresponding p-values from log-rank test to compare groups with regards to time to occurrence of the first new episode. To analyse multiple episodes per child, we compared groups using count regression models, with zero-inflation as appropriate to deal with count distributions with excess zero values. Results are reported as marginal effects.

A new episode of maternal-reported fever was defined as one of more successive days of reported fever, which was separated from a previous episode by an arbitrarily selected period of at least 3 successive days without reported fever. When an intermittent period of ≤3 fever-free days occurred, then these days were included in the calculation of the duration of the episode (e.g., **Figure S2**, episode 2: day 15). During an episode, children were considered at risk only on the first day, and not for the remainder of the episode, so that these additional days were subtracted from the observation time (Figure S2: 5 days for episode 2, and 2 days for episode 3, for a total of 7 days). In addition, children were not considered at risk for 3 days after the episode had finished, because any fever occurring during this time would be considered to be part of the previous episode. Thus, in the example provided in Figure S2, the observation time was 11 days (= 30 scheduled days – 7 days – 9 days [3 days subsequent to each of the episodes 1-3] – 3 days [missing values]).

#### FIGURE S2. Definition of episodes and analysis of episode frequency: hypothetical example of maternal-reported fever, with a 30-day scheduled follow-up time for a single child

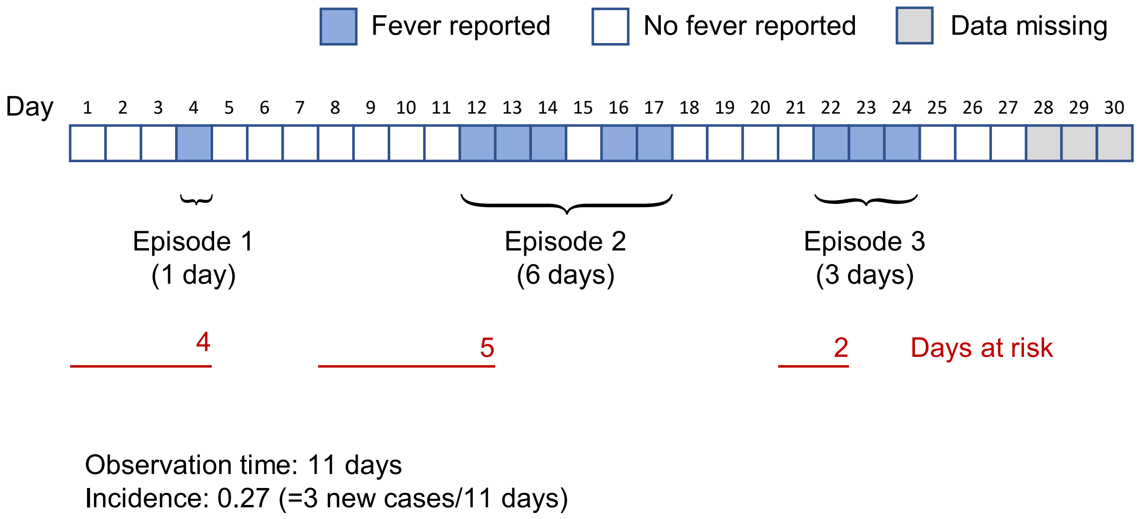

Other adverse outcomes were analysed similarly except that, in the analysis of diarrhoea, we considered new episodes as the passage of 3 or more loose or liquid stools per day (Type 4, 5A or 5A;[1] : Figure 6; [2] which were separated by an arbitrarily selected time period of at least 5 days.

We used Poisson and negative binomial regression models to estimate the incidence of maternally reported episodes by intervention group, using counts for individual children as obtained by the procedures as outlined above and accounting for differences between children in observation time that could occur due to missing values and censoring following each episode (see preceding sections). In these analyses, we estimated a robust variance using a ‘sandwich' estimator to account for events being possible related to each other. We used zero-inflation to account for excess zero values due to individuals not being at risk. Models were compared using Akaike’s and Bayesian information criteria and, when nested, by likelihood ratio test. Model selection was furthermore based on a comparison of predicted and observed counts. No adjustments were made for baseline variables. Using these models, we predicted the incidence using Stata’s ‘margins’ command, conditioned to a 100-day intervention period, to allow comparison with the observed incidence.

To account for multiple new episodes per child, we compared groups using count regression models, with zero-inflation as appropriate to deal with count distributions with excess zero values (‘peak at zero’), i.e., the number of zero episodes exceeds the number that may be expected on the basis of a Poisson or negative binomial distribution. As applied to the present study, zero-inflated models assume there are two latent groups of children: a) one group consists of children susceptible to episodes of diarrhoea but who also have a nonzero probability of having zero episodes; b) another group of children who experience zero episodes because they are not susceptible (e.g., because they can tolerate supplementation; they happen not to be exposed to infection during the intervention period due to environmental or behavioural characteristics; infections are not serious enough to produce symptoms; acquired immunity; or because they receive medication to prevent or suppress symptoms). Thus, the zero counts that originate from the first group is ‘inflated’ by the zero count from the second group. We used zero-inflated count regression models that comprised a Poisson or negative binomial regression component to model the count process, and a binary logistic regression component to model the excess zeroes.

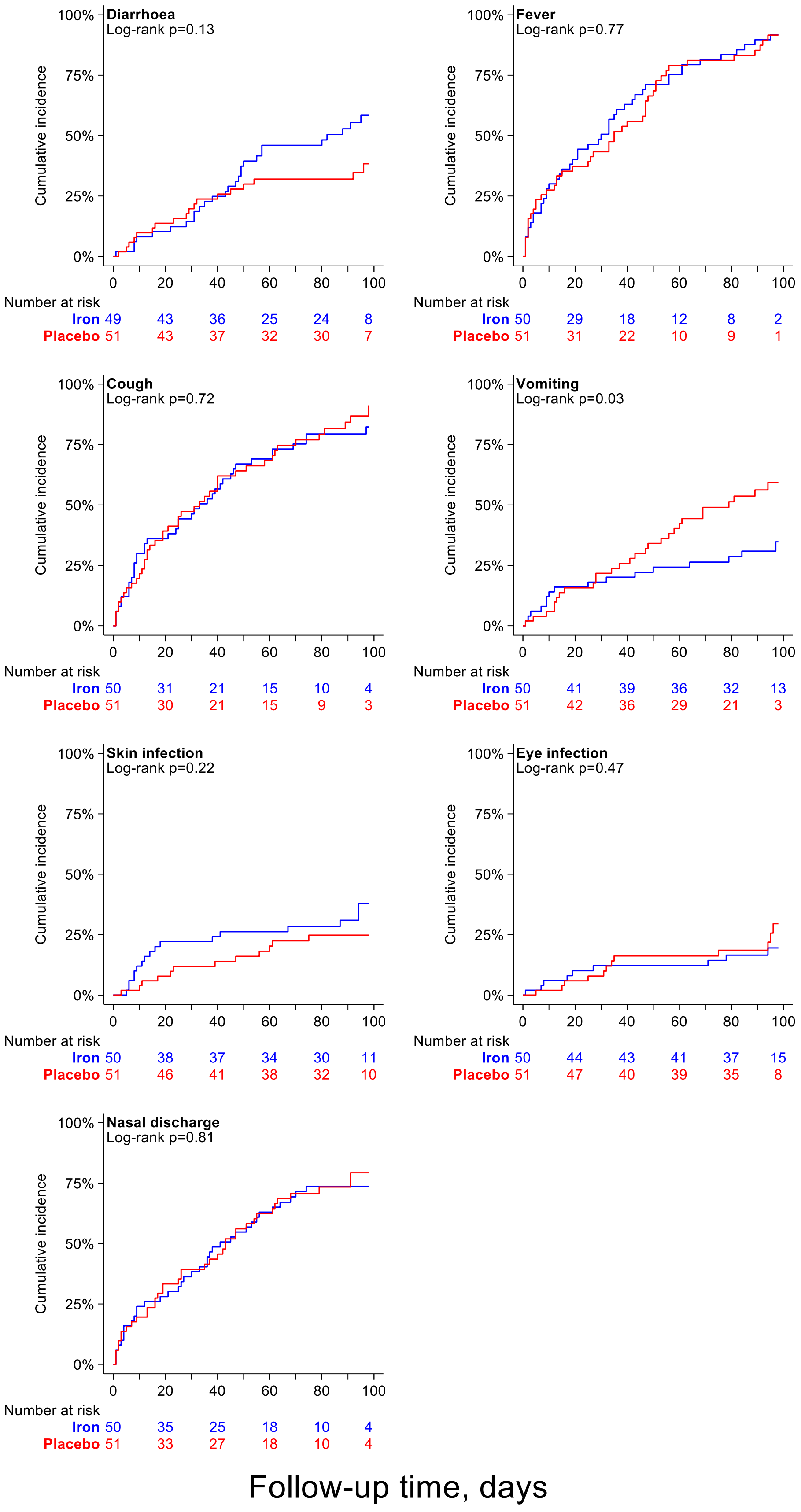

**FIGURE S3. Group differences in time to first episode for various adverse effects of interest, Kaplan-Meier plots**

**FIGURE S4. Maternal reports of child illnesses (see Excel sheet labelled Bah_etal_Supplementary_Fig S4)**

**CLINICAL TRIAL PROTOCOL**

---

**Title**
Enhancing brain development by early iron supplementation of African infants: An enabling pilot study

---

**Protocol No:** 2.0

**SCC No:** 19092
**Alias** Iron Babies Pilot Supplementation Trial
**Other Number(s)**
**Protocol Version – Date:** 2.0 30 June 2021

| | |
|---|---|
| **Sponsor:** | Medical Research Council Unit The Gambia at London School of Hygiene & Medicine<br>PO Box 273 Banjul,<br>The Gambia, West Africa |

**Principal Investigator:** Dr Carla Cerami

**Signature page**

The clinical trial will be carried out in accordance with the protocol, the ICH Harmonised Tripartite Guideline for Good Clinical Practice, <<*insert other regulations if applicable*>>, and in accordance to local legal and regulatory requirements.

| Principal Investigator: | Signature: | Date: 30.06.2021 |
|---|---|---|
| *Name: Carla Cerami* | | |

| Sponsor's representative: | Signature: | Date: |
|---|---|---|
| Elizabeth Batchilly | | |
| *Name* | | |

**Table of contents**

Page
Signature page 12
Protocol amendment(s) **Error! Bookmark not defined.**
Key roles 16
List of abbreviations 18
Protocol summary 19
1 Background information and rationale 21
1.1 Background information 21
1.2 Rationale 21
1.3 Potential risks and benefits **Error! Bookmark not defined.**
2 Study objectives 22
2.1 Study endpoints 22
3 Study design **Error! Bookmark not defined.**
3.1 Type of study and design 22
3.2 Randomisation and blinding procedures **Error! Bookmark not defined.**
3.2.1 Randomisation 22
3.2.2 Blinding 23
3.3 Sub-studies 23
3.4 Investigational products **Error! Bookmark not defined.**
3.4.1 Description of products 23
3.4.2 Formulation, packaging and labelling 23
3.4.3 Product storage and stability 23
3.4.4 Dosage, preparation and administration of investigational products 23
3.4.5 Concomitant medications/treatments 24
4 Selection and withdrawal of participants **Error! Bookmark not defined.**
4.1 Selection of participants 24
4.2 Eligibility of participants 24
4.2.1 Inclusion criteria 24
4.2.2 Exclusion criteria 24
4.3 Withdrawal of participants **Error! Bookmark not defined.**
5 Study procedures and evaluations **Error! Bookmark not defined.**
5.1 Study schedule 25
5.1.1 Screening **Error! Bookmark not defined.**
5.1.2 Enrollment (Baseline) 25
5.1.3 Follow-up 25
5.1.4 Final study visit 25
5.1.5 Early termination visit 25
5.2 Study evaluations 25
5.2.1 Clinical evaluations 25
5.2.2 Laboratory evaluations 25
6 Safety considerations 26
6.1 Methods and timing for assessing, recording, and analysing safety parameters 27
6.1.1 Adverse events 27
6.1.2 Reactogenicity 27
6.1.3 Serious adverse events (SAEs) 27
6.2 Reporting procedures 28
6.3 Safety oversight 28
7 Discontinuation criteria 28
7.1 Participant's premature termination 28
7.2 Study discontinuation 29
8 Statistical considerations 29
9 Data handling and record keeping 30
9.1 Data management and processing 30

12 Financing and insurance 33
13 Publication policy 33
14 References 34
Supplements, appendices and other documents 35
Appendix: Schedule of events **Error! Bookmark not defined.**

**Key roles**

For questions regarding this protocol, contact Isabella Stelle at Kings College London, St Thomas' Hospital, Westminster Bridge, London, SE1 7EH.. +44(0)7704906558 or Carla Cerami at MRCG

| | |
|---|---|
| **Author(s):** | Ms Isabella Stelle, Dr Carla Cerami |
| **Sponsor's representative:** | Elizabeth Batchilly,<br>Quality & Governance Manager<br>London School of Hygiene & Tropical Medicine<br>Keppel St, London, WC1E 7HT<br>UK |
| **Chief Investigator:** | NA |
| **Principal Investigator(s):** | Dr Carla Cerami<br>Research Group Lead: Iron, Infection and Anemia<br>Nutrition Theme<br>MRC Unit The Gambia @ LSHTM<br>PO Box 273, Banjul<br>The Gambia<br> |
| **Sub-Investigator(s):** | Prof Andrew M Prentice<br>Nutrition Theme Leader<br>MRC Unit The Gambia @ LSHTM<br>PO Box 273, Banjul<br>The Gambia<br><br><br>Dr Sophie Moore<br>Research Group Lead: Early, Growth and Development<br>MRC Unit The Gambia @ LSHTM<br>PO Box 273, Banjul<br>The Gambia<br>And<br>Reader<br>Department of Women's and Children's Health<br>Kings College London<br>North Wing St Thomas<br>London, United Kingdom<br><br>Ms. Isabella Stelle, PhD Student<br>Department of Women's and Children's Health<br>Kings College London<br>North Wing St Thomas<br>London, United Kingdom<br><br><br>Mamadou Bah, PhD Candidate<br>Nutrition Theme<br>MRC Unit The Gambia @ LSHTM<br>PO Box 273, Banjul<br>The Gambia<br> |

**Sponsor's Medical Expert:** MRC Unit The Gambia
PO Box 273, Banjul
The Gambia

**Trial monitor(s):** Aja Nyarra Sanyang
MRC Unit The Gambia
PO Box 273, Banjul
The Gambia


**Local Safety Monitor:** Dr Patrick Nshe
MRC Unit The Gambia
PO Box 273, Banjul
The Gambia


**Chair of DMC/DMSB:** Dr Seth Adu-Afarwuah
University of Ghana
Department of Nutrition and Food Sciences


**Statistician:** Dr Hans Verhoef
Waginengen University
The Netherlands


**External Adviser:** Dr Kamija Phiri
Professor of Clinical Epidemiology
University of Malawi


Dr Helen Nabwera
Liverpool School of Medicine
Liverpool, UK


Ms Miriam Wamaitha Wathuo
MRC Unit the Gambia at LSHTM
The Gambia


**Clinical Laboratory/ies:** MRC Unit the Gambia at LSHTM
**Other institutions/ Collaborators:** Aminata Trawally
Nutrition Program Officer
Gambian National Nutrition Agency
Bakau, The Gambia


**Local Ethics Committee** Gambia Government/MRC Joint Ethics Committee,
c/o MRC Unit, The Gambia
PO Box 273, Banjul, The Gambia, West Africa

**List of abbreviations**

| | |
|---|---|
| AE | Adverse Event |
| CRF | Case Report Form |
| DMC | Data Monitoring Committee |
| DSMB | Data & Safety Monitoring Board |
| ENID | Early Nutrition and Immune Development (Trial) |
| GCP | Good Clinical Practice |
| DM | Database Manager |
| FW | Field Worker |
| Hb | Hemoglobin |
| ICH | International Conference on Harmonization |
| ID | Iron Deficiency |
| IDA | Iron Deficiency Anaemia |
| IEC | Independent Ethics Committee |
| IP | Investigational Product |
| LBW | Low birthweight |
| LSM | Local Safety Monitor |
| MRC at LSHTM | Medical Research Council Unit The Gambia at London School of Hygiene & Tropical Medicine |
| MRCG | Medical Research Council Unit The Gambia |
| PI | Principal Investigator |
| SAE | Serious Adverse Event |
| SAR | Serious Adverse Reactoin |
| SOP | Standard Operating Procedure |
| sTfR | Soluble Transferrin Receptor |
| SSP | Study Specific Procedure |
| TSC | Trial Steering Committe |

#### Protocol summary

| | |
|---|---|
| **Title:** | Enhancing brain development by early iron supplementation of African infants: An enabling pilot study |
| **Alias :** | Iron Babies Pilot Supplementation Trial |
| **Phase**: | Pilot |
| **Population:** | Rural Gambian infants in Jarra West |
| **Number of participants:** | 100 |
| **Number of Sites:** | 6-25 villages |
| **Location of Sites (including satellite sites):** | Medical Research Council Unit The Gambia (MRCG), Keneba Field Station, The Gambia<br>(Jarra Soma.) |
| **Trial Duration:**<br>**- Clinical Phase:**<br>**- Whole trial:** | 14 weeks clinical /participant phase and 12 months total<br>18 months |
| **Duration for Participants:** | 14 weeks |
| **Description of Investigational Products:** | Daily Drops with and without iron<br>Test: 7.5mg/day of elemental iron as ferrous sulphate<br>Control: flavoured syrup |
| **Objectives:** | **Primary objective:**<br>Will provision of a daily iron supplement as paediatric drops for 98 days starting at 6-10wks of age reverse the decline in serum iron?<br>**Secondary objectives:**<br>1. Can we introduce supplementary iron without undermining the duration of exclusive breast feeding?<br>2. Would iron supplementation at this age cause diarrhea (potentially by altering the infant gut microbiome)?<br>3. Would iron supplementation cause an increase in fecal iron losses? |
| **Endpoints:** | **Primary:** Serum iron after 98 days of iron supplementation. Serum iron will be measured in venous blood collected at trial enrolment and 14 weeks after initiation of iron supplementation.<br><br>**Secondary:**<br>**Related to the primary objective:**<br>Proportion of infants with anaemia (Hb < 11 g/dL) at study day 98.<br>Proportion of iron deficiency (sTfR/logFerritin ratio <2.0; hepcidin < 5.5 ng/L) at day 98.<br>Proportion of infants that are iron deficiency anaemia (Hb < 11 g/dL & sTfR/logFerritin ratio < 2.0 and ferritin < 12 ug/L or < 30 ug/L in the presence of inflammation) will be determine at day 98<br><br>**Related to secondary objective 1:**<br>Duration of breast feeding assessed using weekly questionnaires previously validated in ENID (Early Nutrition and Immune Development) trial [1]<br><br>**Related to secondary objective 2:**<br>Proportions of maternal-reported illnesses.<br>Proportion of adverse events (AEs). |

Proportion of serious adverse events (SAEs).
Proportion with raised inflammatory markers (CRP/AGP).
Proportion of raised recticulocytes counts

**Related to secondary objective 3:**
Fecal iron at baseline and endline stool sampling.
Gut pathogen analysis at baseline and endline stool sampling

**Description of Study Design:**

2 arm, double blind, placebo controlled, randomised trial, with 50 infants per arm.

Healthy infants 6-10wks of age will be randomised to receive daily supplementation for 98 days of either a) iron drops or b) placebo drops. Infants with significant illness or any clinical syndromes that would affect interpretation will be excluded. Low birthweight (LBW) infants and infants born prematurely will not be excluded. Venous blood samples will be collected at enrolment (age 6-10 weeks of age) and after 14 weeks (98 days) of iron/placebo supplementation.

Participants will be visited daily in their villages by Fieldworkers (FWs) to help administer the iron/placebo dose and will interview parents/guardians to complete a short health questionnaire. The iron will be dosed at 75% of WHO guideline dose for a 6week old infant (ie 1.5mg/kg/day equivalent to 7.5mg/day of elemental iron). Weekly, a more detailed morbidity and breastfeeding questionnaires will be administered. Infants will be weighed and measuredat endline and baseline, along with a faecal sample being taken.

During the daily visits, the FWs will record any adverse events (AEs) and ensure the safety of participants. If an infantis found unwell or if the mother/guardian reports that the infant is unwell, the study nurse will check on the infant and decide on treatment/referral to the nearest health centre.

### 1 Background information and rationale

#### 1.1 Background information

Healthy full-term babies are born with an endowment of iron accumulated in the fetal liver and stored as ferritin[2]. Human breast milk contains very low levels of iron so the hepatic store of iron accrued in utero is used as a buffer to meet the needs of new tissue formation. Most of the body iron (~60-70% in infants) is contained within haemoglobin (Hb) and erythropoiesis is prioritised over other organs including the brain if iron supply is restricted[3]; thus, iron deficiency (ID) occurs well ahead of anaemia. This risk is greatest in infancy when the iron needs of growth compete with those of erythropoiesis. Studies in infant humans [4][5], monkeys [6], lambs [7] and rats [8] demonstrate that brain iron is reduced prior to the appearance of anaemia.
During the first 3 years of life there is rapid myelination especially of the frontal cortex and basal ganglia (motor control) [9]. Infants with ID can have symptoms that are consistent with impaired hippocampal function, reduced myelination, and altered temperament and dopamine metabolism. Iron-deficient infants can present with decreased attention and memory [10] with deficits in visual and auditory systems and altered temperament and social and emotional behaviours [11].
A large body of evidence both from humans and animal models indicates that ID in early life can permanently alter the brain and nervous system [12]. Pre-clinical models have clearly demonstrated that these effects are due to brain tissue ID and not to anaemia. Therefore, treatment of ID after the onset of anaemia may not have any effect on the neurological deficits.

References of literature and data are listed in Section 14.

#### 1.2 Rationale

We have recently conducted a pooled analysis of iron and haematological status in 317 rural Gambian infants from two longitudinal birth cohorts [2]. We additionally studied the role of inflammation and changes in the hormone hepcidin (the master regulator of iron metabolism). Cord ferritin levels were high; confirming that babies were born with a reasonable endowment of iron despite being born to iron deficient mothers. Following birth there was a very rapid deterioration of Hb and ferritin status, especially in the fastest growing babies [2]. Most notably, the children showed extremely low levels of serum iron starting in early infancy and continuing beyond the first year of life (see the blue and red diamonds in Figure 1). By 5 months of age, close to 95% of the infants had serum iron levels below the 7.9µmol/l lower limit of the clinical reference range (Fig 2).
Serum/plasma iron (bound to transferrin) is the source of iron for growing tissues including neuronal cells and hence such very low levels of circulating iron suggest that the growing infant brain must be short of iron. Changes in plasma levels of soluble transferrin receptor (sTfR) reinforce this conclusion. sTfR rose sharply from 2 months of age in the Gambian infants indicating that target cells were upregulating their receptors to sequester iron in the face of an impoverished supply. These data strongly suggest that the infants' brain development is likely impaired by an inadequate supply of iron.
Recent meta-analyses and reviews have concluded that, although there is evidence suggesting a benefit of iron supplementation on cognitive development in young children, the evidence base is weak due to an absence of high quality randomised trials [13]. Furthermore, a large portion of published and registered on-going trials have initiated supplements after 6 months of age; by which time children living in poor communities in rural Africa and elsewhere are already likely to be iron deficient (see Fig 2).

Iron supplementation of breast fed babies is routine in the United States. In term infants, the American Academy of Pediatrics recommends supplementation with elemental iron 1mg/kg/day (maximum 15 mg) in term infants and 2-4mg/kg/day in preterm infants. In the UK, the UK Department of Health and the Royal College of Paediatrics, do not recommend iron supplementation in term babies, but the current recommendation for stable/growing preterm infants is to give iron at 2mg/kg of body weight up to a maximum of 15mg/day until one year of age.
Our goal in this study is therefore to conduct a pilot randomized trial by introducing iron supplements much earlier in infancy than has previously been attempted in a low-income setting with an aim to improve serum iron concentration that may positively impact on the neurological development of infants in the long term.

#### 1.3 Potential risks and benefits

The potential risks to human subjects and known benefits, if any, are summarised in Section "Human Subject Protection".

#### 2 Study objectives

**Primary Objective:**

Will provision of a daily iron supplement as paediatric drops starting at 6-10 wks of age reverse the decline in serum iron?

**Secondary Objectives:**

1. Can we introduce supplementary iron without undermining the duration of exclusive breast feeding?
2. Would iron supplementation at this age cause diarrhea (potentially by altering the infant gut microbiome)?
3. Would iron supplementation cause an increase in fecal iron losses?

##### 2.1 Study endpoints

**Primary:** Serum iron concentration after 98 days of iron supplementation. Serum iron will be measured in venous blood collected at trial enrolment and 14 weeks after initiation of iron supplementation.

**Secondary:**

**Related to the primary objective:**

Proportion of infants with anaemia (Hb < 11 g/dL) at study day 98.
Proportion of iron deficiency (sTfR/logFerritin ratio <2.0 hepcidin < 5.5 ng/L) at day 98.
The proportion of infants that are iron deficiency anaemia (Hb < 11 g/dL & sTfR/logFerritin ratio < 2.0 and ferritin < 12 ug/L or < 30 ug/L in the presence of inflammation) will be determine at day 98.

**Related to secondary objective 1:**

Duration of breast feeding assessed using weekly questionnaires previously validated in ENID (Early Nutrition and Immune Development) trial [1].

**Related to secondary objective 2:**

Proportions of maternal-reported illnesses.
Proportion of adverse events (AEs).
Proportion of serious adverse events (SAEs).
Proportion with raised inflammatory markers (CRP/AGP).
Proportion of raised reticulocytes count markers

**Related to secondary objective 3:**

Fecal iron at baseline and endline stool sampling.
Gut pathogen measurement at baseline and endline stool sampling

#### 3 Study design

##### 3.1 Type of study and design

This is going to be a 2-arm parallel double-blind randomised control trial with 50 infants per arm. Healthy 6- to 10-week-old infants will be randomised to 14 weeks of daily supplementation with either: a) iron drops (at 7.5 mg/day of elemental iron); or b) placebo drops. Infants with significant illness or any clinical syndromes that would affect interpretation will be excluded. Low birthweight (LBW) infants will not be excluded. Anthropometry, and venous blood and faecal samples will be collected at enrolment (age 6 to 10 weeks) and after 14 weeks (98 days) of iron supplementation. Eligible infants in the participating communities will be enrolled into the study once a signed inform consent are obtained from parents/guardians.

Blood samples will be analysed at MRC Unit The Gambia laboratories.
Fecal samples will be analyzed at the MRC Unit the Gambia or shipped overseas.

##### 3.2 Randomisation and blinding procedures

###### 3.2.1 Randomisation

Permuted block randomisation with a block size of 6 (e.g. 1-6) will be use to randomize participants to either intervention or control to prevent imbalance sample size between arms. This will be done in RedCap by the database manager (DM) with support from the statistician. To prevent potential biased during randomization, only the DM will have access to the randomization CRF module which is password protected. Each participants' study ID will be assigned to a unique intervention code that is only known by a dedicated person who is independent from the study. This will prevent the unblinding of the entire study should there be a request for unblinding from the Data Safety Monitoring Board (DSMB) due to AEs. A list of IDs will be generated in advance by the MRC data team with the oversight of the DM before the consenting visits. The study ID will comprise of a three-letter study code (Iro), a 3-digit unique number (e.g. 001-140) and a check letter (e.g. A-Z). Every consented infantl will be assigned a study ID e.g. Iro001X). After screening and obtaining signed inform consent from parents/guardians, participants will be invited to a dedicated location for their baseline sample collection. At this point the study nurse will confirm eligibility. At the end of the baseline day the DM/sub-investigators will assign one of the two blinded trial arms ('iron', 'placebo') to each eligible participant's study ID by running the randomization module in REDCap cap. The sub-investigator will be responsible for organising the FWs to ensure the implementation of the trial follows the treatment allocation assigned by the study IDs.

##### 3.2.2 Blinding

This will be a two-arm, randomised, placebo-controlled double-blind study. There will be 6 intervention codes (e.g. 1-6) and three will be assigned to the intervention arm, whereas the other three will be assign to the placebo arm, by the dedicated person. The pharmacist will re-bottle and label the investigational product (IP) according to this block randomization (unknown to the blinded study team). The randomization code will be kept in a locked box, only accessed if unblinding become necessary.

Infants will be randomised (1:1) to iron drops or placebo arm.

#### 3.3 Sub-studies

#### 3.4 Investigational products

##### 3.4.1 Description of products

One study arm will consume daily drops: 7.5mg/day of elemental iron as ferrous sulphate. A second study arm will be administered daily placebo drops in an identical bottle.

##### 3.4.2 Formulation, packaging and labelling

Daily Iron Drops

Formulation: Ferrous sulphate BP equivalent to 15mg iron per ml
Packaging: Type III Glass dropper bottle vial 15ml; 18mm white HDPE child resistant cap (ISO 8317) supplied with calibrated oral syringe 1ml
Labelling:
MRC Unit, The Gambia Study #19092
PI: Dr Carla Cerami, +220 787 5756
Expiry Date 03/2022
Intervention arm: (ie 1 –6)
0.5ml to be taken daily by the trial participant
*Trial medication for the use of Iron Babies trial participants only*

Daily Placebo Drops

Formulation: Sorbitol Solution 70% (in-active ingredient)
Packaging: Type III Glass dropper bottle vial 15ml; 18mm white HDPE child resistant cap (ISO 8317) supplied with calibrated oral syringe 1ml
Labelling: As above

##### 3.4.3 Product storage and stability

Storage of products will be at room temperature (15-30˚C), away from direct sunlight, at both the MRCG Keneba Field Station main store and Jarra Soma health clinic in a locked room. The IPs are stable when stored at this temperature. The procured products have a 12-month shelf-life after manufacture, and expiry dates will be carefully checked by the sub-investigator prior to study implementation.

##### 3.4.4 Dosage, preparation and administration of investigational products

The products are designed to be given as a daily dose based on average infant weight at a dose of 1.5mg/kg/day, and composition details are given in section 3.4.2. The dosage will remain at 7.5mg/day of elemental iron, the

equivalent does for a 6-week old female infant. The supplements will be given to the participants by the FWs using individual marked droppers while parents/guardians hold the infant.
Daily, morbidity data will be captured. If an infants found unwell, the study nurse will check on the infant and decide on treatment/referral to the MRC Keneba health centre. All parents/guardians will complete a weekly questionnaire asking whether they have offered foods or fluids other than breast milk.

###### 3.4.5 Concomitant medications/treatments

None.

#### 4 Selection and withdrawal of participants

##### 4.1 Selection of participants

Infants will be recruited at 6-10 weeks of age. The eligibility criteria will be explained to the parents/guardians and they will be invited to join the study by providing consent for their infant.

##### 4.2 Eligibility of participants

Participants must meet all the inclusion criteria and none of the exclusion criteria to be eligible to participate in the trial. Inclusion and exclusion criteria will be assessed during recruitment. All potentially eligible infants whose parents/guardians provide consent will be invited to the Baseline visit. At the Baseline visit a study nurse and FWs will assess the subjects and continue with the baseline activities. After which they will be formally enrolled into the study and assigned an intervention. A Study Specific Procedure (SSP) will be developed to give more details on this process.

###### 4.2.1 Inclusion criteria

- Infants (male or female) at 6 to 10 weeks of age.
- Breast fed infants (with plans to continue breastfeeding through six months of age).
- Parent/guardian with participant reside in study site area and are able and willing to adhere to all protocol visits and procedures (willingness to stay in the study area for the 14 weeks of supplementation).
- Healthy with no current illness and no chronic health problems.
- Signed or fingerprinted informed consent obtained from participants parent/guardian.

###### 4.2.2 Exclusion criteria

- LBW infants (ie less than 2.5kg at birth) or infants born prematurely (ie less than 37 weeks) will NOT be excluded.
- Formula fed infants or those whose parents/guardians are planning to use commercially available infant formular before six months of age.
- Acute illness (once acute illness is resolved, if appropriate, as per investigator assessment, participant may be re-revaluated for eligibility).
- Fever (for eligibility purpose defined as a body temperature greater than 37.5°C or mother report of fever) within 3 days prior to study initiation (once fever/acute illness is resolved, if appropriate, as per investigator assessment, participant may be re-revaluated for eligibility).
- Administration of any investigational drug within 30 days prior to study initiation or planned administration during the study period.
- Unwilling to avoid (their infant to avoid) the ingestion of supplements or herbal/other traditional medications during the study period.
- Any history of or evidence for chronic clinically significant (as per investigator assessment) disorder or disease (including, but not limited to, immunodeficiency, autoimmunity, congenital abnormality, bleeding disorder, and pulmonary, cardiovascular, metabolic, neurologic, renal, or hepatic disease).
- Any history of human immunodeficiency virus, chronic hepatitis B or chronic hepatitis C infections.
- History of meningitis, seizures, Guillain-Barré syndrome, or other neurological disorders.
- Any condition that in the opinion of the investigator might compromise the safety or well- being of the participant or compromise adherence to protocol procedures.

##### 4.3 Withdrawal of participants

Parents/guardians of infants have the right to freely withdraw the infant’s participation from a study at any time, without providing a rationale for their decision.

Every reasonable effort shall be made to keep infants in the study insofar as the parents allow and it is considered safe for the infant by the field staff. Parents/guardians who initially seek to withdraw their child from the study will be contacted by phone call or through a visit from study personnel explaining the importance of their continued participation to the study goals and the willingness to find alternative solutions to address the reasons for their withdrawal

The reason(s) for and date of infant withdrawal will be documented in relevant forms, with specification of the primary reason. Before entering any category as the reason for the infant's withdrawal from the study, the investigator should make every effort to investigate whether an AE may have been related to the participant's discontinuation from the study. If an AE has been associated with the participant's withdrawal, this must be described, even if it is not the primary reason for the participant's withdrawal.

- Study procedures and evaluations

For an overview see annex "Schedule of Events".

##### 4.4 Study schedule

###### 4.4.1 Screening & Enrollment (Baseline)

Healthy infants will be identified at 6-10 weeks of age. At screening, once parents/guardians of the infant have signed the informed consent form, the infants will be physically examined by a study nurse and, if the infant is considered as generally healthy (e.g. no fever, not severely malnourished), their height and weight will be measured. At study enrolment, a 3ml blood sample will be collected to assess the full blood count and reticulocyte count and determine iron status (Biochemistry analyser) as well as a faecal sample being collected for gut pathogen and feacal iron loss Infants with Hb < 7 g/dL will not be enrolled and referred to the regional health centre for treatment according to national guidelines. LBW infants and infants born prematurely will not be excluded.

###### 4.4.2 Follow-up

Infants will be randomised to 14 weeks of daily supplementation of either a) iron drops or b) placebo drops. Severely iron deficient (Hb <7 g/dL) infants or those with significant illness or any clinical syndromes that would affect interpretation will also be excluded. LBW infants and infants born prematurely will not be excluded.

The syrups with and without iron will be given to the participants daily. Both will be provided in flavoured drops. The iron will be provided at 7.5mg/day as elemental iron.

All infants will be monitored by the FWs for two weeks post supplementation.

###### 4.4.3 Final study visit

The final study visit will be after 14 weeks of iron supplementation. A second 3ml blood and fecal sample will be obtained as well as anthropometry performed.

###### 4.4.4 Early termination visit

An early termination visit may occur in this study because of a participant's voluntary withdrawal, trial team decision or at the discretion of the DSMB as described in Section 7. Apart from the safety evaluations, no other evaluations required for the final study visit will be done.

##### 4.5 Study evaluations

#### 4.5.1

This trial is powered to test if iron drops from 6-10 weeks of age are effective in increasing serum iron after 14 weeks (98 days) versus placebo. To detect an effect of 3.0µM, we intend to enrol 100 infants (total for both groups, with equal group size) in the trial. This will be sufficient to have 80% probability that, if the real ratio of the geometric mean serum iron concentration in the iron relative to the control group is 70% or more, the 95% CI for the ratio would exclude zero. A p-value of less than 0.05 will be considered statistically significant.

**Clinical evaluations**

FWs will record any AEs and ensure the safety of participants. Trained FWs will visit all infants daily during the supplementation period in order to administer the drops as described above and to check on the infant's health status. If an infant is found unwell, the study nurse will check on the infants and decide on treatment/referral to the MRC Keneba health centre. Parents/guardians will complete a weekly questionnaire asking whether they have offered foods or fluids other than breast milk.

###### 4.5.2 Laboratory evaluations

**Blood samples:**

At study enrolment, a 3ml blood sample will be collected (0.5 ml in EDTA tube and 2.5 ml in serum tube). At study termination (after 98 days of supplementation), a 3ml blood sample will be collected (0.5ml in EDTA and 2.5ml in a serum tube). Blood samples will be taken by venipuncture into EDTA and serum tubes and kept on ice. Within four hours of collection the samples will be processed by the MRCG laboratory. EDTA samples will be run on a hematology analyser to obtain full blood and reticulocytes count. Serum samples will be centrifuged, separated serum aliquots will be stored at -70°C. Samples will be analysed using a Biochemistry analyser for iron markers (ferritin, transferrin, transferrin saturation, unbound iron-binding capacity, soluble transferrin receptor), inflammatory markers (CRP/AGP). We will also use one of the serum aliquots to measure hepcidin concentration using an ELISA.

**Fecal samples:**

Iron content in fecal samples will be analysed by flame cytometery or biochemistry analyser.

#### 5 Safety considerations

This trial will be overseen by a DSMB).

The DSMB will be responsible for reviewing:

- To evaluate, on an ongoing basis, the accumulating solicited and unsolicited safety data (e.g. AEs, SAEs, deaths) from the infants enrolled in the trial and to advise on trial continuation on this basis. Safety data will be reviewed by the DSMB as overall pulled data (open session) and tabulated by coded treatment arm (closed session) at given time intervals. Unblinding of the treatment will only be requested by the DSMB in individual cases if there is a pattern of SAEs which may be related to study treatment where the DSMB feels there is 'potential for harm'.
- To monitor the rate of recruitment and level of retention of infants and to examine any trends apparent related to non-retention (e.g. consent withdrawal, loss to follow-up etc) and to provide advice accordingly.
- To assess data quality and completeness with the aim of ensuring the subsequent validity and credibility of the data generated.
- To consider factors external to the trial when relevant information becomes available, such as scientific or therapeutic developments that may have an impact on the safety of the participants or the ethics of the trial.
- To ensure at all times the confidentiality of the trial data and the DSMB discussions.
- To consider the ethical implications of any recommendations made.
- To report conflicts of interest guidelines as detailed in this charter.
- To document the outcome of all DSMB reviews and to provide the necessary information to the sponsor via the trial steering committee (TSC).
- To decide whether to recommend that the trial continues to recruit participants or whether recruitment should be stopped either for everyone or for some treatment groups and/or some participant subgroups.
- To suggest additional data analyses.
- The DSMB will not evaluate the efficacy or other non-safety endpoints during the trial although they may be asked to comment on these outcomes following trial completion. The majority of data related to these endpoints will only become available at this point.

Recruitment to the trial may be paused upon discretion of the DSMB if any infant has a serious adverse reaction (SAR) to any of the interventions administered during the trial. SAR is defined in the study protocol and constitutes any SAE where a causal relationship between the study supplements (or placebo) and the SAE is at least a reasonable probability, i.e. 'likely to be caused by' (probably related or definitely related). In an instance of need to pause the study, follow-up of the infants who have already been recruited will continue as planned pending the review of the DSMB.

Causality will be determined initially by the PIs in discussion with the study clinician, the clinical team, and with input from the Local Safety Monitor (LSM), considering the chronology of events and evidence for other causes – either from medical history, examination or further investigations. Such assessment, particularly in relation to 'expectedness', will take into consideration the safety profile of iron supplements; a conservative approach will be taken when there is doubt.

All SAR will be reported to the Sponsor and LSM by email using the standard SAE report for the trial within 24 hours of the investigators becoming aware.

The complete report of an SAR will be sent to the DSMB members as soon as the detailed report of the event is available (usually within 7 days of the investigator becoming aware). The DSMB chair or his deputy will acknowledge receipt of the report and any member of the DSMB may seek further information if judged to be required.

The DSMB will be asked to review all available data related to the SAR and advice on trial continuation. The DSMB will endeavour to respond within 7 days, initially by email to the TSC. Additional meetings may be necessary to make a full assessment of the reaction and its outcome, and these may be chaired by another member of the DSMB if necessary.

In addition to the DSMB, an independent (LSM) will regularly review all AEs and SAEs. This review will focus particularly on AEs causality and reasons for losses to follow up, raising any concerns or issues that present immediate safety concern with the PIs for reporting to the DSMB, while protecting the confidentiality of the trial data and the results of monitoring.

All deaths and SAE's related to the study iron supplements within 15 calendar days of receiving the report to the Medicines Control Agency, or within 7 calendar days if the event is fatal or life-threatening.

Experimental supplements shall be withheld for 7 days following detection of the following conditions:

1. Confirmed fever (axillary temperature >37.5°C) not associated with teething or vaccination[1];
2. Visually confirmed bloody diarrhoea;
3. Hospitalisation for somatic illness;
4. Treatment with antibiotics for any confirmed or suspected somatic infection[2].

Notes:

[1] This includes malaria (confirmed fever plus dipstick-test confirmed parasitaemia). In case of 48-hour history of fever as reported by the mother (or guardian) that is not confirmed axillary temperature ≤37.5°C), ask the mother/guardian to bring the child after 6 hours, and repeat measurement.

[1, 2] Suspected ear infections or urinary tract infections in children aged <six months shall be treated with antibiotics (check with physician) - thus, iron shall be withheld in these cases. If antibiotics are given for localised infections and there is no confirmed fever or a reported 48-hour history of fever, then supplemental iron can be given.

#### 5.1 Methods and timing for assessing, recording, and analysing safety parameters

This study will be conducted according to Good Clinical Practice (GCP) principles. The medical expert will be Dr Babucarr Susso, Head of Clinical Services at the MRCG field station in Keneba. The Local Safety Monitor will be Dr Patrick Nshe. The trial will be reviewed by the Gambia Government/MRC Joint Ethics Committee.

##### 5.1.1 Adverse events

An AE is defined as any untoward or unfavourable medical occurrence in a human subject, including signs and symptoms which are temporarily associated with the individual's participation in the research, whether considered related to the individual's participation in the research. Participants' parents/guardians will be asked verbally about AEs daily by the FWs during the study. Bi-weekly meeting with FWs will be held by the sub-investigator to summarise any AEs in a written report. There are no expected serious or moderate AEs associated with this study. However, a SSP will be drawn up to detail those symptoms that are more likely to be due to the intervention (e.g. stomach upset, diarrhoea), and those that are likely unrelated (e.g. infection-related symptoms). This process will continue after the end of supplementation for a further two weeks. In the unlikely case of an AE the FWs will immediately call the study coordinator to liaise with the study nurse as to whether a clinic referral is required. All FWs will be provided mobile credit for this purpose. All symptoms or signs reported or observed will be documented as an AE after evaluation by the study nurse or clinician. Any serious AE will be reported according to SOP-CTS-009 and followed up by study personnel until resolved or considered stable.

##### 5.1.2 Reactogenicity

None.

##### 5.1.3 Serious adverse events (SAEs)

A SAE is any AE that is life-threatening or results in death or requires hospitalisation or prolongation of hospitalisation or is a persistent or significant disability/incapacity. Though none are expected in this study, all SAEs will be investigated by a clinician.

##### 5.2 Reporting procedures

The *Medical Expert* will assess and document the severity or intensity of the AEs and laboratory changes as follows:

| Grade | Description |
|---|---|
| Mild | Awareness of sign or symptom, but easily tolerated |
| Moderate | Enough discomfort to cause interference with usual activity |
| Severe | Incapacitating with inability to work or do usual activity |
| Life-threatening | This grade will be considered as SAE |

The term “severe” is often used to describe the intensity (severity) of a specific event (as in mild, moderate, or severe myocardial infarction); the event itself, however, may be of relatively minor medical significance (such as severe headache). This is not the same as “serious”, which is based on the outcome or criteria defined under the SAE definition. An event can be considered serious without being severe if it conforms to the seriousness criteria; similarly, severe events that do not conform to the criteria are not necessarily serious. Seriousness (not severity) serves as a guide for defining regulatory reporting obligations. The PI shall report all SAEs without filtration, whether related to the intervention, within 24 hours of becoming aware of the event to the Sponsor. If the SAE is related to the intervention, The Gambia/MRC Joint Ethics Committee will be notified within seven calendar days if fatal or life-threatening, and all others within 15 calendar days. SAEs related to the study drugs and all deaths within 15 calendar days of receiving the report will be reported to the Medicines Control Agency, or within 7 calendar days if the event is fatal or life-threatening.

The minimum information required for this initial SAE report is:

- Trial number and (short) title.
- Participant’s ID.
- Nature of the event.
- Reporter’s name.

The PI will not wait for additional information to fully document the event before notifying. This initial report must be followed by a completed SAE Report within 2 working days, detailing relevant aspects of the SAE in question. All actions taken by the PI and the outcome of the event must also be reported immediately. For documentation of the SAE, any actions taken, outcome and follow-up, a SAE Report Forms will be used. All follow-up activities must be reported, if necessary, on one or more consecutive SAE report forms in a timely manner. All fields with additional or changed information must be completed and the report form should be forwarded to the Gambia/MRC Joint Ethics Committee within seven calendar days after receipt of the new information. Hospital case records and autopsy reports, including verbal autopsy, will be obtained where applicable.

##### 5.3 Safety oversight

Safety oversight shall be provided by the LSM who will provide independent advice. There will be a DSMB for this low risk trial. Dr Seth Adu-Afarwuah will be the chair of the DSMB. The LSM will also regularly review all AEs and SAEs. This review will focus particularly on AE’s causality and reasons for losses to follow up, raising any concerns or issues that present immediate safety concern with the named investigators for reporting to the medical expert, while protecting the confidentiality of the trial data and the results of monitoring.

#### 6 Discontinuation criteria

##### 6.1 Participant’s premature termination

Participants have the right to withdraw from the study at any time without giving a reason and this will not affect the medical care that would normally be received. The study team may also withdraw a participant from the study if deemed necessary at any time documenting the reason as one of the following:

- SAE
- AE.

- Participant's consent withdrawal.
- Migrated/moved from the study area.
- PI/SI initiated withdrawal

A 'lost to follow-up' is any participant who completed all protocol specific procedures up to the administration of the IP or intervention, but was then lost during the follow-up period, with no safety information and no efficacy endpoint data ever became available.

In case the participant decides to withdraw participation or consent during the study, we will not work on participant's samples without permission, but any information already generated from the samples until the time of withdrawal will be used and samples already collected, for which they have given consent, will also be analysed and data used. The study clinician may also ask for tests for the participant's safety. The PI/sub-investigator will ask about the reason for any withdrawal and follow-up with the participant regarding any unresolved AEs.

##### 6.2 Study discontinuation

A DSMB will oversee safety. The rules for study termination will be set by the DSMB at their first meeting.

#### 7 Statistical considerations

With regards to our primary endpoint (serum iron concentration at 98 days after randomisation), we consider intervention effects below 3.0 µM (absolute group difference in geometric means) to be irrelevant from medical and public health points of view.

We intend to enrol 100 infants (total for both groups, with equal group size) in the trial, which should be sufficient to have 80% probability that, if the real ratio of the geometric mean serum iron concentration in the iron relative to the control group is 70% or more, the 95% CI for the ratio would exclude zero [with the additional assumptions that the variance of $\log_e$ (serum iron concentration, µM) equals 0.76 for both groups (HIGH-kids); 10% of infants in the iron group stop taking iron supplements in the course of the intervention period; and the percentage of infants in the control group who stop taking placebo supplements is negligible.]

**FIGURE 2. Sample size requirements for various intervention effect sizes**

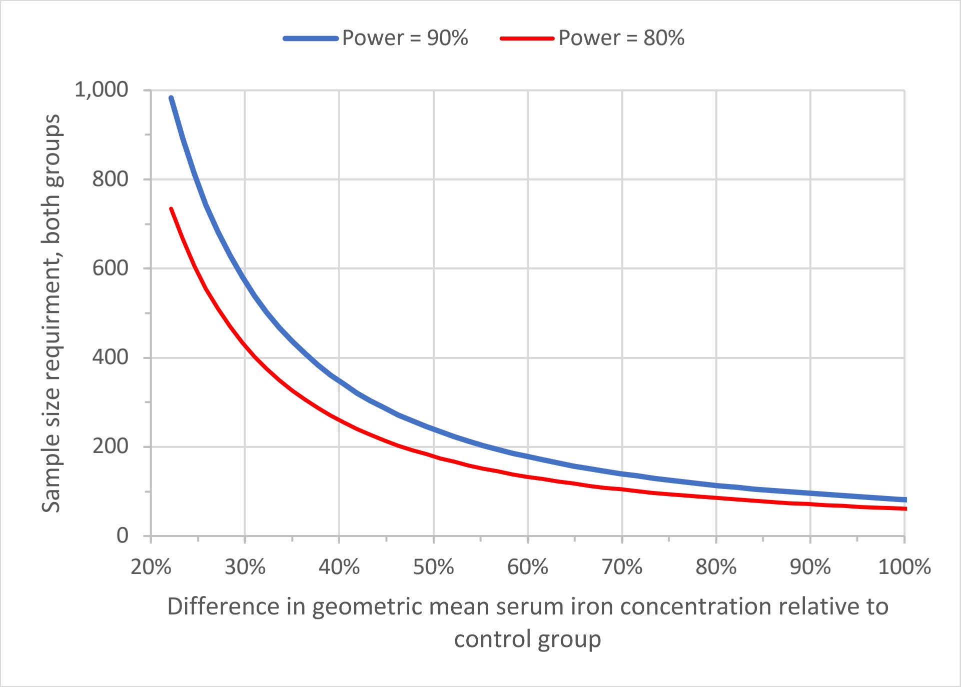

Statistical analysis will be perform using R studio version 1.3.1073/ STATA. Intervention groups will be described using conventional summary statistics (e.g., means or counts with SDs, medians with 25- and 75-centiles). The primary analysis concerns the effect of intervention on serum iron concentration at day 99 (i.e., 98 days of intervention with a one-day wash-out period after administration of the last supplement, assuming that a transient increase in serum iron concentration following iron supplementation will disappear within this period), adjusted for serum iron concentration measured at baseline. Continuous outcomes will be analysed by linear regression, with transformation of the outcome as needed to normalise or symmetrise the distribution of residuals. Analysis will be by modified intention-to-treat, i.e., all infants who were randomised and who received at least one supplemental dose will be included. We will also plan to perform per protocol analysis (i.e, to assess the efficacy that can be attained under controlled conditions) to determine the sensitivity of our modified intention-to-treat analysis. To account for missing data, we will use multiple imputations. In secondary analyses, we will adjust for any group imbalances that may occur in prognostic variables measured at baseline. Serum iron concentrations are known to undergo diurnal fluctuation (Dale et al. 2002), and they are known to be reduced in inflammation (Ganz and Nemeth, 2006). Thus, to account for these effects, we will also conduct secondary analyses with adjustment for the time of blood collection and for plasma concentrations of C-reactive and $\alpha_1$-acid glycoprotein at the time of blood collection.

Group differences in AEs will be analysed using negative binomial regression. In this analysis, we will count adverse events as the number of days with AEs divided by the total number of days that the infant was observed during the intervention period. In the analysis of diarrhoea, we will count an event if the parent/guardian reported three or more fluid stools (as indicated by stool chart, (Gustin *et al.*, 2018)) in the previous 24h recall period. To produce conservative effect estimates, analyses of adverse events will be per protocol.

Lastly, we will explore to what extent the magnitude of the intervention effect on serum iron concentration depends on iron status at baseline, as indicated by plasma concentrations of ferritin and soluble transferrin receptor (with adjustment for plasma concentrations of C-reactive and $\alpha_1$-acid glycoprotein at baseline as inflammation markers). A 95% confidence interval and a p-value of less than 0.05 will be considered statistically significant.

#### 8 Data handling and record keeping

##### 8.1 Data management and processing

**Type of Data:** Two types of data, qualitative and quantitative, will be collected

Qualitative: Data collected from questionnaires on demography, daily review of systems/health/morbidity from parents/guardians and once weekly feeding questionaires.

Quantitative: Data will include anthropometric measurements, vital signs, stool samples, haematology analysis, and biochemistry analysis.

##### 8.2 Format and scale of the data:

**Format and scale of the data:** REDCap software will be used. The final data format can be downloaded to Excel and common statistical packages (SPSS, SAS, Stata, R). It is specifically geared to support online or offline data capture for research studies and operations. This data management system is a fully supported externally validated (GCP compliant to 21 CFR parts 11) clinical database. The database will be hosted and managed by MRCG. Data on a total of 100 infants will be collected. The work will take place over the course of 1 year and infants will be followed as per protocol. All collected data will be subject to the metadata standards of the MRCG which render it fit for sharing and long-term validity. The trial dataset from REDCap uses an internationally recognised standard for its data representation known as Operational Data Model (ODM) which is a model endorsed by an internationally recognized clinical research standards body called Clinical Data Interchange Standards Consortium (CDISC). The final file format will be in CDISC ODM XML. ODM data format facilitates archiving, sharing, and interchange of metadata and data for clinical research.

**Methodologies for data collection/generation**

**Data will be collected as below:**

**Clinical data:** Will be collected by research nurses or research clinicians using the RedCap database accessed via benchtop computers or tablets.

**Laboratory data:** Blood will be collected by research nurses. Lab analysis will be performed by trained lab personnel.

All laboratory analysis results will be capture electronically. Each laboratory results will have unique study ID for each participant and the results will be automatically send to the central MRCG server or stored on the machine locally. The data manager will extract this data as agreed on the data management plan and uploaded into the REDCap project database.
Only those individuals who have been trained on the CRF/eCRFs and/or other document completion and have been authorized by the PI may complete CRFs and other study specific documentation. The study data collected on the eCRFs will be captured using devices i.e. tablets and/or laptops used by the study staff. Where there's internet connectivity, collected data will be stored directly onto the central server and when there is no/unreliable connectivity, data will be stored locally on the tablet and later synchronised to the central servers at MRCG in Fajara. Field staff will be trained to use the REDCap mobile application and clinicians will be trained to use the REDCap web portal.

**Data quality and standards:**
Review of data collection tools against the protocol will occur prior to database build by the DM to ensure that all data is being captured, at the correct time points and in a format conducive to the proposed statistical analysis. Site based staff will be fully trained in conducting any study assessments and subsequent completion of the eCRF. A site delegation log will be maintained. Study staff will be trained in the use of REDCap. Principles of good clinical practice will be adhered to and all training will be documented. To ensure standardisation of processes, standard operating procedures with respect to trial management, quality assurance, data management, IT security and statistics will be adhered to. The entry screens will be designed with range checks, skip patterns and validations to assist/guide data collectors and entry clerks to ensure high level accuracy. Multiple choice options will be employed when and where applicable. Mandatory field checks and other computable data will be prepopulated to eliminate errors. All staff will be trained on standard operating procedures (SOPs), SSPs, and the protocol prior to the start of the study. The data manager will conduct periodical data cleaning routines to flag data queries that were not picked up at earlier stages. All data queries raised by the data manager will be answered in writing by the sub-investigator normally within one week. All changes to any CRFs or databases will be governed by a change control SOP, ensuring that all sites are using the same version of both CRFs and databases (see MRC Unit The Gambia's SOPs on Database Development, Data Entry, Data Verification, Change Control and Data Query Management).

#### 8.3 Source documents and access to source data

The PI will maintain appropriate medical and research records for this study in compliance with the principles of GCP and regulatory and institutional requirements for the protection of confidentiality of participants. The study team members will have access to records.

The authorised representatives of the sponsor, the ethics committee(s) or regulatory bodies may inspect all documents and records required to be maintained by the investigator, including but not limited to, medical records (office, clinic, or hospital) for the participants in this study. The clinical study site will permit access to such records.

#### 8.4 Protocol deviations

A protocol deviation (PD) is any noncompliance with the clinical trial protocol, GCP, or other applicable regulatory requirements. The noncompliance may be either on the part of the participant or the investigator including the study team members, and may result in significant added risk to the study participant. As a result of a deviation, corrective actions will be developed and implemented promptly.

If a deviation from, or a change of, the protocol is implemented to eliminate an immediate hazard(s) to trial participant without prior ethics approval, the PI or designee will submit the implemented deviation or change, the reasons for it, and, if appropriate, the proposed protocol amendment(s) as soon as possible to the sponsor for agreement and the relevant independent ethics committee for review and approval.

The PI or designee will document and explain any deviation from the approved protocol on the CRF, where appropriate, and record and explain any deviation in a protocol deviation form that will be maintained as an essential document.

### 9 QUALITY CONTROL AND QUALITY ASSURANCE

Quality control will be applied to each stage of the study. It will be the responsibility of the PI or designated trial team member to ensure that all source documents and CRFs are reviewed for accuracy and completeness. Any correction will be accurately accounted for.

Dispensing of the supplements to the village level, together with the breast-feeding information will be done by the FWs or study nurse. Venous blood collection will be done by the study nurse or a clinician working with the trial team.

All procedures will be recorded on the CRF. All sample tubes will be labelled with ID barcodes. Quality control material will be run on the Cobas Integra equipment before analysing the samples in order to ensure precision and accuracy.

All Field assistants and their supervisors, study nurses and FWs will be trained. Weekly meetings of the entire trial field team will be convened in order to discuss all problems and lessons from the study.

Accountability of IPs report will be submitted at end of study and unused IPs will only be destroyed after written approval from MCA.

#### 9.1 Study monitoring

The study may be subject audit by the London School of Hygiene & Tropical Medicine under their remit as sponsor, the Study Coordination Centre and other regulatory bodies to ensure adherence to GCP.

Risk-based trial monitoring will be conducted by the designated monitor of the MRCG Clinical Trials Support Office in keeping with the approved monitoring plan. The monitoring of study will follow the Unit's SOP-CTS-005 on Monitoring. An initiation visit will occur, and formal study start approval obtained from the Sponsor before the start of recruitment. Interim visits will be conducted during the conduct of the study in line with the monitoring plan. At the end of the study, a close out visit will be conducted after last participant last visit and database cleaning completed and ready for database lock.

### 10 Ethical considerations

This study is conducted in accordance with the principles set forth in the ICH Harmonised Tripartite Guideline for Good Clinical Practice and the Declaration of Helsinki in its current version (see appendix), whichever affords the greater protection to the participants. The study will be reviewed by The Scientific Coodinating Committee of MRC Unit The Gambia at The London School of Hygiene and Tropical Medicine, The Gambian Government/MRC Unit The Gambia Joint Ethics Committee and the Ethics Committees at the The London School of Hygiene and Tropical Medicine and Kings College London.

#### 10.1 General considerations on human subject protection

##### 10.1.1 Rationale for participant selection

Before testing the impact of early post-natal iron supplementation on neurological development and cognitive performance it is important to run a proof-of-concept trial. The participants enrolled in this pilot trial will be infants selected as healthy volunteers to help test the efficacy of the supplement on lowering ID and on supplementation and breast-feeding compliance. If the supplement works well in this group, it will provide the justification to scale up to a larger trial run in a large-scale randomised control trial.

##### 10.1.2 Evaluation of risks and benefits

There has been a long-standing controversy regarding iron and infections after the Pemba Trial indicated an increase in malaria-related hospital admissions. Malaria rates in West and East Kiang are now extremely low. In our past two trials involving almost 1000 pregnant women and children 6-24m (HAPn and HIGH-kids) studied for three months each we had only five positive malaria tests. Even in the IHAT Trial in the North Bank around Basse (509 children 6-36m studied for three months each) we had only four cases of malaria and notably three of these were in the placebo group receiving no iron. We have also investigated the possible effects of iron on other health outcomes, on adverse events and on serious adverse events. In the children's trials (HIGH-kids and IHAT) we had ~250 child-years of daily follow-up. The HIGH-kids Study had three groups (A: Gambian Government standard of care = 12mg/d iron; B) a screen & treat group that overall had 6mg/d iron; and C) a screen & treat group that overall had 3mg/d iron). There were no differences in the numbers of AEs or SAEs between groups. There were no deaths. The IHAT Study also had three groups (iron in a novel IHAT supplement, iron as the traditional FeSO4, and no iron (placebo)). Again, there were no significant differences in any AEs or SAEs or in the gut microbiome. In fact, there was a slight tendency for the placebo group to have worse outcomes. There was one death unrelated to the interventions (peanut asphyxiation). Thus, in summary, although we do not have data on young infants all of our recent data has refuted the prior suggestion that children will be put at risk by receiving iron. As with many other MRCG studies our data show that participation in a study is highly protective against serious adverse outcomes and death. Nonetheless, all infants will be visited daily, both to ensure compliance with taking the iron or placebo and to ensure their safety. Any unwell infant will be reviewed by a nurse and immediately referred to a physician if there is any significant health issue.

**10.2 Informed consent**

Individual consent for the study will be sought. FWs will be trained to explain the project in full detail to the eligible participants parents, covering all aspects of such study as laid out in the 'Participant Information Sheet'. Literate parents will then be given the Information Sheet while illiterate parents will have the full Information Sheet read to them in the language they understand. Illiterate consenting subjects will require an independent literate witness. Any questions that arise will be answered by the FWs and will also be given the possibility to obtain further clarifications and explanations by speaking to one of the study investigators if they wish. Participation is entirely voluntary, and we do not intend to enroll individuals whose parents are not able to give consent. If parents agree to enroll their infant, written Informed Consent will be obtained.

**10.3 Participant confidentiality**

Any participants' identifiable data collected by the Study Coordination Centre will be stored securely and their confidentiality protected in accordance with the Data Protection Act. Participant confidentiality, privacy and anonymity will always be ensured. All data will be anonymised and individuals will not be identifiable. The study will be run in compliance with the MRC Corporate Information security Policy and the MRC Unit The Gambia's INFORMATION AND COMMUNICATION TECHNOLOGY SECURITY POL-INT-001 available on the MRC Gambia intranet (Available on request).

The risk to confidentiality is the personal information on subjects kept by the PI for study follow up verification purposes. These will be kept separate from the study data and not available to the study staff so that no linkages of personal data to study records would be easily made. The study data are stored in a limited access, password protected database so that only staff who have the required permissions can view the study records. Personal information will not be available to and study staff except the PI and the datamanger.

Data about the participants may be shared via a public data repository or by sharing directly with other researchers, participant information will not be identifiable from this information.

**10.4 Future use of stored specimen**

Aliquots of blood and faecal samples will be kept frozen at -70ºC for future analysis. This may include DNA analysis and export of samples. We will obtain informed consent from the parents/guardians for this to be the case within the study informed consent. The blood and faecal samples collected during the trial may be used to support other research in the future, and may be shared anonymously with other researchers, for their ethically-approved projects. Any future use would require PI, MRCG SCC and EC approval.

**11 Financing and insurance**

London School of Hygiene & Tropical Medicine holds Public Liability ("negligent harm") and Clinical Trial ("non-negligent harm") insurance policies which apply to this trial.

Funding for the trial has been received from DFID/NIHR/MRC/Wellcome Joint Global Health Trials - Call 9. All funds will be used in The Gambia. The study will benefit from the infrastructure already in place at MRCG at LSHTM.

**12 Publication policy**

All key findings from this study will be submitted for publication in peer-reviewed journals. Our planned dissemination avenues include: at least three publications in high impact peer-reviewed open-access scientific journals with a wide readership (e.g. Lancet, JAMA, NEJM), presentation at international conferences (e.g. the

Micronutrient Forum) and dissemination of the trial findings to organizations such as the WHO, UNICEF, UN World Food Programme and to the National Nutrition Agency and the Ministry of Health in The Gambia.

**Supplements, appendices and other documents**

**Schematic of Study Design**:

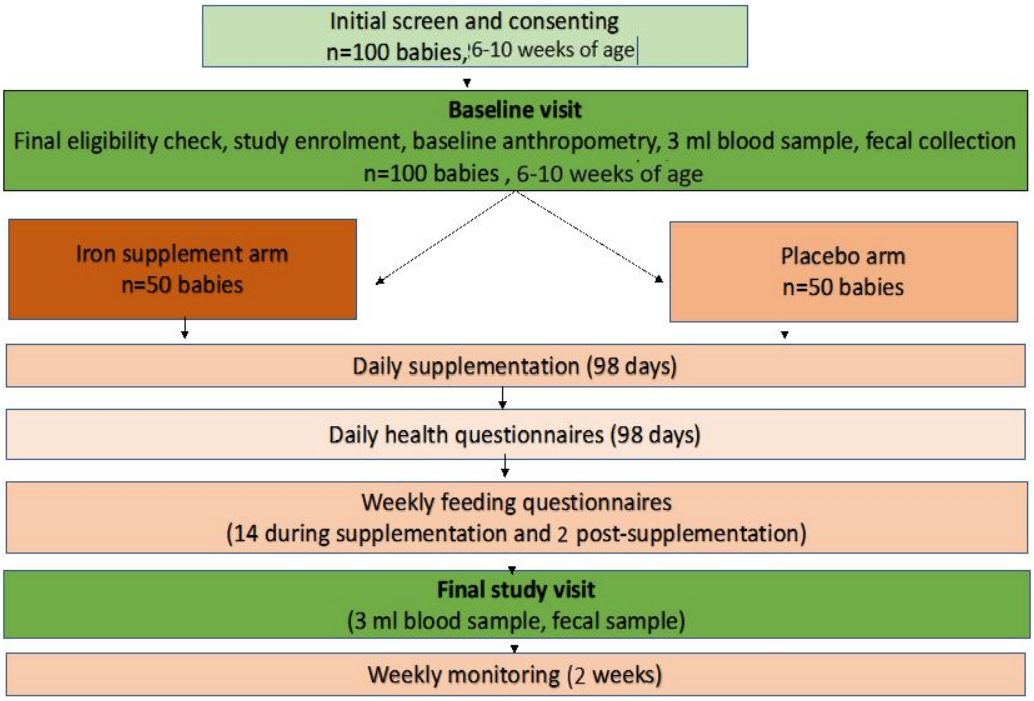

**Appendix: Schedule of events**

| Activities | 2021 | | | | | | | | | | | | 2022 | | | | | |
|---|---|---|---|---|---|---|---|---|---|---|---|---|---|---|---|---|---|---|
| | Q1 | | | Q2 | | | Q3 | | | Q4 | | | Q1 | | | Q2 | | |
| | J | F | M | A | M | J | J | A | S | O | N | D | J | F | M | A | M | J |
| Ethics and study set up | | | | | | | | | | | | | | | | | | |
| Sensitisation | | | | | | | | | | | | | | | | | | |
| Recruitment/consent (rolling) | | | | | | | | | | | | | | | | | | |
| Sample collection and iron analysis (in Keneba) | | | | | | | | | | | | | | | | | | |
| Microbiome analysis | | | | | | | | | | | | | | | | | | |
| Data cleaning, analysis, write-up | | | | | | | | | | | | | | | | | | |
| Writing of RCT trial funding application | | | | | | | | | | | | | | | | | | |

**Appendix:**

**WORLD MEDICAL ASSOCIATION DECLARATION OF HELSINKI**
**Ethical Principles for Medical Research Involving Human Subjects**

Adopted by the 18th WMA General Assembly, Helsinki, Finland, June 1964
and amended by the:
29th WMA General Assembly, Tokyo, Japan, October 1975
35th WMA General Assembly, Venice, Italy, October 1983
41st WMA General Assembly, Hong Kong, September 1989
48th WMA General Assembly, Somerset West, Republic of South Africa, October 1996
52nd WMA General Assembly, Edinburgh, Scotland, October 2000
53rd WMA General Assembly, Washington DC, USA, October 2002 (Note of Clarification added)
55th WMA General Assembly, Tokyo, Japan, October 2004 (Note of Clarification added)
59th WMA General Assembly, Seoul, Republic of Korea, October 2008
64th WMA General Assembly, Fortaleza, Brazil, October 2013

**Preamble**

1. The World Medical Association (WMA) has developed the Declaration of Helsinki as a statement of ethical principles for medical research involving human subjects, including research on identifiable human material and data.
The Declaration is intended to be read as a whole and each of its constituent paragraphs should be applied with consideration of all other relevant paragraphs.
2. Consistent with the mandate of the WMA, the Declaration is addressed primarily to physicians. The WMA encourages others who are involved in medical research involving human subjects to adopt these principles.

**General Principles**

3. The Declaration of Geneva of the WMA binds the physician with the words, “The health of my patient will be my first consideration,” and the International Code of Medical Ethics declares that, “A physician shall act in the patient's best interest when providing medical care.”
4. It is the duty of the physician to promote and safeguard the health, well-being and rights of patients, including those who are involved in medical research. The physician's knowledge and conscience are dedicated to the fulfilment of this duty.
5. Medical progress is based on research that ultimately must include studies involving human subjects.
6. The primary purpose of medical research involving human subjects is to understand the causes, development and effects of diseases and improve preventive, diagnostic and therapeutic interventions (methods, procedures and treatments). Even the best proven interventions must be evaluated continually through research for their safety, effectiveness, efficiency, accessibility and quality.
7. Medical research is subject to ethical standards that promote and ensure respect for all human subjects and protect their health and rights.
8. While the primary purpose of medical research is to generate new knowledge, this goal can never take precedence over the rights and interests of individual research subjects.
9. It is the duty of physicians who are involved in medical research to protect the life, health, dignity, integrity, right to self-determination, privacy, and confidentiality of personal information of research subjects. The responsibility for the protection of research subjects must always rest with the physician or other health care professionals and never with the research subjects, even though they have given consent.
10. Physicians must consider the ethical, legal and regulatory norms and standards for research involving human subjects in their own countries as well as applicable international norms and standards. No national or international ethical, legal or regulatory requirement should reduce or eliminate any of the protections for research subjects set forth in this Declaration.
11. Medical research should be conducted in a manner that minimises possible harm to the environment.

12. Medical research involving human subjects must be conducted only by individuals with the appropriate ethics and scientific education, training and qualifications. Research on patients or healthy volunteers requires the supervision of a competent and appropriately qualified physician or other health care professional.
13. Groups that are underrepresented in medical research should be provided appropriate access to participation in research.
14. Physicians who combine medical research with medical care should involve their patients in research only to the extent that this is justified by its potential preventive, diagnostic or therapeutic value and if the physician has good reason to believe that participation in the research study will not adversely affect the health of the patients who serve as research subjects.
15. Appropriate compensation and treatment for subjects who are harmed as a result of participating in research must be ensured.

**Risks, Burdens and Benefits**

16. In medical practice and in medical research, most interventions involve risks and burdens.
Medical research involving human subjects may only be conducted if the importance of the objective outweighs the risks and burdens to the research subjects.
17. All medical research involving human subjects must be preceded by careful assessment of predictable risks and burdens to the individuals and groups involved in the research in comparison with foreseeable benefits to them and to other individuals or groups affected by the condition under investigation.
Measures to minimise the risks must be implemented. The risks must be continuously monitored, assessed and documented by the researcher.
18. Physicians may not be involved in a research study involving human subjects unless they are confident that the risks have been adequately assessed and can be satisfactorily managed.
When the risks are found to outweigh the potential benefits or when there is conclusive proof of definitive outcomes, physicians must assess whether to continue, modify or immediately stop the study.

**Vulnerable Groups and Individuals**

19. Some groups and individuals are particularly vulnerable and may have an increased likelihood of being wronged or of incurring additional harm.
All vulnerable groups and individuals should receive specifically considered protection.
20. Medical research with a vulnerable group is only justified if the research is responsive to the health needs or priorities of this group and the research cannot be carried out in a non-vulnerable group. In addition, this group should stand to benefit from the knowledge, practices or interventions that result from the research.

**Scientific Requirements and Research Protocols**

21. Medical research involving human subjects must conform to generally accepted scientific principles, be based on a thorough knowledge of the scientific literature, other relevant sources of information, and adequate laboratory and, as appropriate, animal experimentation. The welfare of animals used for research must be respected.
22. The design and performance of each research study involving human subjects must be clearly described and justified in a research protocol.
The protocol should contain a statement of the ethical considerations involved and should indicate how the principles in this Declaration have been addressed. The protocol should include information regarding funding, sponsors, institutional affiliations, potential conflicts of interest, incentives for subjects and information regarding provisions for treating and/or compensating subjects who are harmed as a consequence of participation in the research study.
In clinical trials, the protocol must also describe appropriate arrangements for post-trial provisions.

**Research Ethics Committees**

23. The research protocol must be submitted for consideration, comment, guidance and approval to the concerned research ethics committee before the study begins. This committee must be transparent in its functioning, must be independent of the researcher, the sponsor and any other undue influence and must be duly qualified. It must take into consideration the laws and regulations of the country or countries in which the research is to be performed as

well as applicable international norms and standards but these must not be allowed to reduce or eliminate any of the protections for research subjects set forth in this Declaration.
The committee must have the right to monitor ongoing studies. The researcher must provide monitoring information to the committee, especially information about any serious adverse events. No amendment to the protocol may be made without consideration and approval by the committee. After the end of the study, the researchers must submit a final report to the committee containing a summary of the study's findings and conclusions.

**Privacy and Confidentiality**

24. Every precaution must be taken to protect the privacy of research subjects and the confidentiality of their personal information.

**Informed Consent**

25. Participation by individuals capable of giving informed consent as subjects in medical research must be voluntary. Although it may be appropriate to consult family members or community leaders, no individual capable of giving informed consent may be enrolled in a research study unless he or she freely agrees.

26. In medical research involving human subjects capable of giving informed consent, each potential subject must be adequately informed of the aims, methods, sources of funding, any possible conflicts of interest, institutional affiliations of the researcher, the anticipated benefits and potential risks of the study and the discomfort it may entail, post-study provisions and any other relevant aspects of the study. The potential subject must be informed of the right to refuse to participate in the study or to withdraw consent to participate at any time without reprisal. Special attention should be given to the specific information needs of individual potential subjects as well as to the methods used to deliver the information.
After ensuring that the potential subject has understood the information, the physician or another appropriately qualified individual must then seek the potential subject's freely-given informed consent, preferably in writing. If the consent cannot be expressed in writing, the non-written consent must be formally documented and witnessed.
All medical research subjects should be given the option of being informed about the general outcome and results of the study.

27. When seeking informed consent for participation in a research study the physician must be particularly cautious if the potential subject is in a dependent relationship with the physician or may consent under duress. In such situations the informed consent must be sought by an appropriately qualified individual who is completely independent of this relationship.

28. For a potential research subject who is incapable of giving informed consent, the physician must seek informed consent from the legally authorised representative. These individuals must not be included in a research study that has no likelihood of benefit for them unless it is intended to promote the health of the group represented by the potential subject, the research cannot instead be performed with persons capable of providing informed consent, and the research entails only minimal risk and minimal burden.

29. When a potential research subject who is deemed incapable of giving informed consent is able to give assent to decisions about participation in research, the physician must seek that assent in addition to the consent of the legally authorised representative. The potential subject's dissent should be respected.

30. Research involving subjects who are physically or mentally incapable of giving consent, for example, unconscious patients, may be done only if the physical or mental condition that prevents giving informed consent is a necessary characteristic of the research group. In such circumstances the physician must seek informed consent from the legally authorised representative. If no such representative is available and if the research cannot be delayed, the study may proceed without informed consent provided that the specific reasons for involving subjects with a condition that renders them unable to give informed consent have been stated in the research protocol and the study has been approved by a research ethics committee. Consent to remain in the research must be obtained as soon as possible from the subject or a legally authorised representative.

31. The physician must fully inform the patient which aspects of their care are related to the research. The refusal of a patient to participate in a study or the patient's decision to withdraw from the study must never adversely affect the patient-physician relationship.

32. For medical research using identifiable human material or data, such as research on material or data contained in biobanks or similar repositories, physicians must seek informed consent for its collection, storage and/or reuse. There may be exceptional situations where consent would be impossible or impracticable to obtain for such research. In such situations the research may be done only after consideration and approval of a research ethics committee.

**Use of Placebo**

33. The benefits, risks, burdens and effectiveness of a new intervention must be tested against those of the best proven intervention(s), except in the following circumstances:

Where no proven intervention exists, the use of placebo, or no intervention, is acceptable; or

Where for compelling and scientifically sound methodological reasons the use of any intervention less effective than the best proven one, the use of placebo, or no intervention is necessary to determine the efficacy or safety of an intervention

and the patients who receive any intervention less effective than the best proven one, placebo, or no intervention will not be subject to additional risks of serious or irreversible harm as a result of not receiving the best proven intervention.

Extreme care must be taken to avoid abuse of this option.

**Post-Trial Provisions**

34. In advance of a clinical trial, sponsors, researchers and host country governments should make provisions for post-trial access for all participants who still need an intervention identified as beneficial in the trial. This information must also be disclosed to participants during the informed consent process.

**Research Registration and Publication and Dissemination of Results**

35. Every research study involving human subjects must be registered in a publicly accessible database before recruitment of the first subject.

36. Researchers, authors, sponsors, editors and publishers all have ethical obligations with regard to the publication and dissemination of the results of research. Researchers have a duty to make publicly available the results of their research on human subjects and are accountable for the completeness and accuracy of their reports. All parties should adhere to accepted guidelines for ethical reporting. Negative and inconclusive as well as positive results must be published or otherwise made publicly available. Sources of funding, institutional affiliations and conflicts of interest must be declared in the publication. Reports of research not in accordance with the principles of this Declaration should not be accepted for publication.

**Unproven Interventions in Clinical Practice**

37. In the treatment of an individual patient, where proven interventions do not exist or other known interventions have been ineffective, the physician, after seeking expert advice, with informed consent from the patient or a legally authorised representative, may use an unproven intervention if in the physician's judgement it offers hope of saving life, re-establishing health or alleviating suffering. This intervention should subsequently be made the object of research, designed to evaluate its safety and efficacy. In all cases, new information must be recorded and, where appropriate, made publicly available.
