## Supplementary material for "Early iron supplementation of exclusively breastfed African infants: a proof-of-principle, placebo-controlled, randomised, double-blinded efficacy trial": Figure S4

Bah et al_Supplementary Fig S4

This file is also available as a Microsoft Excel file here:
https://figshare.com/articles/dataset/Adverse_events_table/21878706

Key to variables:

**ID:** study identification number;
**treat:** intervention group (iron or placebo; groups are separated by a yellow bar);
**N-epi:** number of episodes;
**Totd:** Total duration (number of days with reported condition);
**obs:** total observation time

**Light red/light blue/light green:** condition of interest reported to be absent;
**Dark red/dark blue/dark green:** condition of interest reported to be present;
**White cells:** censored (child not present or no report received).

Individual episodes (see text for case definitions) are indicated by borders (black or red)

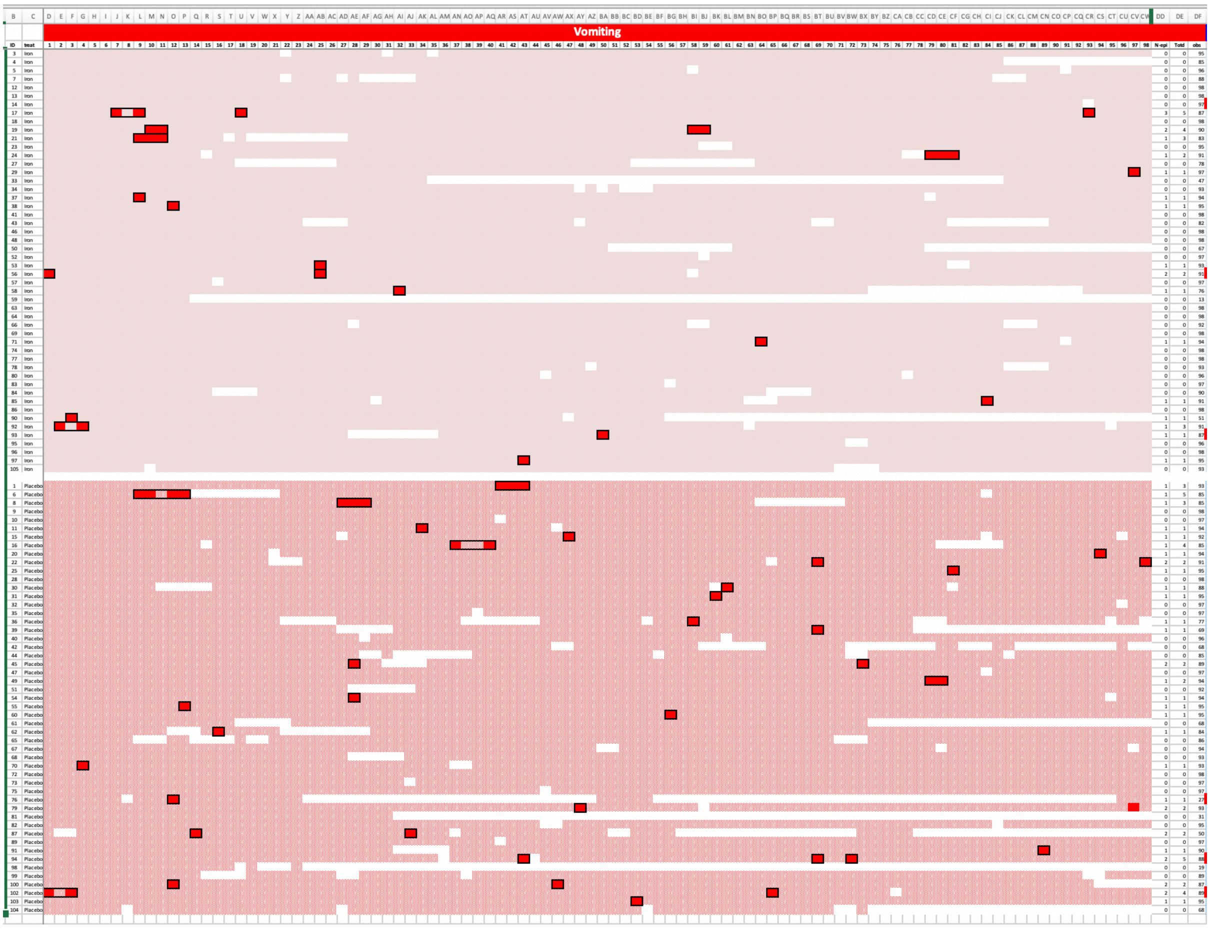
Vomiting

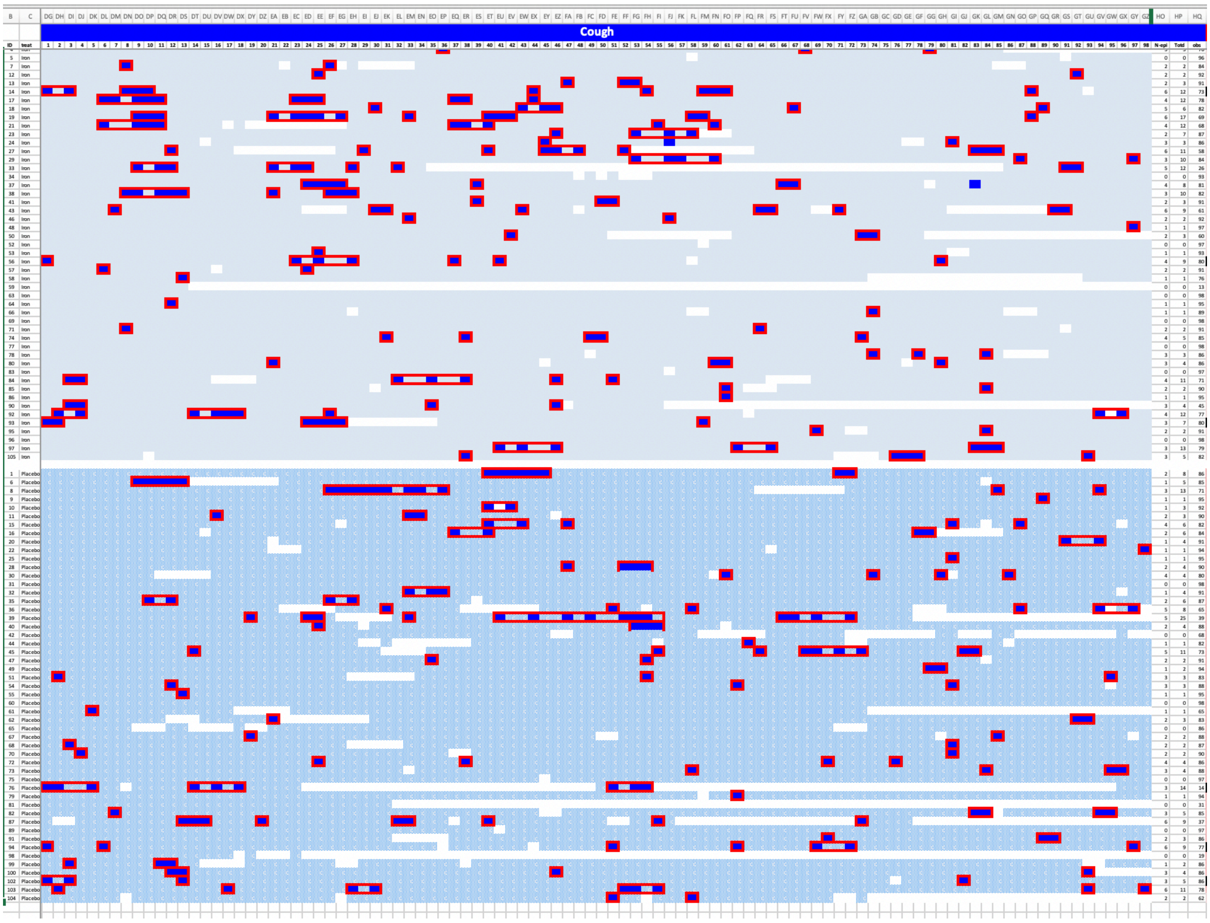

Cough

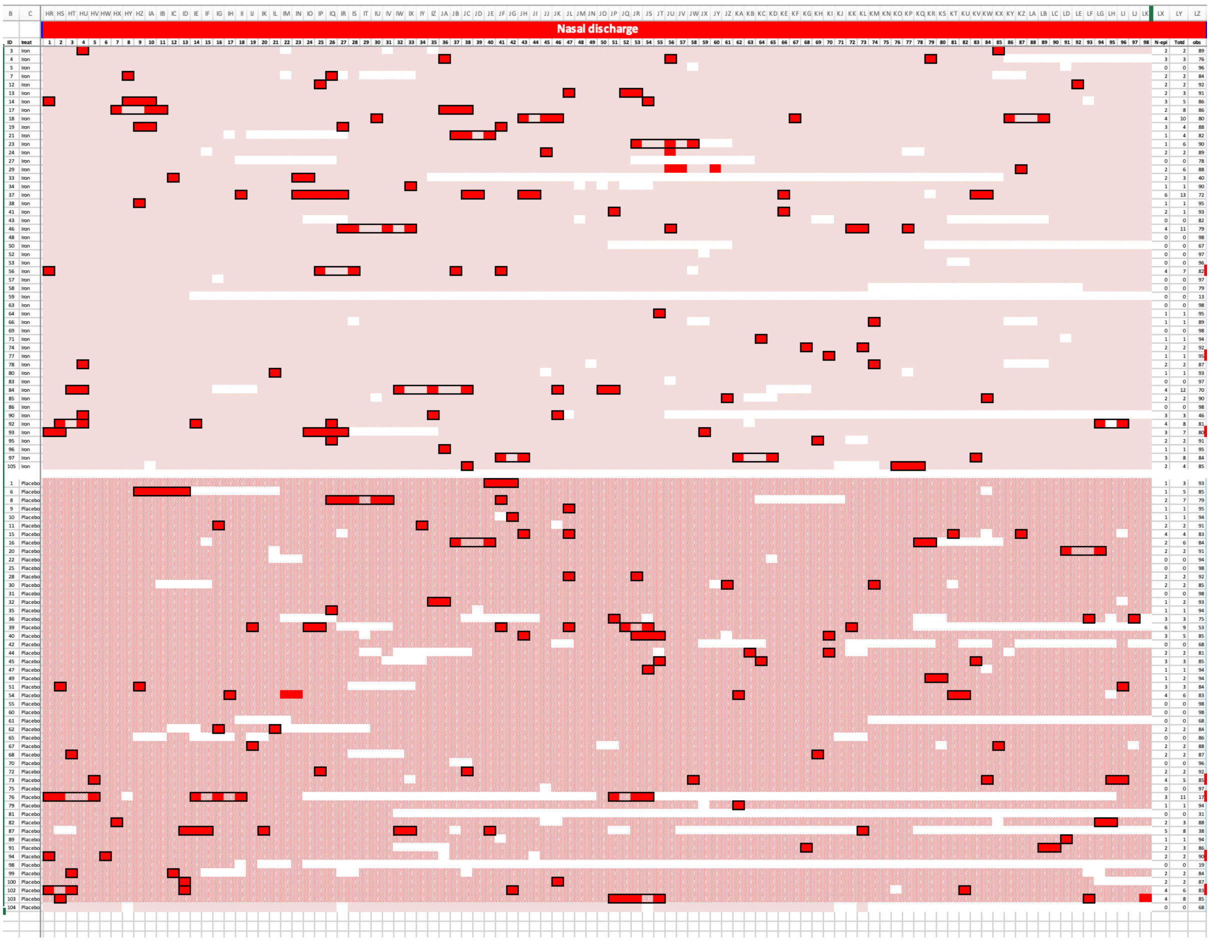
Nasal discharge

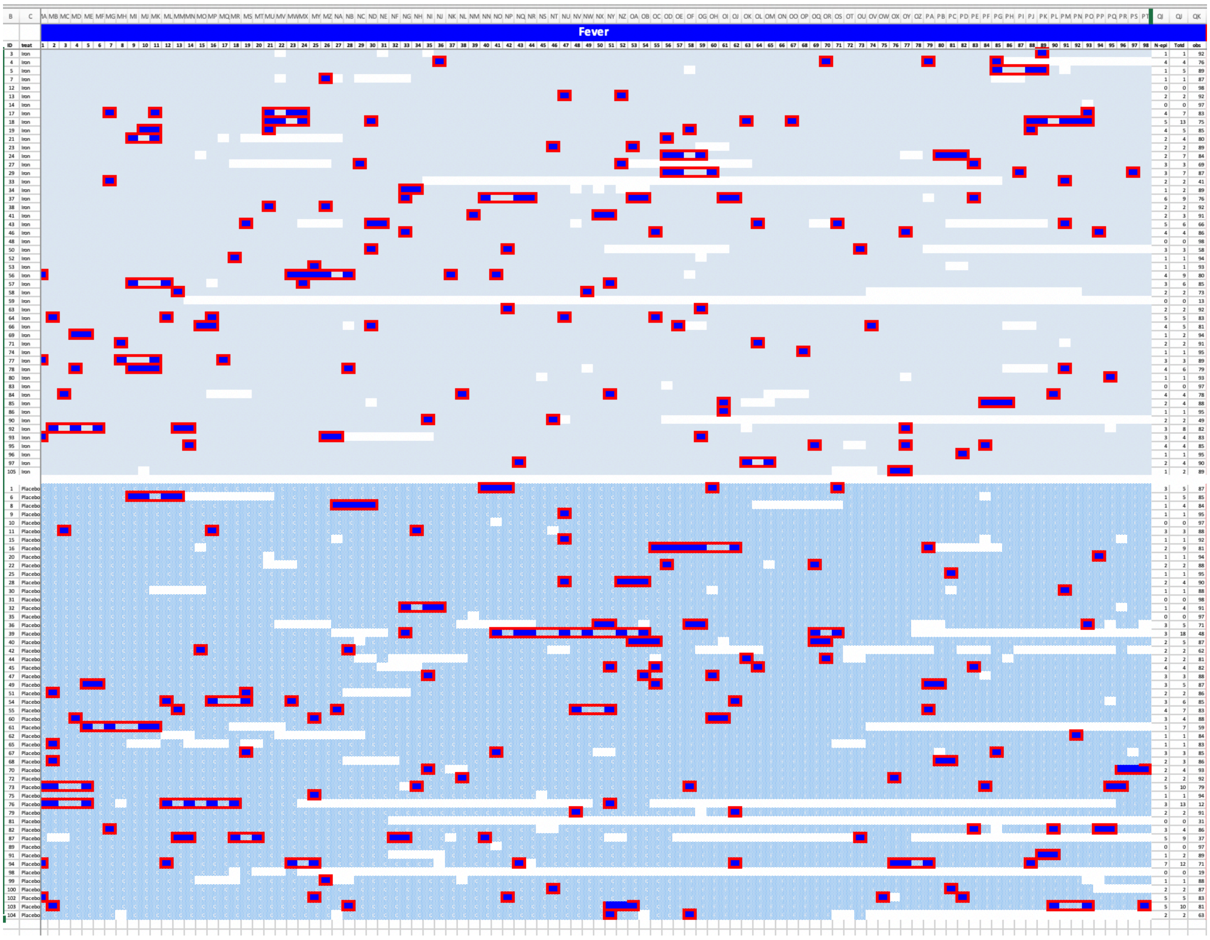

Fever

### Skin infection

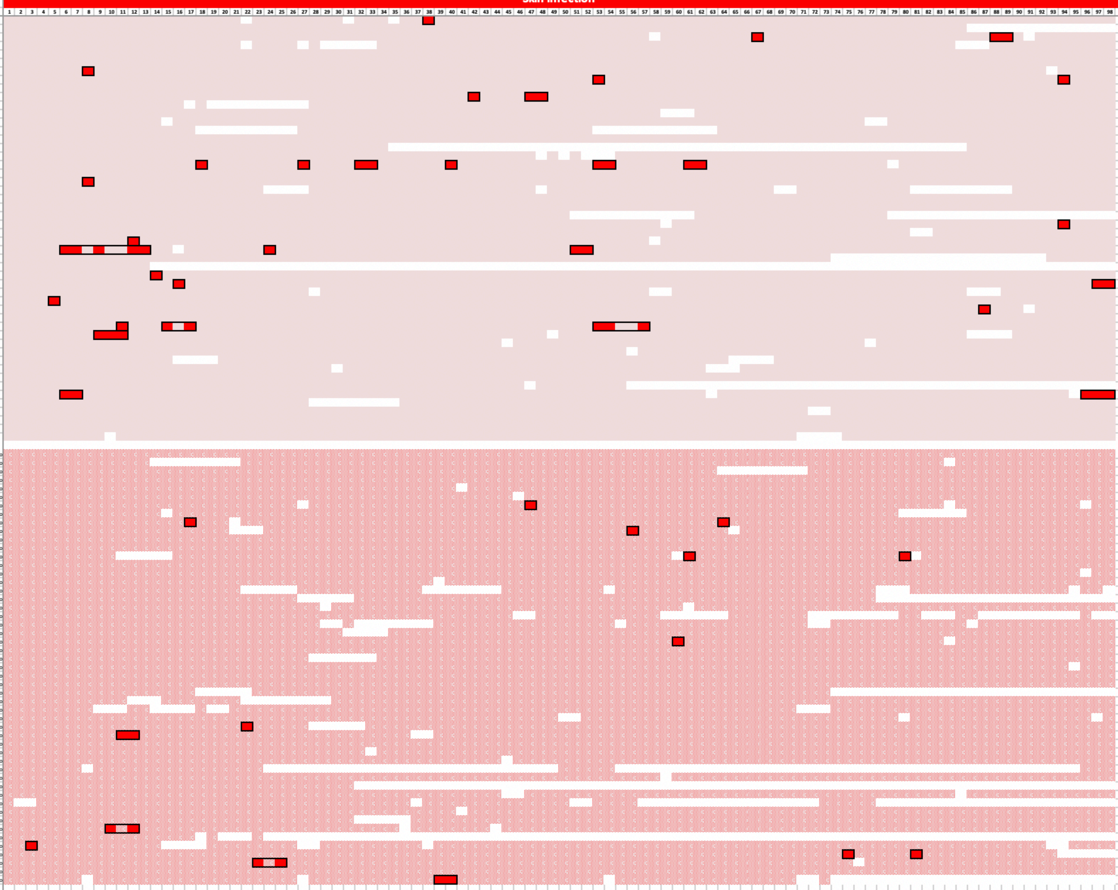

| ID | treat | N-epi | Total | obs |
|---|---|---|---|---|
| 3 | Iron | 1 | 1 | 92 |
| 4 | Iron | 0 | 0 | 85 |
| 5 | Iron | 2 | 3 | 90 |
| 7 | Iron | 0 | 0 | 88 |
| 12 | Iron | 0 | 0 | 98 |
| 13 | Iron | 0 | 0 | 98 |
| 14 | Iron | 1 | 1 | 94 |
| 17 | Iron | 1 | 1 | 95 |
| 18 | Iron | 0 | 0 | 98 |
| 19 | Iron | 2 | 3 | 91 |
| 21 | Iron | 0 | 0 | 88 |
| 23 | Iron | 0 | 0 | 95 |
| 24 | Iron | 0 | 0 | 95 |
| 27 | Iron | 0 | 0 | 78 |
| 29 | Iron | 0 | 0 | 98 |
| 33 | Iron | 0 | 0 | 47 |
| 34 | Iron | 0 | 0 | 93 |
| 37 | Iron | 6 | 9 | 76 |
| 38 | Iron | 0 | 0 | 98 |
| 41 | Iron | 1 | 1 | 95 |
| 43 | Iron | 0 | 0 | 82 |
| 46 | Iron | 0 | 0 | 98 |
| 48 | Iron | 0 | 0 | 98 |
| 50 | Iron | 0 | 0 | 67 |
| 52 | Iron | 1 | 1 | 94 |
| 53 | Iron | 0 | 0 | 96 |
| 56 | Iron | 1 | 1 | 94 |
| 57 | Iron | 2 | 10 | 84 |
| 58 | Iron | 0 | 0 | 79 |
| 59 | Iron | 0 | 0 | 13 |
| 63 | Iron | 1 | 1 | 95 |
| 64 | Iron | 2 | 3 | 94 |
| 66 | Iron | 0 | 0 | 92 |
| 69 | Iron | 1 | 1 | 95 |
| 71 | Iron | 1 | 1 | 94 |
| 74 | Iron | 0 | 0 | 98 |
| 77 | Iron | 3 | 9 | 83 |
| 78 | Iron | 1 | 3 | 88 |
| 80 | Iron | 0 | 0 | 96 |
| 83 | Iron | 0 | 0 | 97 |
| 84 | Iron | 0 | 0 | 90 |
| 85 | Iron | 0 | 0 | 94 |
| 86 | Iron | 0 | 0 | 98 |
| 90 | Iron | 0 | 0 | 54 |
| 92 | Iron | 2 | 5 | 90 |
| 93 | Iron | 0 | 0 | 90 |
| 95 | Iron | 0 | 0 | 96 |
| 96 | Iron | 0 | 0 | 98 |
| 97 | Iron | 0 | 0 | 98 |
| 105 | Iron | 0 | 0 | 93 |
| 1 | Placebo | 0 | 0 | 98 |
| 6 | Placebo | 0 | 0 | 89 |
| 8 | Placebo | 0 | 0 | 90 |
| 9 | Placebo | 0 | 0 | 98 |
| 10 | Placebo | 0 | 0 | 97 |
| 11 | Placebo | 0 | 0 | 97 |
| 15 | Placebo | 1 | 1 | 92 |
| 16 | Placebo | 0 | 0 | 91 |
| 20 | Placebo | 2 | 2 | 91 |
| 22 | Placebo | 1 | 1 | 91 |
| 25 | Placebo | 0 | 0 | 98 |
| 28 | Placebo | 0 | 0 | 98 |
| 30 | Placebo | 2 | 2 | 86 |
| 31 | Placebo | 0 | 0 | 98 |
| 32 | Placebo | 0 | 0 | 97 |
| 35 | Placebo | 0 | 0 | 97 |
| 36 | Placebo | 0 | 0 | 80 |
| 39 | Placebo | 0 | 0 | 72 |
| 40 | Placebo | 0 | 0 | 96 |
| 42 | Placebo | 0 | 0 | 68 |
| 44 | Placebo | 0 | 0 | 85 |
| 45 | Placebo | 0 | 0 | 94 |
| 47 | Placebo | 1 | 1 | 94 |
| 49 | Placebo | 0 | 0 | 98 |
| 51 | Placebo | 0 | 0 | 92 |
| 54 | Placebo | 0 | 0 | 97 |
| 55 | Placebo | 0 | 0 | 98 |
| 60 | Placebo | 0 | 0 | 98 |
| 61 | Placebo | 0 | 0 | 68 |
| 62 | Placebo | 0 | 0 | 87 |
| 65 | Placebo | 0 | 0 | 86 |
| 67 | Placebo | 0 | 0 | 94 |
| 68 | Placebo | 1 | 1 | 90 |
| 70 | Placebo | 1 | 2 | 92 |
| 72 | Placebo | 0 | 0 | 98 |
| 73 | Placebo | 0 | 0 | 97 |
| 75 | Placebo | 0 | 0 | 97 |
| 76 | Placebo | 0 | 0 | 30 |
| 79 | Placebo | 0 | 0 | 97 |
| 81 | Placebo | 0 | 0 | 31 |
| 82 | Placebo | 0 | 0 | 95 |
| 87 | Placebo | 0 | 0 | 56 |
| 89 | Placebo | 0 | 0 | 97 |
| 91 | Placebo | 0 | 0 | 93 |
| 94 | Placebo | 1 | 3 | 91 |
| 98 | Placebo | 0 | 0 | 19 |
| 99 | Placebo | 1 | 1 | 86 |
| 100 | Placebo | 2 | 2 | 87 |
| 102 | Placebo | 1 | 3 | 92 |
| 103 | Placebo | 0 | 0 | 98 |
| 104 | Placebo | 1 | 2 | 64 |

#### Eye infection

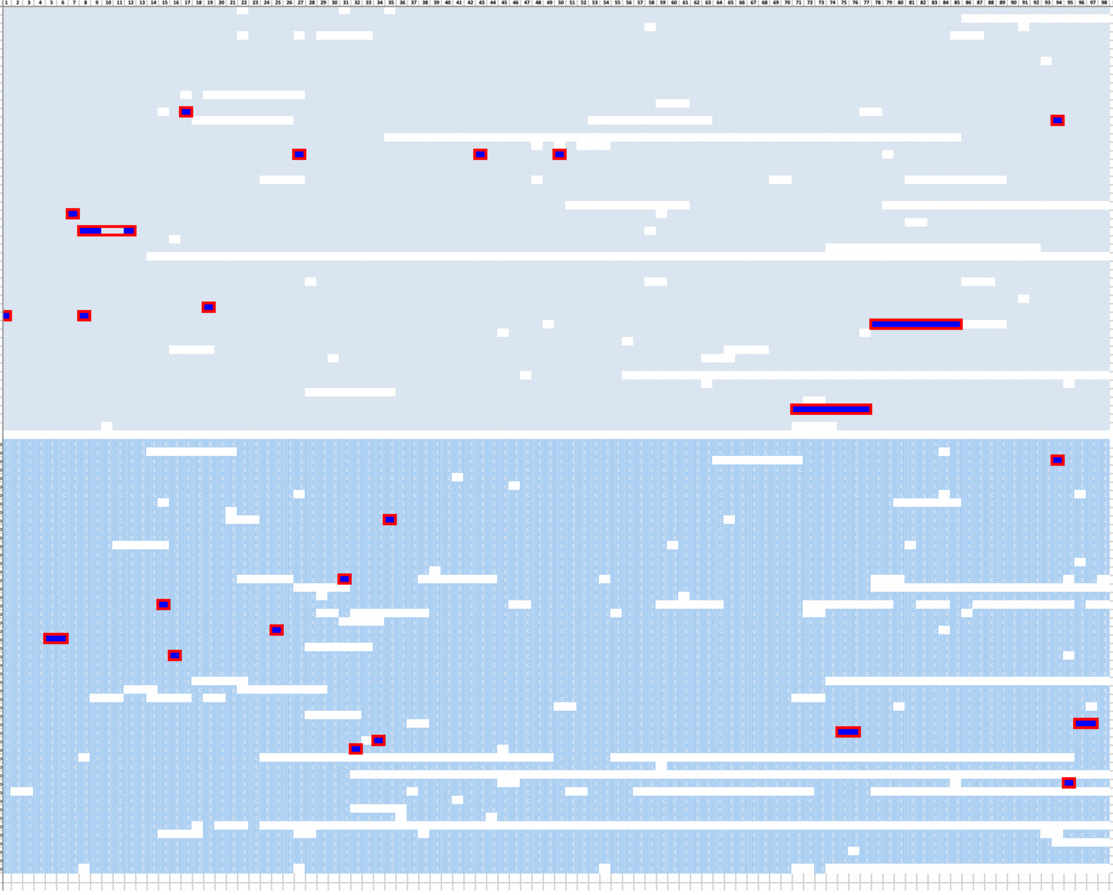

| ID | treat | N-epi | Total | obs |
|---|---|---|---|---|
| 3 | Iron | 0 | 0 | 95 |
| 4 | Iron | 0 | 0 | 85 |
| 5 | Iron | 0 | 0 | 96 |
| 7 | Iron | 0 | 0 | 88 |
| 12 | Iron | 0 | 0 | 98 |
| 13 | Iron | 0 | 0 | 98 |
| 14 | Iron | 0 | 0 | 97 |
| 17 | Iron | 0 | 0 | 98 |
| 18 | Iron | 0 | 0 | 98 |
| 19 | Iron | 0 | 0 | 98 |
| 21 | Iron | 0 | 0 | 88 |
| 23 | Iron | 0 | 0 | 95 |
| 24 | Iron | 1 | 1 | 92 |
| 27 | Iron | 1 | 1 | 75 |
| 29 | Iron | 0 | 0 | 98 |
| 33 | Iron | 0 | 0 | 47 |
| 34 | Iron | 0 | 0 | 93 |
| 37 | Iron | 3 | 3 | 88 |
| 38 | Iron | 0 | 0 | 98 |
| 41 | Iron | 0 | 0 | 98 |
| 43 | Iron | 0 | 0 | 82 |
| 46 | Iron | 0 | 0 | 98 |
| 48 | Iron | 0 | 0 | 98 |
| 50 | Iron | 0 | 0 | 67 |
| 52 | Iron | 1 | 1 | 94 |
| 53 | Iron | 0 | 0 | 96 |
| 56 | Iron | 1 | 5 | 90 |
| 57 | Iron | 0 | 0 | 97 |
| 58 | Iron | 0 | 0 | 79 |
| 59 | Iron | 0 | 0 | 13 |
| 63 | Iron | 0 | 0 | 98 |
| 64 | Iron | 0 | 0 | 98 |
| 66 | Iron | 0 | 0 | 92 |
| 69 | Iron | 0 | 0 | 98 |
| 71 | Iron | 0 | 0 | 97 |
| 74 | Iron | 1 | 1 | 95 |
| 77 | Iron | 2 | 2 | 92 |
| 78 | Iron | 1 | 8 | 86 |
| 80 | Iron | 0 | 0 | 96 |
| 83 | Iron | 0 | 0 | 97 |
| 84 | Iron | 0 | 0 | 90 |
| 85 | Iron | 0 | 0 | 94 |
| 86 | Iron | 0 | 0 | 98 |
| 90 | Iron | 0 | 0 | 54 |
| 92 | Iron | 0 | 0 | 96 |
| 93 | Iron | 0 | 0 | 90 |
| 95 | Iron | 0 | 0 | 96 |
| 96 | Iron | 1 | 7 | 89 |
| 97 | Iron | 0 | 0 | 98 |
| 105 | Iron | 0 | 0 | 93 |
| 1 | Placebo | 0 | 0 | 98 |
| 6 | Placebo | 0 | 0 | 89 |
| 8 | Placebo | 1 | 1 | 87 |
| 9 | Placebo | 0 | 0 | 98 |
| 10 | Placebo | 0 | 0 | 97 |
| 11 | Placebo | 0 | 0 | 97 |
| 15 | Placebo | 0 | 0 | 95 |
| 16 | Placebo | 0 | 0 | 91 |
| 20 | Placebo | 0 | 0 | 97 |
| 22 | Placebo | 1 | 1 | 91 |
| 25 | Placebo | 0 | 0 | 98 |
| 28 | Placebo | 0 | 0 | 98 |
| 30 | Placebo | 0 | 0 | 91 |
| 31 | Placebo | 0 | 0 | 98 |
| 32 | Placebo | 0 | 0 | 97 |
| 35 | Placebo | 0 | 0 | 97 |
| 36 | Placebo | 1 | 1 | 77 |
| 39 | Placebo | 0 | 0 | 72 |
| 40 | Placebo | 0 | 0 | 96 |
| 42 | Placebo | 1 | 1 | 65 |
| 44 | Placebo | 0 | 0 | 85 |
| 45 | Placebo | 0 | 0 | 94 |
| 47 | Placebo | 1 | 1 | 94 |
| 49 | Placebo | 1 | 2 | 94 |
| 51 | Placebo | 0 | 0 | 92 |
| 54 | Placebo | 1 | 1 | 94 |
| 55 | Placebo | 0 | 0 | 98 |
| 60 | Placebo | 0 | 0 | 98 |
| 61 | Placebo | 0 | 0 | 68 |
| 62 | Placebo | 0 | 0 | 87 |
| 65 | Placebo | 0 | 0 | 86 |
| 67 | Placebo | 0 | 0 | 94 |
| 68 | Placebo | 0 | 0 | 93 |
| 70 | Placebo | 1 | 2 | 94 |
| 72 | Placebo | 1 | 2 | 94 |
| 73 | Placebo | 1 | 1 | 94 |
| 75 | Placebo | 1 | 1 | 94 |
| 76 | Placebo | 0 | 0 | 30 |
| 79 | Placebo | 0 | 0 | 97 |
| 81 | Placebo | 0 | 0 | 31 |
| 82 | Placebo | 1 | 1 | 92 |
| 87 | Placebo | 0 | 0 | 56 |
| 89 | Placebo | 0 | 0 | 97 |
| 91 | Placebo | 0 | 0 | 93 |
| 94 | Placebo | 0 | 0 | 96 |
| 98 | Placebo | 0 | 0 | 19 |
| 99 | Placebo | 0 | 0 | 89 |
| 100 | Placebo | 0 | 0 | 93 |
| 102 | Placebo | 0 | 0 | 97 |
| 103 | Placebo | 0 | 0 | 98 |
| 104 | Placebo | 0 | 0 | 68 |

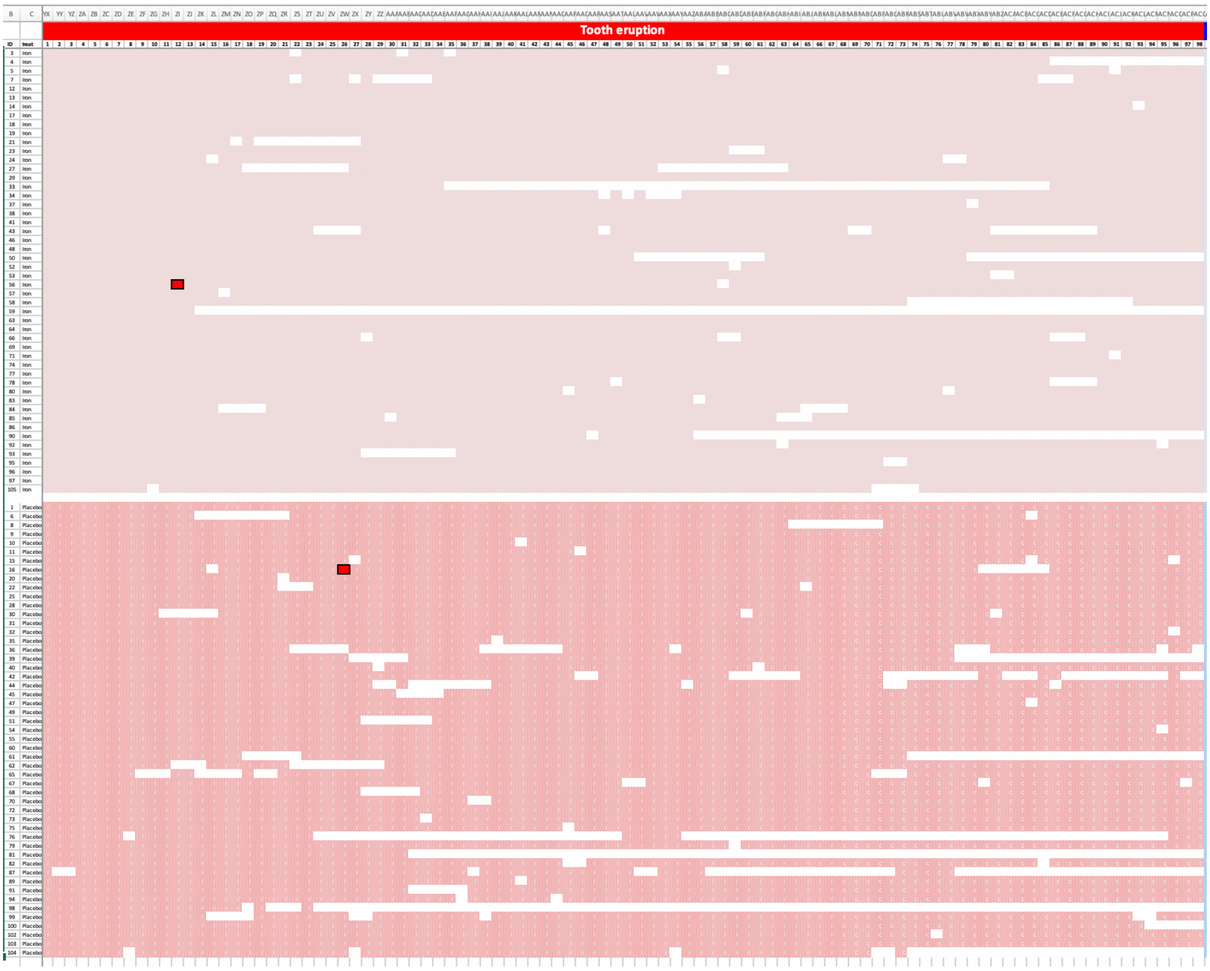
Tooth eruption
ID
treat
Iron
Placebo

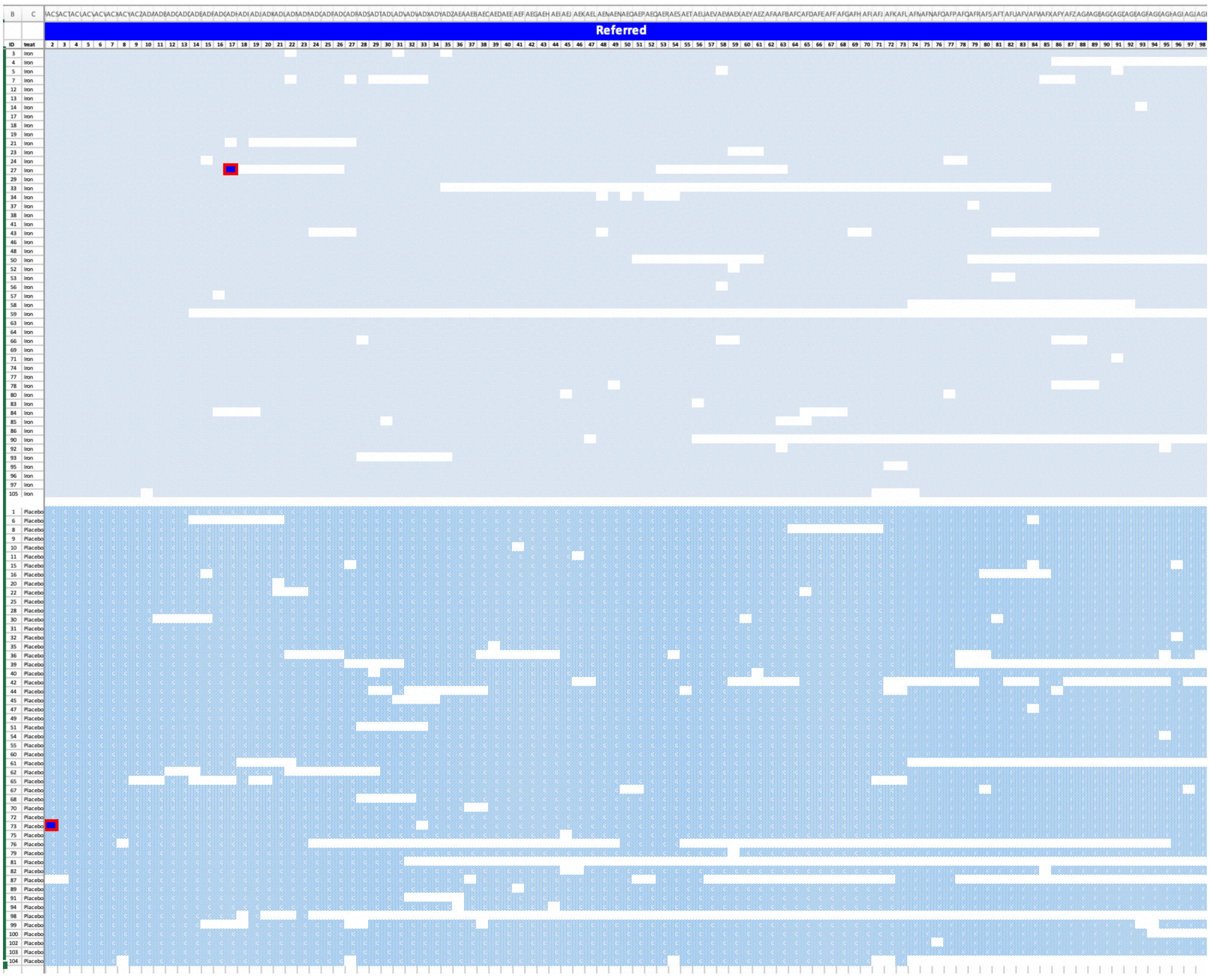
Referred

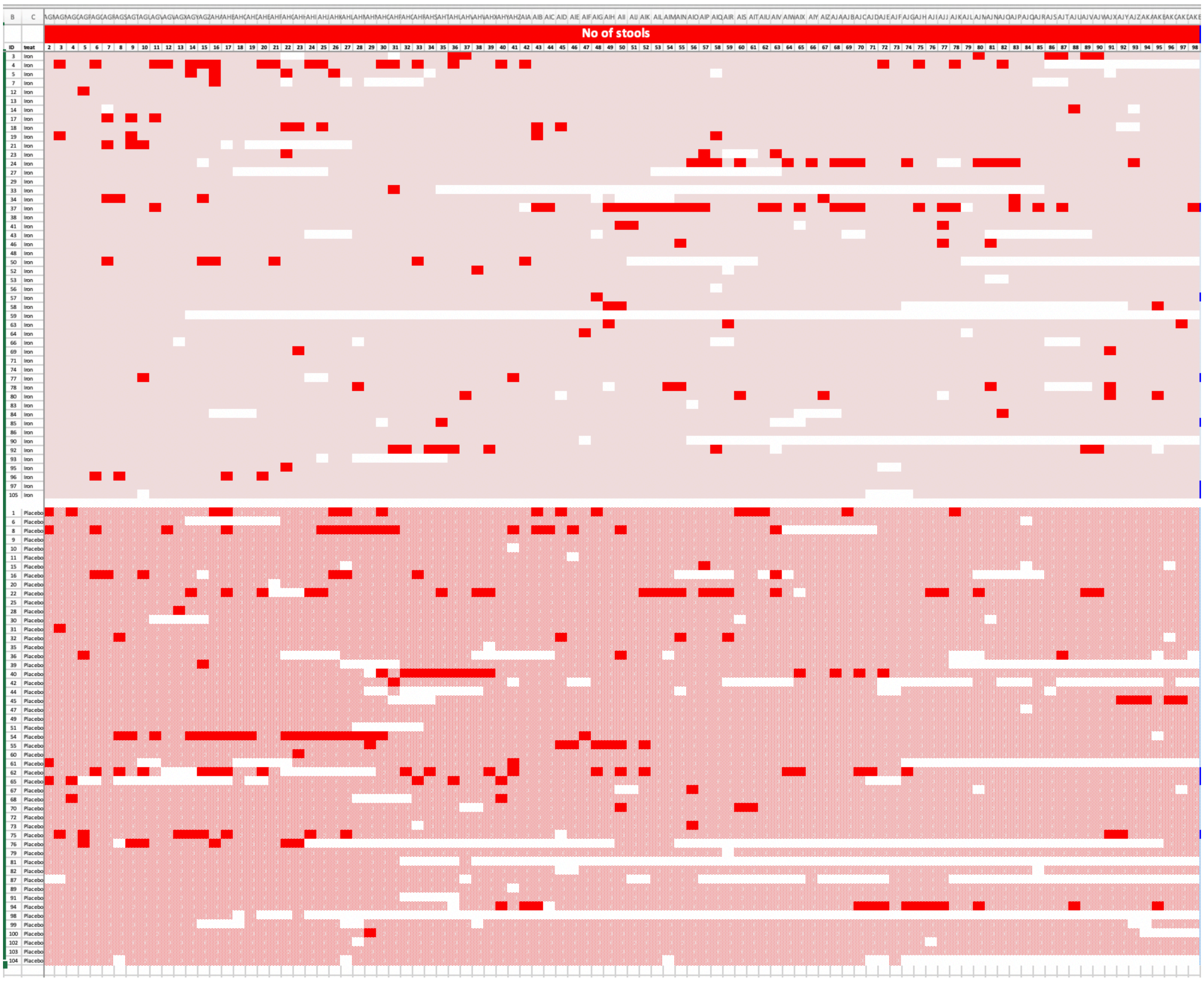
No of stools
ID
treat
Iron
Placebo

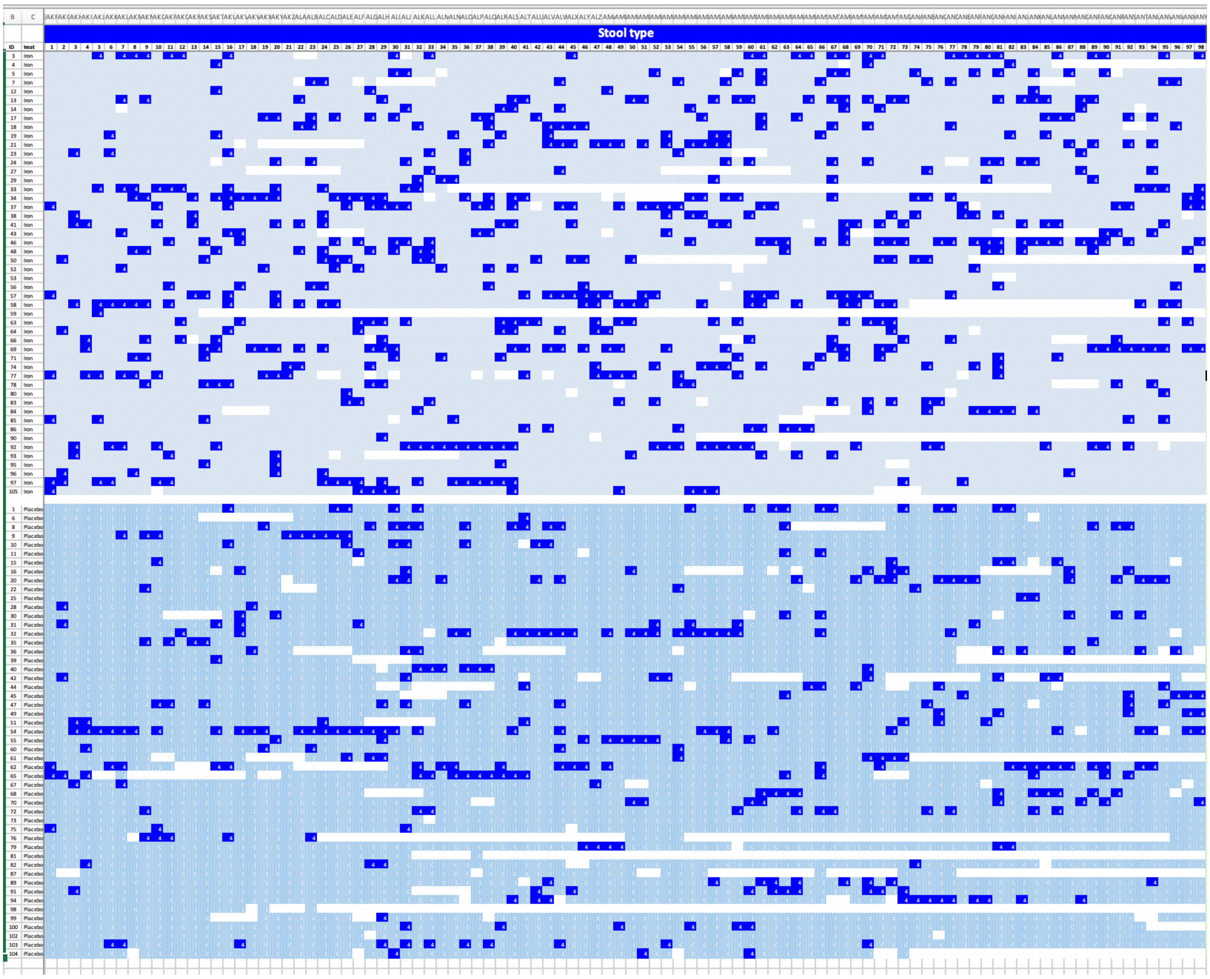
Stool type

#### Diarrhoea

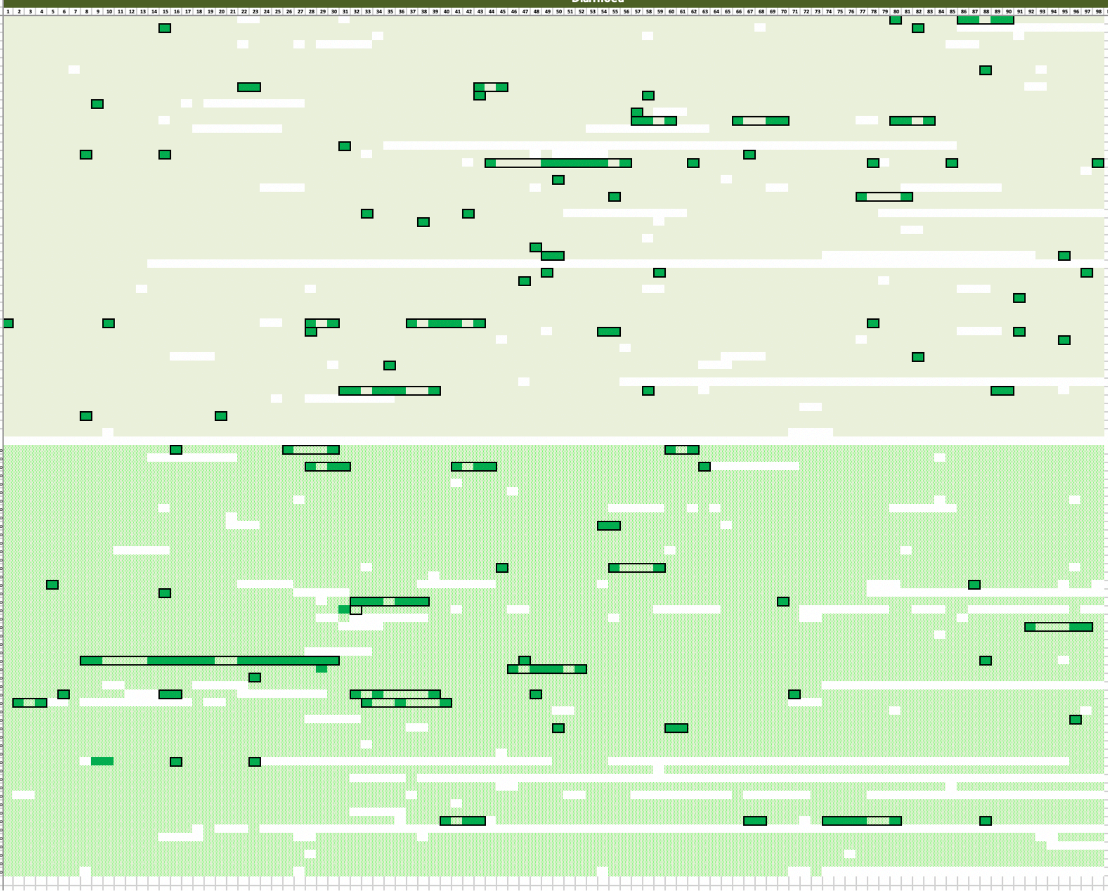

| ID | treat | N-epi | Totd | obs |
|---|---|---|---|---|
| 3 | Iron | 2 | 7 | 80 |
| 4 | Iron | 2 | 2 | 74 |
| 5 | Iron | 0 | 0 | 95 |
| 7 | Iron | 0 | 0 | 88 |
| 12 | Iron | 0 | 0 | 98 |
| 13 | Iron | 0 | 0 | 98 |
| 14 | Iron | 1 | 1 | 92 |
| 17 | Iron | 0 | 0 | 98 |
| 18 | Iron | 2 | 5 | 83 |
| 19 | Iron | 2 | 2 | 88 |
| 21 | Iron | 1 | 1 | 83 |
| 23 | Iron | 1 | 1 | 93 |
| 24 | Iron | 2 | 10 | 77 |
| 27 | Iron | 0 | 0 | 79 |
| 29 | Iron | 0 | 0 | 98 |
| 33 | Iron | 1 | 1 | 44 |
| 34 | Iron | 3 | 3 | 76 |
| 37 | Iron | 4 | 16 | 65 |
| 38 | Iron | 0 | 0 | 97 |
| 41 | Iron | 1 | 1 | 92 |
| 43 | Iron | 0 | 0 | 82 |
| 46 | Iron | 2 | 6 | 84 |
| 48 | Iron | 0 | 0 | 98 |
| 50 | Iron | 2 | 2 | 57 |
| 52 | Iron | 1 | 1 | 92 |
| 53 | Iron | 0 | 0 | 96 |
| 56 | Iron | 0 | 0 | 97 |
| 57 | Iron | 1 | 1 | 93 |
| 58 | Iron | 2 | 3 | 68 |
| 59 | Iron | 0 | 0 | 13 |
| 63 | Iron | 3 | 3 | 83 |
| 64 | Iron | 1 | 1 | 92 |
| 66 | Iron | 0 | 0 | 91 |
| 69 | Iron | 1 | 1 | 93 |
| 71 | Iron | 0 | 0 | 98 |
| 74 | Iron | 0 | 0 | 98 |
| 77 | Iron | 5 | 13 | 63 |
| 78 | Iron | 3 | 4 | 77 |
| 80 | Iron | 1 | 1 | 93 |
| 83 | Iron | 0 | 0 | 97 |
| 84 | Iron | 1 | 1 | 85 |
| 85 | Iron | 1 | 1 | 89 |
| 86 | Iron | 0 | 0 | 98 |
| 90 | Iron | 0 | 0 | 54 |
| 92 | Iron | 3 | 12 | 74 |
| 93 | Iron | 0 | 0 | 89 |
| 95 | Iron | 0 | 0 | 96 |
| 96 | Iron | 2 | 2 | 88 |
| 97 | Iron | 0 | 0 | 98 |
| 105 | Iron | 0 | 0 | 93 |
| 1 | Placebo | 3 | 9 | 77 |
| 6 | Placebo | 0 | 0 | 89 |
| 8 | Placebo | 3 | 9 | 72 |
| 9 | Placebo | 0 | 0 | 98 |
| 10 | Placebo | 0 | 0 | 97 |
| 11 | Placebo | 0 | 0 | 97 |
| 15 | Placebo | 0 | 0 | 95 |
| 16 | Placebo | 0 | 0 | 84 |
| 20 | Placebo | 0 | 0 | 97 |
| 22 | Placebo | 1 | 2 | 88 |
| 25 | Placebo | 0 | 0 | 98 |
| 28 | Placebo | 0 | 0 | 98 |
| 30 | Placebo | 0 | 0 | 91 |
| 31 | Placebo | 0 | 0 | 98 |
| 32 | Placebo | 2 | 6 | 82 |
| 35 | Placebo | 0 | 0 | 97 |
| 36 | Placebo | 2 | 2 | 70 |
| 39 | Placebo | 1 | 1 | 67 |
| 40 | Placebo | 2 | 8 | 81 |
| 42 | Placebo | 1 | 1 | 62 |
| 44 | Placebo | 0 | 0 | 85 |
| 45 | Placebo | 1 | 6 | 84 |
| 47 | Placebo | 0 | 0 | 97 |
| 49 | Placebo | 0 | 0 | 98 |
| 51 | Placebo | 0 | 0 | 92 |
| 54 | Placebo | 3 | 25 | 60 |
| 55 | Placebo | 2 | 8 | 82 |
| 60 | Placebo | 1 | 1 | 93 |
| 61 | Placebo | 0 | 0 | 66 |
| 62 | Placebo | 5 | 13 | 54 |
| 65 | Placebo | 2 | 11 | 66 |
| 67 | Placebo | 0 | 0 | 94 |
| 68 | Placebo | 1 | 1 | 88 |
| 70 | Placebo | 2 | 3 | 85 |
| 72 | Placebo | 0 | 0 | 98 |
| 73 | Placebo | 0 | 0 | 97 |
| 75 | Placebo | 0 | 0 | 97 |
| 76 | Placebo | 3 | 4 | 19 |
| 79 | Placebo | 0 | 0 | 97 |
| 81 | Placebo | 0 | 0 | 32 |
| 82 | Placebo | 0 | 0 | 95 |
| 87 | Placebo | 0 | 0 | 57 |
| 89 | Placebo | 0 | 0 | 97 |
| 91 | Placebo | 0 | 0 | 93 |
| 94 | Placebo | 4 | 14 | 67 |
| 98 | Placebo | 0 | 0 | 19 |
| 99 | Placebo | 0 | 0 | 89 |
| 100 | Placebo | 0 | 0 | 93 |
| 102 | Placebo | 0 | 0 | 96 |
| 103 | Placebo | 0 | 0 | 98 |
| 104 | Placebo | 0 | 0 | 68 |
